# OphJev: endpoint-aware System One decisions for ophthalmic imaging and clinical text

**DOI:** 10.64898/2026.10.03.26364645

**Authors:** Zhicheng Du, Zhuo Deng, Lan Ma

## Abstract

Ophthalmic decisions require probabilities tied to a clinical question, a candidate set and an operating policy. We present OphJev, an endpoint-aware System One framework that combines frozen image and text representations, candidate scoring, schema-specific calibration and selective generalist queries for fundus and OCT imaging. Across eight external resources, multiple representation families, three generalist sizes and three seeds, controlled interventions separate the effects of wording, supervision and computation. Schema-aware affine calibration lowers mean held-out Brier loss and selective risk in all eight primary image–text configurations. On HYGD, matched-supervision fusion raises AUROC from 0.772 to 0.798, while a separate gain-routing policy lowers Brier from 0.618 to 0.599 using at most 20% logical generalist image calls. These effects demonstrate complementary decision-level capabilities rather than universal gains: external transfer and larger specialist banks remain cohort dependent. A retrospective text extension evaluates 1,996 clinical notes and 91 ophthalmic questions; validation-based calibration improves note probability scores, while a lexical control remains strong. OphJev provides an auditable decision framework; prospective clinical benefit remains unvalidated.

## 1 Introduction

Ophthalmic artificial intelligence increasingly offers strong image representations, yet applying their outputs requires a precise clinical question. A glaucoma referral decision, a clinical glaucoma diagnosis, and the distribution of observed reader responses are different targets. The same photograph can support these questions without making their probability scales interchangeable. OCT adds a related challenge: a study’s normal category does not imply absence of all retinal disease, and a subject-level endpoint may require aggregation before restricting the candidate set.

Decision-native interfaces such as visual Jev and open typed-decision systems motivate fast, bounded alternatives to free-form generation [1–4]. OmniMed-Jev investigates medical candidate-conditioned probabilities across heterogeneous tasks, including ophthalmic datasets [5]; EyeAgent addresses a complementary agentic orchestration setting [6]. Retinal and vision–language foundation models supply specialist representations [7–9]. Together, these developments motivate a domain-specific question: how can a fast ophthalmic decision layer preserve the meaning of its outputs as candidate wording, clinical endpoint and computation policy change?

OphJev answers this question with explicit endpoint profiles, a shared candidate-scoring design, reserved-data calibration and budgeted generalist–specialist fusion (Fig. 1). We evaluate language against capacity-matched candidate identities, separate reader-response targets from clinical labels, and compare predicted query benefit with uncertainty and equal-budget random selection. Fundus and OCT provide two imaging modalities; BERT and other text encoders provide controlled candidate-language representations. The contribution is a reproducible ophthalmic decision framework and intervention-based evidence about when its components help. A separate text extension tests the same explicit-decision design on clinical notes and ophthalmic questions. The system combines existing representations rather than introducing a jointly pretrained foundation model or claiming first-in-field priority.

**Figure 1:**
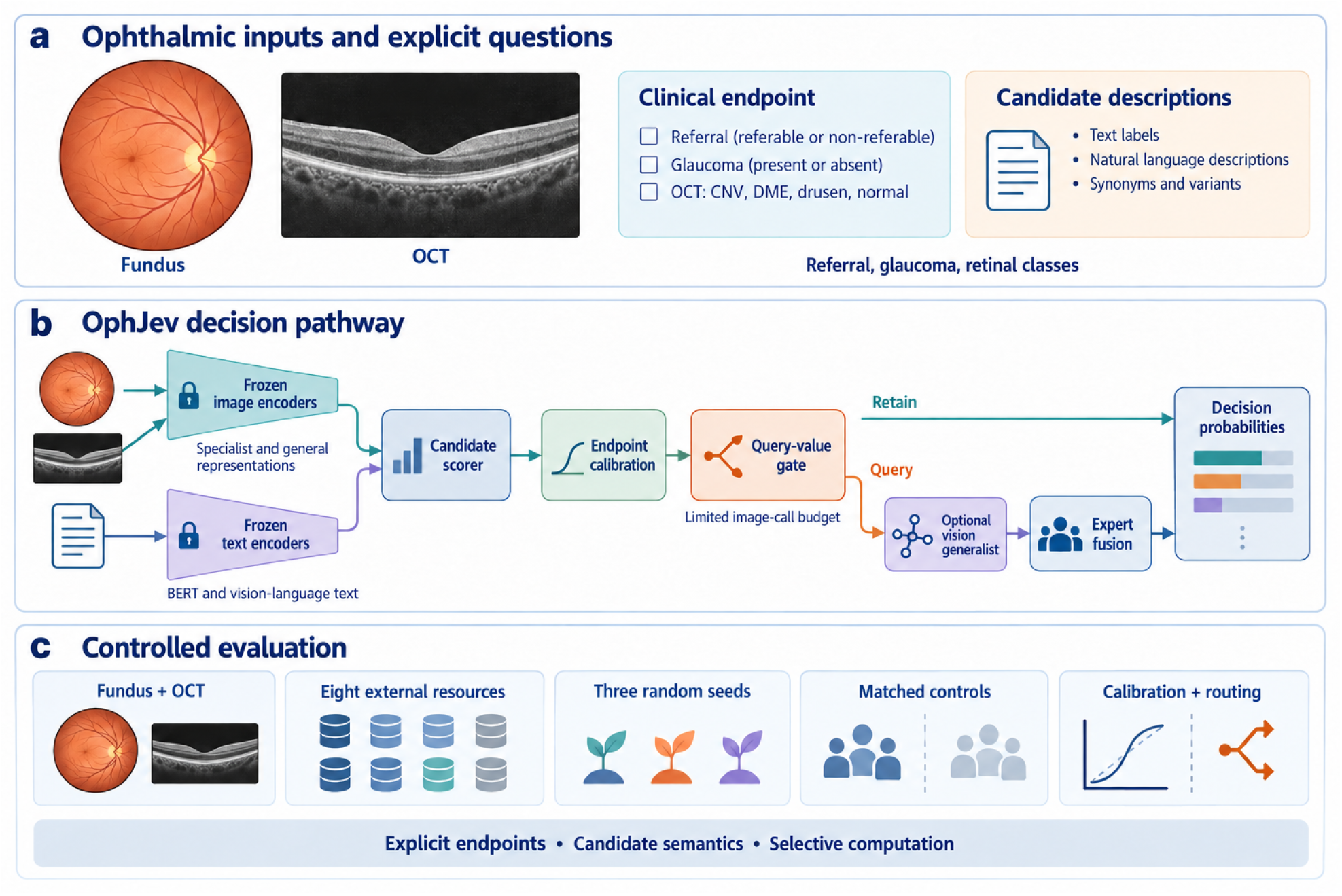
OphJev connects ophthalmic evidence to explicit decisions. (a) Fundus/OCT images and candidate descriptions define the task. (b) Frozen representations support candidate readouts, endpoint-specific calibration and optional generalist queries; the primary system also includes a conventional linear specialist. Separate task profiles share an interface rather than one jointly trained checkpoint. (c) Matched controls, eight external resources and three seeds test probability quality and computation policies. Ocular thumbnails are AI-assisted conceptual illustrations, not study images; bars are schematic and encode no measured probabilities.

## 2 Results

### A common interface retains the meaning of distinct ophthalmic tasks

The source resources comprise 101,423 aligned fundus images and 109,309 OCT entries before OCT duplicate/overlap exclusions; eight external resources test transfer. The evaluated framework covers fundus referral, external glaucoma/suspicion endpoints, reader-response targets, OCT category prediction, and a subject-level DME/normal endpoint. Candidate descriptions are encoded by vision–language or BERT-family text encoders and tested under held-out wording. These imaging experiments are complemented by a separate retrospective clinical-note and question-answering extension below; candidate-language encoders and patient-record inputs are evaluated as distinct uses of text. Figure 2 makes the endpoint contract explicit before presenting the quantitative effects.

**Figure 2:**
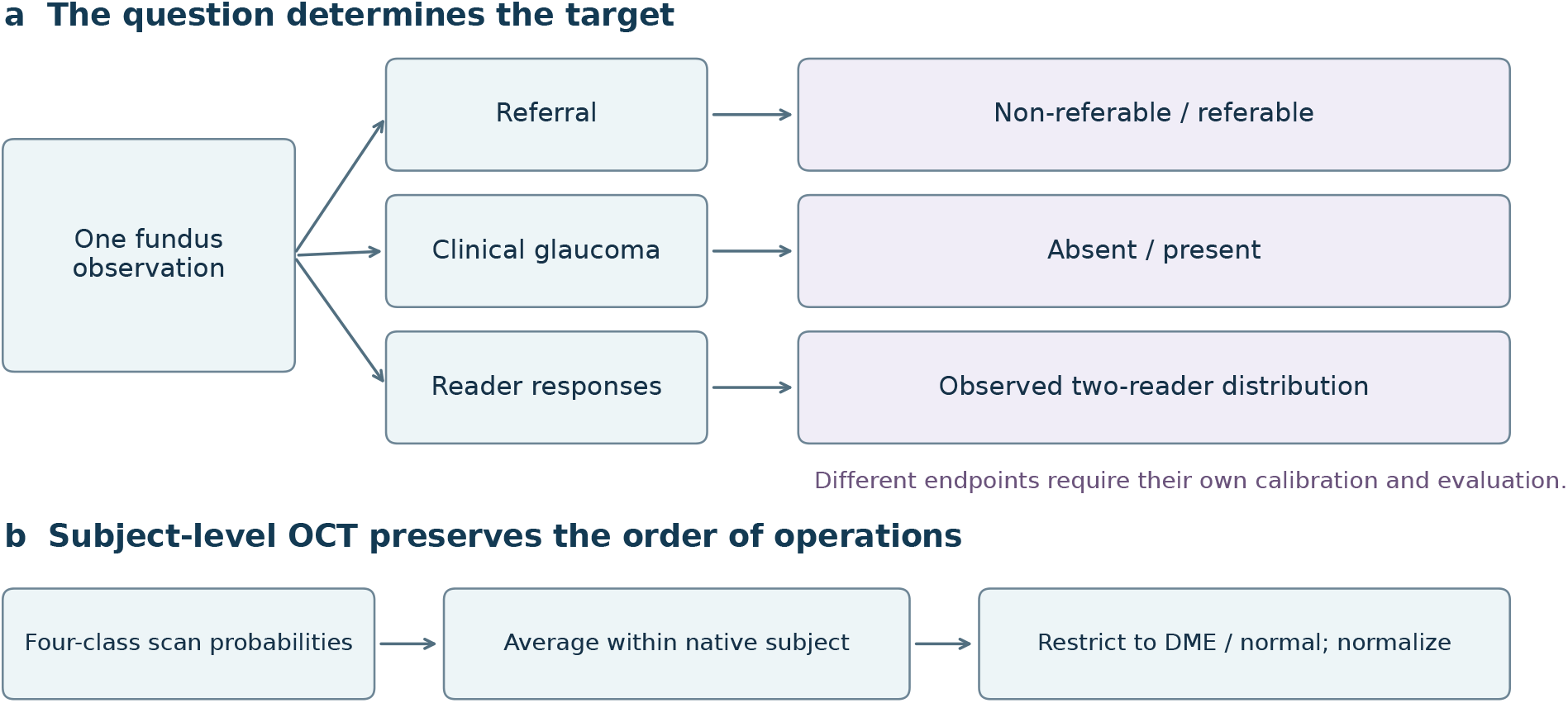
Endpoint semantics determine the evaluated decision. (a) The same fundus observation can support different candidate sets and supervision targets; referral, clinical disease and reader responses are not interchangeable labels. (b) For the subject-level OCT endpoint, averaging four-class scan probabilities precedes restriction and renormalization to DME/normal. This schematic describes protocol choices, not additional clinical outcomes.

### 2.1 Selective generalist queries improve complementary decision objectives

OphJev combines two frozen ophthalmic specialist readouts with a general-purpose visual Decision model, using separate explicit fundus and OCT endpoint profiles (Fig. 1). A shared interface returns aligned probabilities and records which images require the generalist. It neither infers the intended clinical endpoint from the image nor jointly pretrains a new foundation model. Three generalist sizes, three seeds, and eight resources yield 576 evaluation rows across eight policy/control arms. The extension includes 216 primary calibration/mixture fits, 432 auxiliary cross-fitting fits, and 72 small gain regressions; these are not 720 independent foundation-model training runs.

With the 2B generalist, fusion raises HYGD AUROC from 0.7715 to 0.7979, with paired difference +0.0263 (95% cluster interval [0.0118, 0.0421]). This is a gain over a specialist mixture fitted on the same external training cohorts. Brier changes from 0.6183 to 0.6208, and accuracy decreases from 53.87% to 48.44%. Against the earlier strong DINOv2-B control, which additionally uses source-development supervision, the fusion AUROC difference is +0.0085 with interval [−0.0292, 0.0450]; it does not establish superiority over that comparator. Fusion’s Brier is lower than that historical control by 0.0451 (interval [−0.0852, −0.0063]), but remains worse than the uniform binary predictor’s 0.5.

The gain router estimates whether fusion will reduce Brier using specialist outputs available before a generalist query. At an image budget of at most 20%, HYGD Brier falls from 0.6183 to 0.5994 (difference −0.0189, interval [−0.0278, −0.0107]). A label-independent hash gate with exactly the same call count has Brier 0.6181; the gain gate’s paired advantage is −0.0187 (interval [−0.0258, −0.0114]). In contrast, entropy-budget routing increases Brier by 0.0318. These comparisons support distinguishing expected benefit from uncertainty on this cohort. Gain routing does not reproduce fusion’s ranking gain: its HYGD AUROC is 0.7670. They are different operating policies, not simultaneous improvements from one model.

Table 1 reports all eight cohorts. Fusion and query policies improve different objectives; gains are cohort dependent. On HYGD, zero-shot StartLux 2B still has lower absolute Brier (0.5007) than the unified policies, so the proposed gains do not establish best absolute calibration. Complete controls, all generalist sizes and paired intervals are retained in the supplement.

**Table 1:** Three-seed means for the 2B profile. Each AUROC/Brier pair uses the same cohort endpoint. Gain routing caps logical generalist image calls at 20%; no universal improvement is implied.

| Resource | Specialist<br>AUROC/Brier | Fusion<br>AUROC/Brier | Gain routing<br>AUROC/Brier | Calls (%)<br>Gain |
| --- | --- | --- | --- | --- |
| PAPILA | 0.824/0.379 | 0.832/0.346 | 0.826/0.387 | 19.9 |
| Chaksu | 0.814/0.284 | 0.822/0.264 | 0.815/0.292 | 20.0 |
| RIM-ONE | 0.732/0.529 | 0.720/0.504 | 0.719/0.524 | 13.7 |
| Drishti | 0.905/0.252 | 0.902/0.409 | 0.909/0.281 | 19.8 |
| HYGD | 0.772/0.618 | 0.798/0.621 | 0.767/0.599 | 19.9 |
| OCTDL | 0.999/0.028 | 0.999/0.026 | 0.999/0.028 | 20.0 |
| CAVRI-H5 | 0.977/0.297 | 0.977/0.297 | 0.977/0.297 | 19.9 |
| Srinivasan | 1.000/0.081 | 1.000/0.081 | 1.000/0.080 | 20.0 |

The budget is defined on images, not patients or clinician referrals. On the full 45-subject Srinivasan stream, gain routing selects 19.99% of scans but touches 94.81% of subjects on average; the reported DME/normal performance uses only 30 eligible subjects. A fixed source entropy threshold also fails to preserve a fixed external call fraction. All policy outcomes use cached prediction replay, with lazy-callback implementation checks; no measured wall-clock speedup or reduction in clinical workload is claimed.

### 2.2 Candidate-language calibration improves decision quality

#### Schema-aware calibration improves proper scores and selective risk across the primary configurations

The complete eight-setting comparison crosses two image encoders, two candidate-text encoders and two image modalities. Reusing the reserved schema-specific affine calibrators lowers mean held-out summed-class Brier and AURC in all eight settings (Table 2). For DINOv2-B/MedSigLIP fundus scoring, Brier falls from 0.0917 to 0.0464. For DINOv2-B/FLAIR-BERT, it falls from 0.2993 to 0.0516 and AURC from 0.3344 to 0.0387. In OCT, Brier changes are smaller: 0.2872 to 0.2728 for DINOv2-B/MedSigLIP, and 0.3051 to 0.2921 for MedSigLIP/FLAIR-BERT. These are averages over seeds and descriptions, not universal per-case or statistically significant per-setting improvements.

**Table 2:** Decision quality under held-out candidate descriptions. Each cell gives transferred canonical calibration / schema-aware affine calibration; bold identifies the better mean within that pair. Means cover all three seeds and all three held-out descriptions, with no configuration excluded. Brier is summed-class probability loss; the other quantities concern top-label correctness, not disease prevalence. The calibrator uses reserved labeled calibration cases. D denotes DINOv2-B, M denotes MedSigLIP, and B denotes FLAIR-BERT. Differences are descriptive and do not imply significance in every seed or description.

| Task | Image / text encoder | Brier ↓ | ECE ↓ | Reliability ↓ | Resolution ↑ | AURC ↓ |
| --- | --- | --- | --- | --- | --- | --- |
| Fundus | D / M | 0.0917 / **0.0464** | 0.1205 / **0.0046** | 0.0219 / **0.0002** | **0.0051** / 0.0048 | 0.0038 / **0.0037** |
| Fundus | D / B | 0.2993 / **0.0516** | 0.2337 / **0.0020** | 0.1246 / **0.0001** | **0.0046** / 0.0029 | 0.3344 / **0.0387** |
| Fundus | M / M | 0.0747 / **0.0402** | 0.1033 / **0.0034** | 0.0170 / **0.0001** | 0.0049 / **0.0051** | 0.0023 / **0.0023** |
| Fundus | M / B | 0.2740 / **0.0477** | 0.1657 / **0.0023** | 0.0914 / **0.0001** | **0.0053** / 0.0035 | 0.2828 / **0.0403** |
| Oct | D / M | 0.2872 / **0.2728** | **0.0934** / 0.0963 | **0.0129** / 0.0142 | **0.0282** / 0.0269 | 0.0730 / **0.0617** |
| Oct | D / B | 0.2565 / **0.2492** | **0.0737** / 0.0788 | **0.0088** / 0.0098 | 0.0253 / **0.0265** | 0.0617 / **0.0556** |
| Oct | M / M | 0.2522 / **0.2487** | **0.0703** / 0.0785 | **0.0079** / 0.0105 | 0.0258 / **0.0276** | 0.0579 / **0.0510** |
| Oct | M / B | 0.3051 / **0.2921** | **0.1045** / 0.1052 | 0.0165 / **0.0161** | **0.0273** / 0.0272 | 0.0814 / **0.0692** |

Figure 3 separates probability scale from useful confidence variation. Reliability error decreases in five of eight settings and ECE in four; the OCT reliability term can worsen even when proper score and selective risk improve. The mechanism is therefore more specific than a blanket increase in calibration quality: an explicit candidate schema enables a reserved-data correction of decision probabilities, while the chosen metric determines which part of that correction is beneficial.

**Figure 3:**
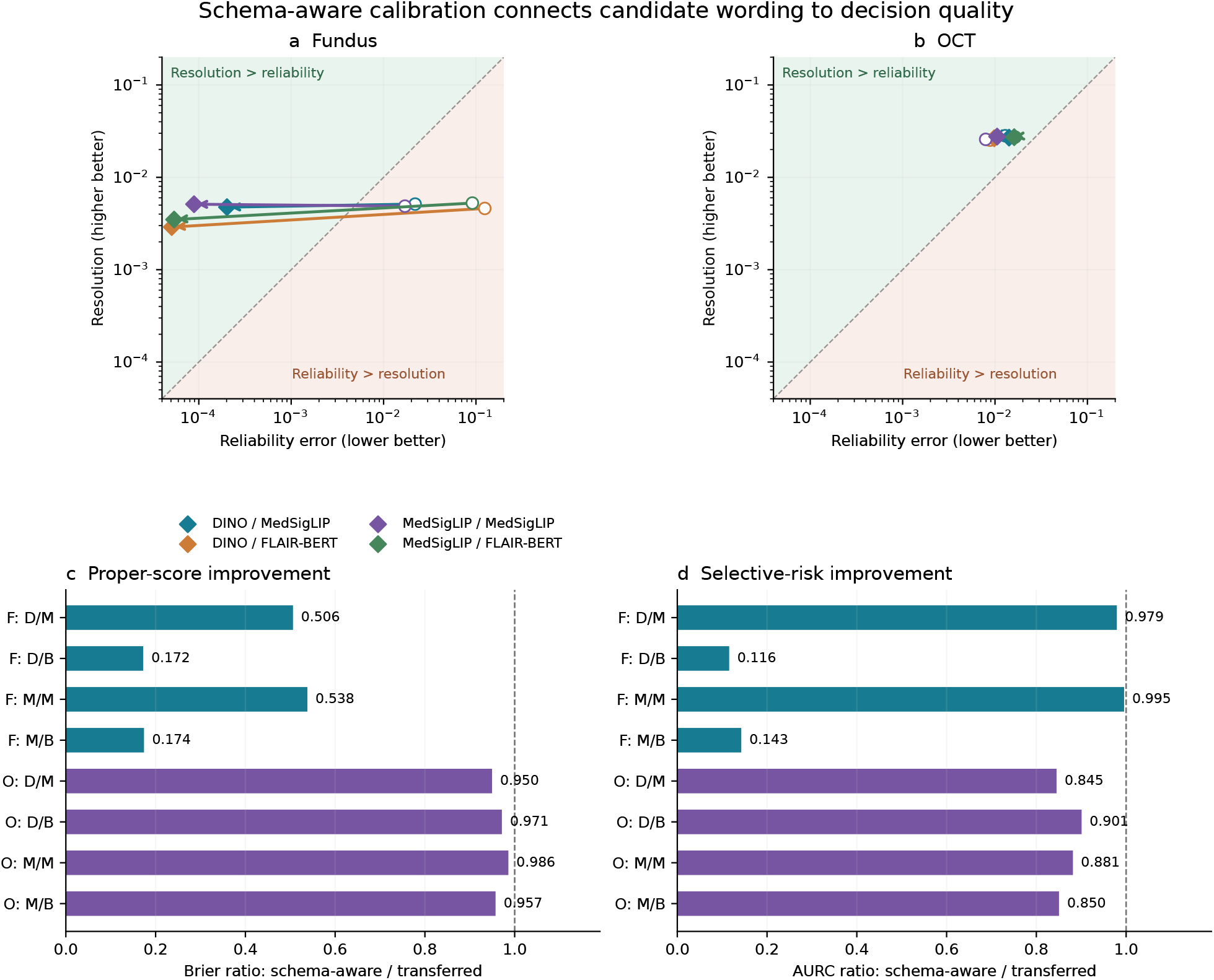
Decision quality across all primary image–text settings. (a,b) Arrows connect transferred canonical calibration (open circles) to schema-aware affine calibration (diamonds). Reliability and resolution concern top-label correctness; the diagonal is a bin-mean skill boundary, not a clinical threshold. (c,d) Ratios below one indicate lower summed-class Brier or AURC after schema-aware calibration. F/O denote fundus/OCT, D/M the DINOv2-B/MedSigLIP image encoder, and M/B the MedSigLIP/FLAIR-BERT text encoder. Means cover three seeds and three descriptions; each setting is retained. Ratios near one represent small effects.

The core experiment uses frozen DINOv2-B and MedSigLIP images with MedSigLIP or FLAIR-adapted clinical BERT text representations. Fixed classifiers, random candidate identities, text candidates, and text candidates with a training-template consistency penalty are evaluated across three seeds. Candidate heads do not uniformly outperform their matched controls; canonical AUROC spans approximately 0.918–0.953 for the fundus text arms and 0.987–0.992 for OCT.

For DINOv2-B/MedSigLIP fundus decisions, held-out descriptions change mean AUROC from 0.9213 to 0.9210 but increase Brier loss from 0.0476 to 0.0917. MedSigLIP/MedSigLIP exhibits the same distinction: AUROC changes from 0.9528 to 0.9527 while Brier rises from 0.0406 to 0.0747. In the evaluated model-upgrade cascade, wording raises the upgrade fraction from 30.1% to 81.8%; schema-specific calibration restores approximately 30.0%. This is measured model workload, not a simulated reduction in physician effort.

Across eight primary task/backbone/text settings, training consistency improves mean held-out Brier in three, schema-specific temperature in four, and schema-specific affine calibration in eight. These are aggregate setting counts, not universal per-seed or per-description gains. Temperature refitting worsens mean held-out Brier in all four primary OCT settings, where calibration and test class proportions differ. Positive temperature also cannot repair the reversed binary ranking observed in a BERT wording condition: affine calibration improves Brier from 0.2993 to 0.0516 while AUROC remains 0.6396. Figure 3 displays the probability changes; complete controls and intervals are retained in the supplement.

The external extension additionally evaluates pinned StartLux Decision 0.8B, 2B, and 4B vision checkpoints without ophthalmic training or labeled recalibration [10]. These are contextual zero-shot comparators, not supervision-matched controls. On HYGD their AUROC is 0.712, 0.741, 0.654 respectively; greater parameter count does not monotonically improve discrimination in this cohort. On subject-level DME/normal discrimination, the 0.8B model reaches AUROC 1.000 but accuracy 73.3%, illustrating that excellent ranking can coexist with a poorly placed decision boundary. Reversing candidate presentation while restoring class keys changes the 2B model’s argmax in 31.25% of 32 prespecified hash-selected HYGD images. This small mechanistic subset is descriptive, not an independent clinical validation. Complete cohort scores and paired contextual contrasts appear in the supplement. No optimized inference speed advantage is inferred from shared-GPU timings. On HYGD, the 2B model also has lower Brier than the matched-adaptation DINOv2-B linear control (difference −0.1652, 95% interval [−0.2301, −0.1026]), while their AUROC difference interval includes zero. This advantage for a general-purpose comparator is retained rather than interpreted as ophthalmic-model superiority.

#### Reader-response targets are a distinct estimand

The observed two-reader distribution is available for eligible fundus cases only. In 36 additional linear-probe fits, reader-distribution supervision lowers mean clinical Brier across three encoders, although paired intervals do not exclude zero for every encoder and seed. Adding a 0.1 Brier training term improves neither target arm in any of the nine encoder–seed settings within the fixed training budget.

Refitting clinical and reader temperatures on exactly the same eligible calibration cases reduces reader Brier for all nine reader-supervised probes. However, clinical Brier changes by only −0.000069, −0.000078, and +0.000348 for DINOv2-S, RETFound-CFP, and RETFound-OCT. The improvement against observed responses is consequently not interchangeable with improved referral prediction. Two readers do not identify population-level diagnostic uncertainty.

### 2.3 External adaptation repairs some scores but changes the source task

Eight external resources cover clinical glaucoma, photographic suspicion, OCT categories, and subject-level OCT discrimination. Their differing labels are retained rather than treated as equivalent diagnoses. Six initially evaluated cohorts subsequently become explicitly inspected development data. In the preceding development stage, a six-expert adapter fitted on permitted development cohorts was frozen before the original HYGD and Srinivasan evaluation. This historical adapter is distinct from the three-expert unified extension above. Later matched controls, the robust extension, and unified routing are exploratory.

On HYGD, the original adapter improves accuracy from 32.20% for MedSigLIP text to 50.25% and Brier from 1.232 to 0.738. However, its AUROC of 0.713 remains below the matched-adaptation DINOv2-B linear control’s approximately 0.789. Absolute calibration remains poor: a uniform binary predictor has summed-class Brier 0.5. At the frozen source operating threshold, limited sensitivity further restricts clinical usefulness. On the 30-subject DME/normal Srinivasan endpoint, every comparator reaches AUROC 1.0; adapted accuracy is 98.89%, tied with the original DINOv2-B text model. This small selected endpoint is saturated and cannot establish general superiority.

Applying the clinical adapter indiscriminately to the source referral task reduces fundus argmax accuracy from 97.53% to 90.31%, whereas source OCT accuracy improves from 85.20% to 91.03%. The 7.23-point fundus decline is an accuracy difference, not a sensitivity difference. Reusing the original screening branch when the requested endpoint is screening preserves its predictions exactly by construction; it does not prove that a learned adapter generalizes.

#### Encoder upgrades retain task-specific trade-offs

An exploratory extension adds four visual backbones and three candidate-text encoders, with 162 new readout fits and 954 external evaluation rows including matched linear controls. On HYGD, RETFound-DINOv3 text scoring reaches AUROC 0.7402 versus 0.6430 for its generic DINOv3-L architecture control. OphthaBERT v2 reaches 0.7432 with DINOv2-B images versus 0.6593 for MedSigLIP text. However, the dimension-matched OphthaBERT identity control reaches 0.7361, and language increases Brier from 1.2304 to 1.2432. The corresponding language–identity AUROC interval includes zero. These comparisons cannot assign the between-tower gain solely to medical semantics. In the fixed source wording test, OphthaBERT reduces the fundus Brier increment from 0.0441 to 0.0042, but increases the OCT increment from 0.0146 to 0.0263. RETFound-DINOv3 reader-target linear scoring reaches HYGD AUROC 0.7938, close to the original DINOv2-B reader-target linear result of 0.7904. Larger and more specialized encoders therefore provide complementary candidates rather than a universal replacement. Complete adverse results, preprocessing contracts, and paired intervals are in the supplement.

#### Medical representation controls reveal modality-specific transfer

Modality-matched MedKCO improves over swapped-modality weights, and an expanded expert bank gives small CAVRI improvements. Its HYGD decline shows that new specialist capacity requires endpoint-specific validation. The complete transfer matrix, paired intervals and negative-transfer figure are retained in the supplement; the original system remains the primary comparison.

### 2.4 Clinical text extends the interface while retaining strong conventional controls

Public text resources supply a separate endpoint-specific extension (Fig. 4). FairVLMed provides de-identified clinical notes and a visual-field-defined glaucoma label [11]. After exact normalized-note and common-input deduplication, the original subject partitions retain 6,946 training, 996 development and 1,996 test records. Inputs are restricted to the first 200 whitespace-delimited words. This is retrospective prediction of the dataset’s glaucoma label from clinical documentation, not a prospective symptom-to-diagnosis study or a literal diagnosis-extraction ground truth.

**Figure 4:**
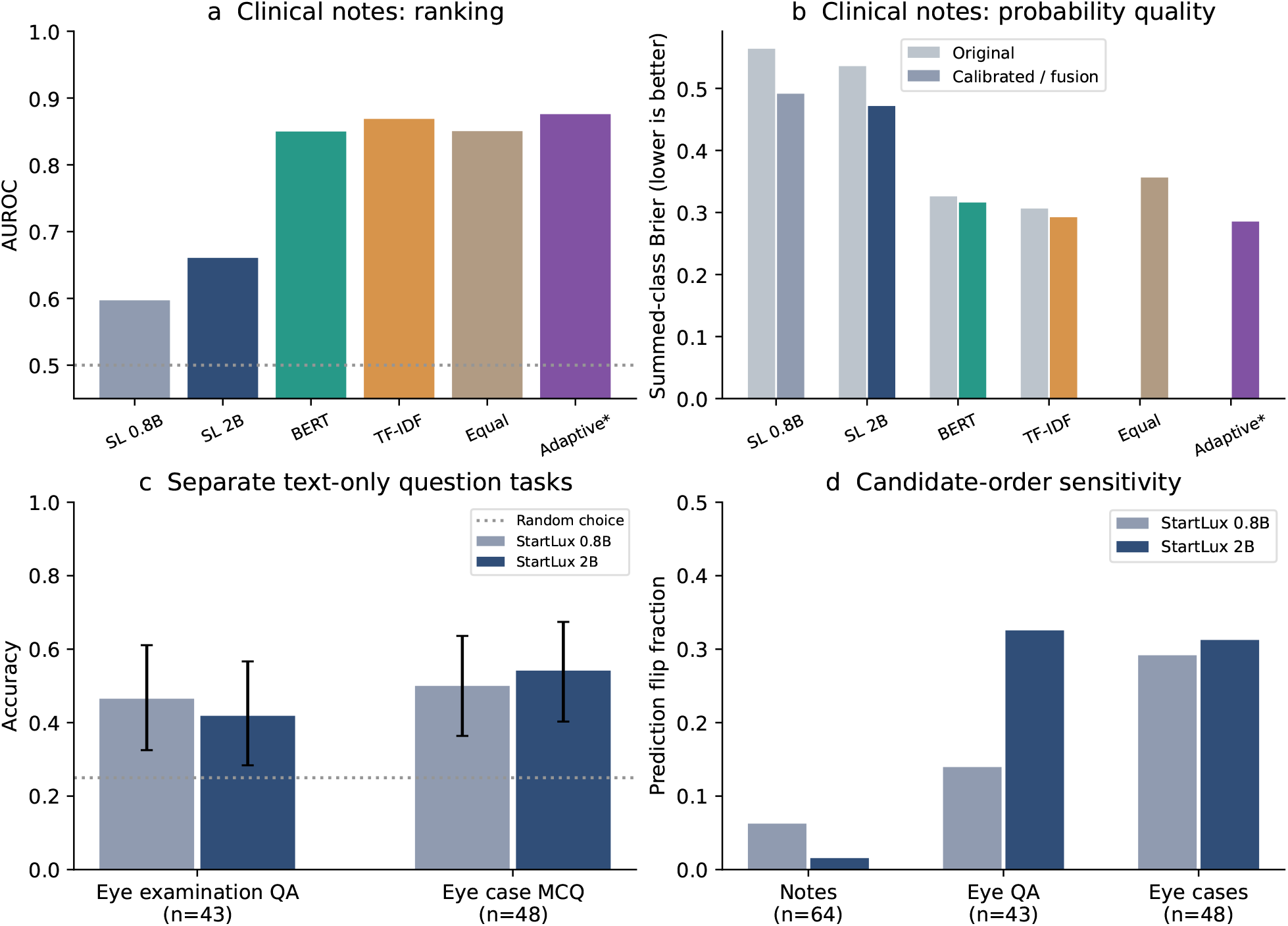
The text extension separates clinical-note prediction from ophthalmic questions. (a,b) All four note models and their probability-calibration effects, plus equal and exploratory adaptive mixtures (asterisk); the two supervised specialists use the original training partition, whereas StartLux is zero-shot before development calibration. (c) Two small question sets with Wilson 95% accuracy intervals, reported separately without pooling their accuracy into note performance. (d) Reversed-option prediction flips on a deterministic 64-note mechanism sample and all included question items. The dotted random-choice line in (c) is the expected accuracy of random selection, not the argmax accuracy of uniform probabilities. All plots use observed predictions.

On the note test set, TF–IDF logistic regression reaches AUROC 0.8688 and a frozen OphthaBERT linear probe reaches 0.8499. Zero-shot StartLux 0.8B and 2B reach 0.5971 and 0.6607, respectively; their lack of task supervision is explicit. Development-fitted temperature calibration lowers mean Brier from 0.3064 to 0.2923 for TF–IDF, from 0.3260 to 0.3161 for OphthaBERT, and from 0.5359 to 0.4716 for StartLux 2B, without changing within-model binary rankings. These means use three development-partition seeds, not independent patient cohorts or foundation-model training runs.

Prespecified equal fusion of calibrated OphthaBERT and StartLux 2B reaches AUROC 0.8504 and Brier 0.3564. Its Brier is worse than calibrated OphthaBERT by 0.0402 (paired 95% interval 0.0324–0.0486); a stronger generalist is therefore not automatically a useful probability-mixture component. A subsequent exploratory three-expert mixture learns nonnegative weights using development data only and reaches mean AUROC 0.8759 and Brier 0.2853. Against calibrated TF–IDF, the paired seed-mean probability differences are +0.00714 in AUROC (95% interval 0.00489–0.00961) and-0.00705 in Brier (interval-0.00997 to-0.00429). All three development partitions assign effectively zero weight to StartLux and retain complementary lexical and BERT experts. All weights and unchanged original comparisons are reported in the supplement. This additional design was introduced after inspecting the original test results and is not confirmatory.

Two smaller tasks retain their distinct interpretation: 43 official MedMCQA ophthalmology single-answer validation items [12], used only for evaluation, and 48 DiagnosisArena ophthalmic case questions selected by a fixed diagnostic vocabulary [13]. StartLux 0.8B/2B accuracy is 46.5%/41.9% on the former and 50.0%/54.2% on the latter. Reversing candidate presentation while restoring semantic class keys flips 14.0%/32.6% of examination answers and 29.2%/31.2% of case answers. These results demonstrate a functioning text decision interface and expose order sensitivity; they do not establish clinical diagnostic superiority. The original labels and options are retained; no physician adjudication of our automated case subset has been performed.

## 3 Discussion

OphJev connects ophthalmic candidate decisions to explicit endpoints and computation policies. Its components provide complementary, measurable capabilities: schema-aware calibration improves average proper score and selective risk across all eight primary representation settings, fusion improves HYGD discrimination relative to a matched specialist mixture, and gain routing improves a proper score under a limited image-call budget. These effects address different parts of a decision workflow. They support an ophthalmic System One interface whose outputs and costs can be audited, rather than a claim that one operating policy improves every metric or population.

The calibration mechanism is most visible when candidate wording changes confidence while largely preserving ranking. Reserved-data affine calibration corrects the resulting scale and class-intercept shifts, but the clinical question remains essential. Reliability error, resolution and class-probability loss describe different properties: ECE improves in only four of eight settings and reliability in five, despite the eight mean Brier and AURC improvements. High top-label correctness under class imbalance also need not indicate useful minority-class discrimination. A positive-slope binary calibrator cannot repair FLAIR-BERT’s reversed ranking. The accompanying mathematical identities explain these boundaries: monotone recalibration preserves ranking, resetting a threshold at the same quantile preserves the selected cases, and endpoint restriction generally cannot be interchanged with expert pooling. These are established consequences applied to the evaluated design, not new general learning theorems.

Selective computation depends on expert complementarity, not uncertainty alone. The gain gate learns from group-excluded predictions whether the alternative model is expected to reduce Brier loss. Its HYGD advantage over an equal-call hash gate supports this distinction, while the full cohort matrix shows that the learned relationship can fail under transfer. Full fusion worsens Drishti Brier from 0.2517 to 0.4091, and gain routing worsens PAPILA Brier from 0.3795 to 0.3871. On HYGD, raw zero-shot StartLux 2B has lower absolute Brier than the adapted policies. Thus additional supervision and computational expenditure do not ensure transportable calibration. Model-call budgets measure computational selection, not reductions in clinician workload or missed disease.

Specialist expansion exposes a related form of negative transfer. The augmented expert bank gives small CAVRI gains but lowers HYGD fusion AUROC from 0.7979 to 0.7474. MedKCO’s native category score reaches HYGD AUROC 0.7417, compared with 0.6715 after source-referral readout training; that comparison changes supervision and cannot causally isolate endpoint mismatch. These observations motivate retaining explicit task profiles and validating new experts against strong conventional controls. They do not justify selecting a different winning system after examining each test cohort. The distinction between referral, clinical disease and observed reader responses also prevents improvements against one target from being presented as equivalent clinical benefit [14].

The evidence remains retrospective. Public resources differ in labels, population composition and identifier completeness; the source OCT mirror supports image-hash isolation rather than verified patient-disjoint splitting. All external cohorts were inspected in earlier development, and the latest plots and expanded-bank comparisons are exploratory. Pretraining exposure is a separate concern: MedKCO recipe manifests include both source resources, PAPILA, Chaksu and Drishti; 495 matching Srinivasan paths cover all 45 subjects. Its results there are not unseen-patient validation, and absence from a named manifest does not prove absence from pretraining. Stored 224-pixel images constrain native-resolution comparisons, and researcher-authored candidate descriptions lack an independent clinical equivalence panel. The immediate translational requirement is a locked evaluation on a genuinely untouched, clinically characterized cohort, followed by clinician-assisted assessment of decision quality and workload. Prospective diagnosis from presentation-time text and patient benefit remain unvalidated; retrospective note and case-question outcomes are reported separately.

The text extension sharpens the same distinction between representations, endpoints and decision policies. Clinical documentation predicts a visual-field-defined label, so a strong lexical baseline can capture useful associations that a zero-shot decision model does not recover. Calibration improves probability scale, whereas equal generalist–specialist fusion can dilute a stronger specialist. On a fixed 64-note subset, masking glaucoma-related diagnosis tokens changes TF–IDF and OphthaBERT accuracy only from 81.3% to 79.7% and from 76.6% to 75.0%, respectively; this limited ablation does not remove treatment or examination proxies and cannot demonstrate prediagnostic validity. Public examination and case questions further expose candidate-order dependence. FairVLMed and OphthaBERT both involve Massachusetts Eye and Ear, so pretraining patient overlap cannot be excluded. The note cohort therefore does not establish pretraining-independent external generalization, and the automatically selected case subset remains unadjudicated.

## 4 Methods

### 4.1 Data, targets, and separation of roles

The source fundus resource aligns 101,423 images with released JustRAIGS labels, and OCT uses the four released CNV, DME, drusen, and normal categories [15, 16]. Fitting, development, calibration, threshold selection, and testing use disjoint stored group partitions. Fundus groups account for fellow-eye proxies; OCT groups preserve exact image hashes. Reader targets include only cases with two binary responses; missing and ungradable observations are excluded, and selectively observed adjudication is not treated as a third exchangeable reader.

The external resources are PAPILA, Cháks.u, RIM-ONE DL, Drishti-GS, OCTDL, CAVRI-H5, HYGD, and Srinivasan OCT. Original dataset citations, access conditions, masks, and split audits are provided in Supplementary Methods. HYGD contains 737 exact-unique images and 283 connected patient/pixel components after deduplication. Its primary endpoint remains image-level glaucoma with cluster-aware uncertainty; linked identifiers are not converted into an invented patient diagnosis. Srinivasan’s primary endpoint averages original four-class probabilities within each subject before conditioning on DME/normal; AMD is not relabeled CNV or drusen. CAVRI’s unsupported VMA–VMT category is excluded from supervised closed-class fitting. Cháks.u suspicion labels are excluded from clinical-glaucoma adapter fitting.

### 4.2 Candidate models and endpoint adapters

Frozen image features *v* and text features *t_c_* enter trainable projections, followed by a shared scorer of [*g_v_*(*v*)*, g_t_*(*t_c_*)*, g_v_*(*v*) ⊙ *g_t_*(*t_c_*)]. A softmax produces probabilities over the supplied closed candidate set. Training rotates three prespecified descriptions; three other descriptions are held out. Random identities match text-head trainable capacity, while fixed classifiers approximately match it. Text encoders remain frozen; alignment occurs in the small projection/scoring head rather than new foundation-model pretraining.

The original fusion uses six experts crossing two encoders with linear, fixed, and candidate readouts. The endpoint adapter predicts

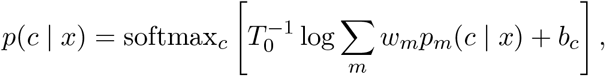

where *w* lies on the simplex, ∑*_c_ b_c_* = 0, and |*b_c_*| ≤ 2. Original expert temperatures and *T*_0_ remain fixed. Each cohort contributes equally, with equal connected-group weight within cohort. OCTDL contributes conditional DME/normal likelihood. Weight and bias penalties are 0.05∥*w* − *w*_0_∥^2^ and 0.01∥*b*∥^2^.

The robust extension minimizes the maximum regularized cohort NLL using an epigraph variable and analytic gradients. SLSQP checks convergence and constraint feasibility. This is a nonconvex optimization with a local solution; we do not claim a global minimax certificate. Old-cohort predictions use leave-one-cohort-out states; HYGD and Srinivasan use states fitted only on original permitted development cohorts. Source screening can explicitly dispatch to the original frozen source branch. Neither dispatch nor optimization assumes that all tasks improve.

### 4.3 Unified generalist–specialist decisions and selective computation

The unified system shares a decision interface and model family across explicit fundus and OCT endpoint profiles. It does not share a newly pretrained image backbone across tasks. Each profile combines the original DINOv2-B hard-label linear readout, MedSigLIP candidate readout, and a pinned StartLux vision model. Separate experiments use 0.8B, 2B, or 4B as the generalist. Their complete results remain reported. The 2B profile is designated for primary detailed routing analysis using previously inspected results and computational feasibility; this designation is exploratory rather than blind selection.

For each endpoint profile, calibrated probabilities take the form

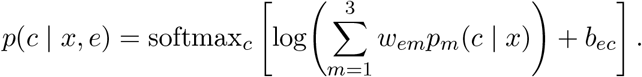

Weights are nonnegative and sum to one; class intercepts sum to zero and are bounded by two. The generalist, specialist, and fusion controls receive the same permitted external training cohorts and intercept constraints. The specialist control fixes the generalist weight to zero; the generalist control fixes its own weight to one. An untrained uniform probability mixture is also retained. Equal-cohort, equal-connected-group NLL is regularized by 0.05∥*w* − *w*_0_∥^2^ + 0.01∥*b*∥^2^, where the specialist/fusion anchor is (1*/*2, 1*/*2, 0) and the generalist anchor is (0, 0, 1). Partial OCTDL labels contribute conditional DME/normal likelihood. Source internal data are not included in this meta-fit because corresponding generalist predictions are unavailable.

Fundus adaptation uses only PAPILA, RIM-ONE, and Drishti; OCT adaptation uses OCTDL and CAVRI. Original-cohort evaluation excludes the evaluated cohort from fitting. HYGD and Srinivasan never enter these optimizers, but they have been inspected during prior analyses and are not new blinded tests. Across three sizes, three seeds, and eight task/fold combinations, the extension fits 216 primary mixture/calibrator arms.

The fixed entropy cascade calls the generalist above the training cohorts’ group-weighted 80th entropy percentile. This source threshold does not guarantee a 20% external call fraction. A separate batch policy selects the highest-entropy ⌊0.2*N* ⌋ images without using evaluation labels. The learned gain policy instead predicts the Brier reduction from replacing specialist output with fusion. Its features are the two original specialist probability vectors, their normalized entropies, and their *L*_1_ disagreement; no generalist output is available to the gate at query time.

Gain targets are obtained with three-fold connected-group cross-fitting within permitted training cohorts. Each target group’s labels are absent from the specialist and fusion calibration fits that produce its target. A weighted ridge regression with penalty 0.01 estimates the gain from standardized specialist features. The batch gate selects at most the top 20% of images with positive predicted gain. This adds 432 auxiliary calibration fits and 72 small gate fits, separately counted from primary models. A label-independent hash gate queries exactly the same number of images as each gain gate, controlling for budget alone.

All cascade metrics replay stored predictions. The reported query fraction is a conservative logical count before removing exactly zero-weight generalist components; it is not a wall-clock speedup. A lazy-callback implementation calls the generalist only for selected images and reproduces the archived outputs. Exactly zero-weight components require no callback. The 20% cap applies to image calls, and group-level coverage is reported separately. Original four-class OCT probabilities are retained until subject aggregation and endpoint conditioning. Pure-text clinical diagnosis is not a validated endpoint: the clinical BERT arm encodes candidate descriptions rather than patient records, and the local VOLMO case narratives require target-leakage and image-dependence review before serving as a text benchmark. The final medical-representation experiment adds MedKCO-CFP/OCT, MerMED, and DINOv3-S [17, 18] under the same source split and readout schedule (108 fits), together with 36 conventional linear fits. Both MedKCO image checkpoints are tested on both image types with common and native candidate encoders. Category-name versus official knowledge-description scoring is evaluated separately without target-driven prompt selection. Stored 224-pixel images are upsampled to 512 for MedKCO, so this is not a native-resolution reproduction of its published benchmark [19]. Fixed text padding avoids batch-dependent averaging in its released encoder.

Two additional expert-bank designs retain all original cohorts and endpoint masks. Replacement uses modality-matched MedKCO and MerMED; augmentation retains the original DINOv2/MedSigLIP experts and adds both medical experts. StartLux 2B remains the optional generalist. All arms receive the same permitted cohort supervision and intercept bounds. The augmented regularizer is anchored to the original two-expert weights; no HYGD or Srinivasan labels enter fitting. After observing individual-encoder results, augmentation was added as an explicitly exploratory test of retaining existing expertise. Fusion changes probabilities, not pretrained model weights. Multi-teacher distillation and compatible-adapter weight merging are different interventions [20, 21]. Query-benefit estimation is also established in related cascade work [22]; our gain gate does not inherit finite-sample clinical risk guarantees from those methods.

### 4.4 Outcomes and statistical analysis

Outcomes include AUROC, accuracy, summed-class Brier loss, NLL, and achieved operating-point sensitivity and specificity. Summed-class binary Brier is twice positive-class Brier. Seed summaries average metrics over three separately fitted models, not predictions from a seed ensemble. The core study retains its original paired group intervals. The robust and unified extensions use 500 paired connected-group bootstrap draws, shared across models and seeds, and reports percentile intervals without multiplicity adjustment. Subject-level resampling is used after Srinivasan aggregation. The source-threshold transport analysis uses 1,000 paired group draws and an illustrative 5% prevalence scenario. These analyses condition on fitted models and do not certify prospective generalization. Code, numerical provenance, and complete adverse results accompany the supplement.

#### Decision-quality visualization

We re-evaluate frozen source test predictions under three held-out descriptions, using the stored canonical and schema-specific affine calibrators. All eight image–text–modality configurations and all three seeds are retained; no calibrator is fitted to test cases. For top-label confidence *q_i_* = max*_c_ p_i_*(*c*) and correctness *z_i_*, ECE, reliability and resolution use 15 equal-width bins. Reliability is 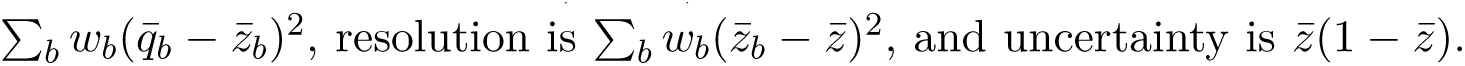 Their decomposition is exact for the Brier score of bin-mean confidence; the difference from the unbinned top-label score is explicitly reported as a binning residual. AURC averages cumulative classification error over confidence-ranked prefixes with stable tie ordering. These descriptive, post-hoc exhibits average metrics across descriptions and seeds rather than pooling repeated observations as independent patients. Paired intervals for the original fusion and routing comparisons remain unchanged.

#### Retrospective text protocol

FairVLMed defines non-glaucoma by normal visual field (mean deviation at least −1 dB with normal hemifield and pattern-standard-deviation results) and glaucoma by mean deviation below −3 dB with abnormal corresponding tests [11]. We retain source subject partitions and remove normalized-note duplicates with priority test, development, then training; the resulting common inputs also have no exact overlap. Original notes, not GPT-4 summaries, are used. TF–IDF and frozen OphthaBERT probes fit only on training records. Three SHA-256 partitions of the development set each provide 498 temperature-fitting and 498 held-out selection records. The primary text fusion is an equal probability mixture; a separately marked exploratory extension fits a three-expert simplex mixture and selects its regularization on the second development half. No test labels enter either fit. Public QA items receive no fitting or calibration. Mechanism controls reverse option order, omit evidence, or mask a declared diagnosis-token vocabulary. Confidence intervals resample test subjects/items 2,000 times; fusion comparisons use seed-mean probabilities. Full inclusion rules, metrics, resource decisions and runtime receipts accompany the source release.

## Data and code availability

Reproduction code and aggregate numerical results are included in the local submission package. Original datasets and pretrained weights must be obtained from their providers under the cited access terms. No native patient identifiers, raw clinical images, or per-image predictions are included in the public-facing archive. The accompanying code-and-results archive provides the aggregate outputs and reproduction entry points.

## Ethics and research transparency

This work reanalyzes existing released resources and does not recruit participants or intervene in care. Original resource governance and consent are described by their providers. No new institutional ethics approval or exemption number is reported for this secondary analysis. Authors remain responsible for verification.

## Data Availability

All data produced in the present study are available upon reasonable request to the authors

## Acknowledgements

This research was funded by Development and Reform Commission of Shenzhen Municipality (F-2024-Z99-503929), Shenzhen Science and Technology Innovation Commission (JCYJ20230807094803007), Shenzhen Medical Research Fund (C2401007), State Key Laboratory of Chemical Oncogenomics, Institute of Biomedical Health Technology and Engineering, Shenzhen Bay Laboratory. We thank the contributors of FairVLMed, MedMCQA, DiagnosisArena, the cited ophthalmic datasets, DINOv2, RETFound, MedSigLIP, FLAIR, Bio_ClinicalBERT, the Jev open-source implementations, MedKCO, MerMED, DINOv3, C-RADIO, OphthaBERT, Qwen embeddings, MedEmbed, MedGemma, StartLux Decision, and the PyTorch and Hugging Face ecosystems for their data, model weights, and code.

## Author contributions

Zhicheng Du: Conceptualization, Methodology, Software, Validation, Formal analysis, Investigation, Data curation, Visualization, Project administration, Writing—original draft, Writing—review and editing. Zhuo Deng: Writing—review and editing. Lan Ma: Conceptualization, Supervision, Funding acquisition.

## 1 Analytical mechanisms and their limits

These statements are algebraic properties of the evaluated models, not new general learning guarantees. Their role is to distinguish changes that calibration can repair from changes that require a different representation or target.

### Positive binary calibration preserves ranking

Let the original binary margin be *d*(*x*) = *z*_1_(*x*) − *z*_0_(*x*). With a positive temperature and constant class intercepts, the positive probability is *σ*(*d*(*x*)*/T* + *b*_1_ − *b*_0_). Its derivative with respect to *d* is *σ*(1 − *σ*)*/T >* 0. Thus all pairwise orderings and ties are preserved, and AUROC is unchanged on a fixed labeled sample. The intercept can alter the decision threshold and probability losses. In the special case of temperature alone, the zero-margin boundary is also unchanged. This statement does not apply when expert mixture weights change or when input-dependent calibration is used.

### Observed reader loss has a separate ambiguity term

For an empirical response distribution *q* and a model probability vector *p*, expansion gives

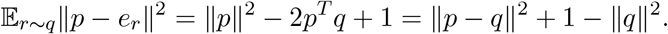

The second term is independent of *p*, so minimizing expected one-reader Brier and distance to the observed response distribution yields the same model ordering. No independence assumption between the observed readers is required for this finite empirical identity. It does not establish that *q* is disease probability or that two responses estimate a population uncertainty distribution accurately.

### Pooling and endpoint conditioning generally do not commute

Let expert *m* assign probabilities *p_m_*(*c*), with fixed mixture weights *w_m_*, and let *S* be the retained class subset. Define *a_m_* = Σ*_c∈S_ p_m_*(*c*) *>* 0. Pooling before conditioning gives

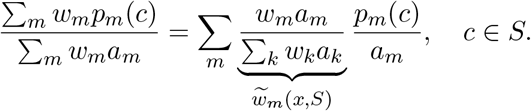

Conditioning first and pooling instead uses the original *w_m_*. They agree when all positive-weight experts have the same retained mass, or in other special cases such as identical conditional predictions. Otherwise endpoint restriction induces input-dependent expert weights. This identity concerns probability pooling before any extra temperature or class-bias transformation. It explains why a binary conditional OCT metric is not equivalent to a full four-class task. Likewise, averaging four-class scan probabilities before subject-level conditioning is not generally the same as averaging already conditioned probabilities.

### What the maximum-cohort objective guarantees

Let *L_k_*(*θ*) be weighted empirical NLL on training cohort *k* and *R*(*θ*) the shared regularizer. The epigraph constraints are *L_k_*(*θ*) + *R*(*θ*) ≤ *t* for every included cohort. A feasible solution bounds these observed regularized cohort losses by its fitted *t*. A global optimum would minimize that empirical bound over the parameter family, but the current mixture-temperature-bias model is nonconvex and SLSQP only supplies a converged feasible local solution. Neither feasibility nor global optimality would bound a new cohort without additional distributional assumptions. In particular, lower worst-training-cohort NLL does not guarantee lower Brier, higher AUROC, or better sensitivity on each external cohort.

### Source preservation requires an explicit endpoint

A dispatch rule that returns the unchanged source prediction when the endpoint key is screening preserves every source probability and any fixed function of it exactly. This is an engineering identity, contingent on a correctly specified endpoint key. It neither infers the clinical endpoint from an image nor guarantees any performance of the other branch. It should not be described as a learned no-regression theorem.

### Quantile-recalibrated operating points cancel monotone recalibration

Let *s* be a binary score and *g* a strictly increasing transformation shared between calibration and evaluation. Select a threshold *t* as an empirical order statistic of the negative source-calibration scores, using the same quantile convention before and after transformation. Order-statistic equivariance gives *t^′^* = *g*(*t*). Therefore *g*(*s*(*x*)) ≥ *t^′^* if and only if *s*(*x*) ≥ *t*, for every evaluation image, regardless of its population. The confusion matrix and referral count are exactly unchanged. A probability-loss gain from monotone recalibration alone cannot improve this operating policy. The identity does not apply when a numerical probability threshold is held fixed, when the ranking changes through expert reweighting, or when a new clinical policy is introduced. Our robust adapter can change ranking, so its measured policy differences are not constrained to be exactly zero.

### A loss-aware query rule depends on expected benefit, not uncertainty alone

Let *p_S_* and *p_F_* denote specialist and fused predictions, *Z* information available without the generalist, and *a*(*Z*) ∈ {0, 1} a query decision. Define Δ(*Z*) = E[*L*(*Y, p_S_*) − *L*(*Y, p_F_*) | *Z*]. With query cost *c*(*Z*) and penalty *λ*, the expected objective is

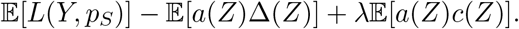

For known conditional gains, pointwise minimization queries where Δ(*Z*) *> λc*(*Z*), with either action allowed at equality. Under a batch cap of *K* equal-cost queries, selecting the largest positive gains is optimal: exchanging a selected lower gain for an unselected higher gain improves the objective. Entropy is not generally sufficient to order these gains. This is a standard decision-theoretic argument, not a new optimality theorem for the fitted gate. Cross-fitted regression supplies an estimate rather than the true conditional gain; distribution shift and estimation error can reverse its empirical advantage. Brier reduction is a probability-quality objective, not a measured clinical utility or treatment benefit.

## 2 Decision-quality exhibits and retained transfer mechanisms

The new plots are descriptive reanalyses of frozen outputs, inspired by the distinction between reliability and resolution in OmniMed-Jev. They are not a direct comparison with its model or training protocol. No test labels enter calibration fitting. Top-label correctness is distinct from disease status. The eight source settings retain positive and adverse calibration metrics.

**Figure 1:**
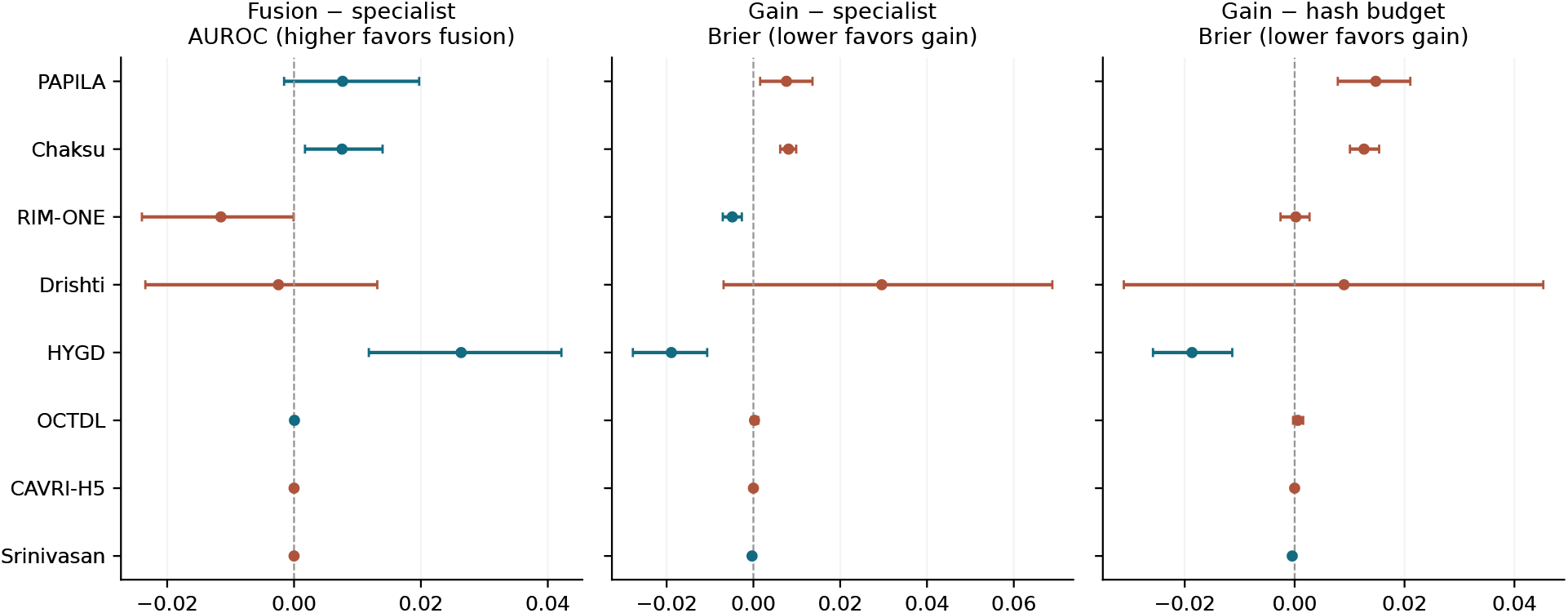
Generalist use has policy-specific benefits and harms. Points are differences of three-seed mean metrics with 500-draw paired group bootstrap intervals for the 2B profile. Fusion is compared with the matched specialist mixture for AUROC (left); gain routing is compared with the same specialist for Brier (middle) and with the same-budget hash policy for Brier (right). Positive AUROC and negative Brier differences favor the named policy. Intervals are exploratory and unadjusted for multiple comparisons.

### Modality-matched transfer helps, but specialist expansion is not uniformly beneficial

A further 144 readout and linear fits test medical image priors and shared representations. On HYGD, MedKCO-CFP with common text attains AUROC 0.6703 versus 0.4423 for MedKCO-OCT; on CAVRI, the direction reverses (0.9021 versus 0.9416). Native MedKCO category scoring reaches HYGD AUROC 0.7417, whereas a source-referral-trained native-text head reaches 0.6715. This comparison changes supervision as well as endpoint wording and cannot identify either factor alone as the cause. Adding dictionary knowledge changes native CAVRI AUROC from 0.7422 to 0.7548, but HYGD from 0.7417 to 0.7400. Clinical descriptions therefore do not provide a universal gain.

The five-expert augmented system retains the original specialists and adds modality-matched MedKCO and MerMED. On OCTDL, fusion reaches AUROC 0.9997 and Brier 0.0248, while on CAVRI it reaches 0.9789 and 0.2955. HYGD instead falls to AUROC 0.7474 and Brier 0.7300, below the original fusion’s 0.7979 and 0.6208. Replacing the original experts altogether further reduces HYGD fusion AUROC to 0.6990. These results identify negative transfer that a larger expert bank does not automatically prevent. The full matrix, paired intervals, knowledge controls, exposure audit, and measured encoder latency are reported in the supplement. No new system is chosen using these evaluation outcomes.

## 3 Unified model complete controls and cohort results

All tables below report three-seed mean metrics rather than an ensemble of seed predictions. Eight policy/control arms are included for every model size and cohort. “Generalist calibrated” receives the same external training cohorts as fusion; its results differ from the separately reported zero-shot StartLux results. Calls are logical selected-image fractions before removal of exactly zero-weight components. They are conservative computation proxies, not timings or patient referrals. LOCO is used for the six original cohorts, including Chaksu, whose suspicion labels never enter clinical adaptation. HYGD and Srinivasan labels never enter the mixture or gate optimizers, but prior inspection makes these exploratory analyses. Cross-fitting excludes each target connected group from the calibration fits generating its gain target, not from the separately pretrained source representations.

**Figure 2:**
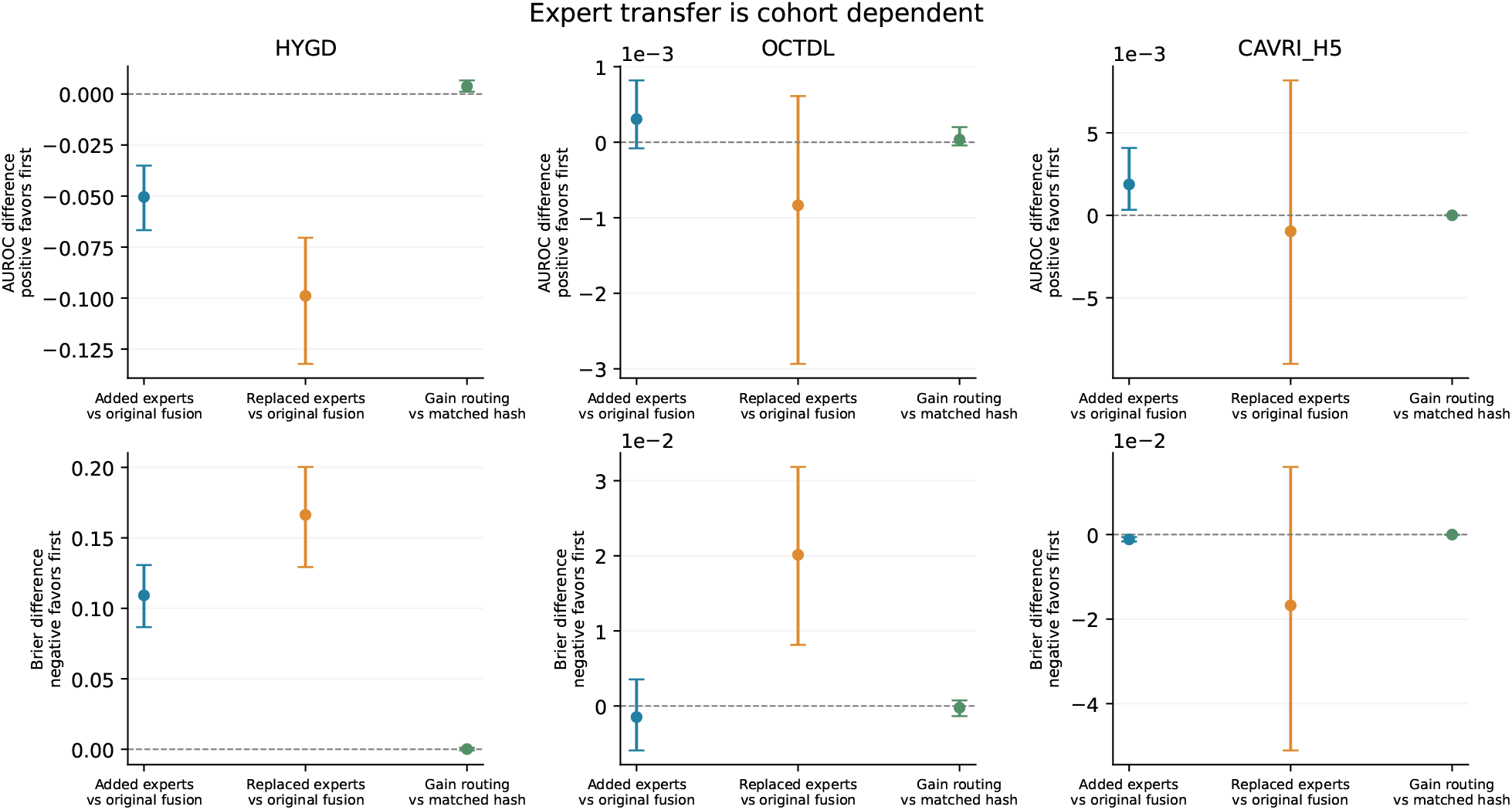
Paired effects of specialist-bank expansion and selective querying. Points are differences of three-seed mean metrics; bars are 500-draw, unadjusted connected-group bootstrap intervals. CAVRI gains coexist with HYGD harm, while OCTDL differences are small. Expert-bank changes remain exploratory; exposure flags and cohort sizes are in the supplement.

**Figure 3:**
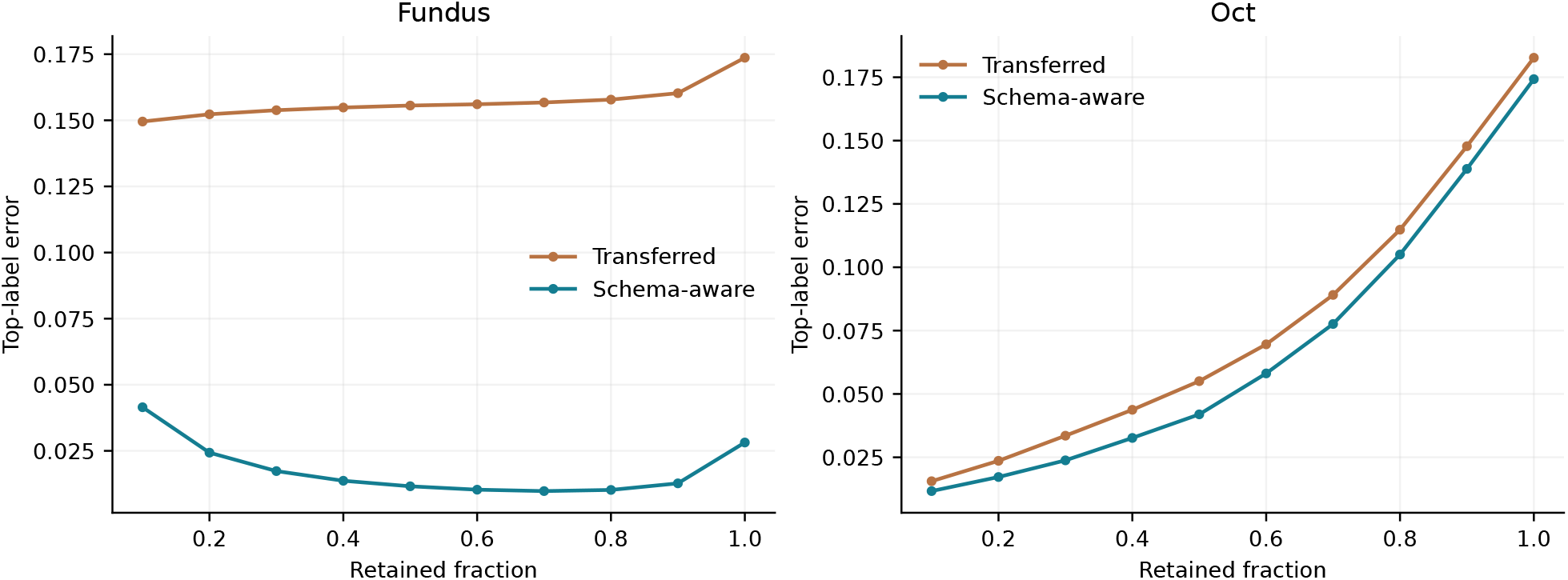
Descriptive risk–coverage curves, averaging the four image/text settings and three seeds/descriptions within each modality. Curves are model-confidence selection, not measured clinical referral outcomes.

**Table 1:**
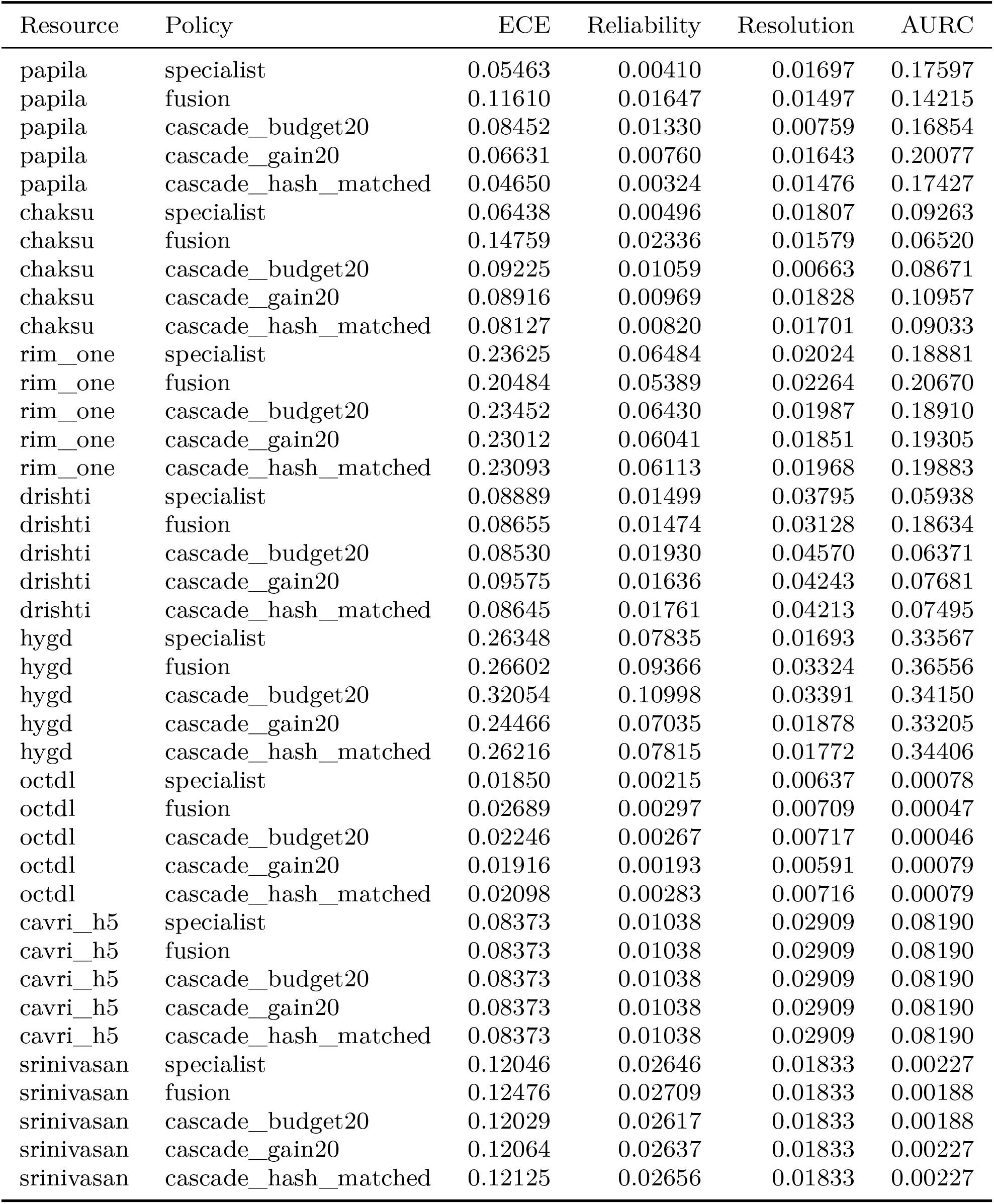
External top-label decision quality, three-seed means; all eight cohorts and five evaluated policy arms retained.

| Resource | Policy | ECE | Reliability | Resolution | AURC |
| --- | --- | --- | --- | --- | --- |
| papila | specialist | 0.05463 | 0.00410 | 0.01697 | 0.17597 |
| papila | fusion | 0.11610 | 0.01647 | 0.01497 | 0.14215 |
| papila | cascade_budget20 | 0.08452 | 0.01330 | 0.00759 | 0.16854 |
| papila | cascade_gain20 | 0.06631 | 0.00760 | 0.01643 | 0.20077 |
| papila | cascade_hash_matched | 0.04650 | 0.00324 | 0.01476 | 0.17427 |
| chaksu | specialist | 0.06438 | 0.00496 | 0.01807 | 0.09263 |
| chaksu | fusion | 0.14759 | 0.02336 | 0.01579 | 0.06520 |
| chaksu | cascade_budget20 | 0.09225 | 0.01059 | 0.00663 | 0.08671 |
| chaksu | cascade_gain20 | 0.08916 | 0.00969 | 0.01828 | 0.10957 |
| chaksu | cascade_hash_matched | 0.08127 | 0.00820 | 0.01701 | 0.09033 |
| rim_one | specialist | 0.23625 | 0.06484 | 0.02024 | 0.18881 |
| rim_one | fusion | 0.20484 | 0.05389 | 0.02264 | 0.20670 |
| rim_one | cascade_budget20 | 0.23452 | 0.06430 | 0.01987 | 0.18910 |
| rim_one | cascade_gain20 | 0.23012 | 0.06041 | 0.01851 | 0.19305 |
| rim_one | cascade_hash_matched | 0.23093 | 0.06113 | 0.01968 | 0.19883 |
| drishti | specialist | 0.08889 | 0.01499 | 0.03795 | 0.05938 |
| drishti | fusion | 0.08655 | 0.01474 | 0.03128 | 0.18634 |
| drishti | cascade_budget20 | 0.08530 | 0.01930 | 0.04570 | 0.06371 |
| drishti | cascade_gain20 | 0.09575 | 0.01636 | 0.04243 | 0.07681 |
| drishti | cascade_hash_matched | 0.08645 | 0.01761 | 0.04213 | 0.07495 |
| hygd | specialist | 0.26348 | 0.07835 | 0.01693 | 0.33567 |
| hygd | fusion | 0.26602 | 0.09366 | 0.03324 | 0.36556 |
| hygd | cascade_budget20 | 0.32054 | 0.10998 | 0.03391 | 0.34150 |
| hygd | cascade_gain20 | 0.24466 | 0.07035 | 0.01878 | 0.33205 |
| hygd | cascade_hash_matched | 0.26216 | 0.07815 | 0.01772 | 0.34406 |
| octdl | specialist | 0.01850 | 0.00215 | 0.00637 | 0.00078 |
| octdl | fusion | 0.02689 | 0.00297 | 0.00709 | 0.00047 |
| octdl | cascade_budget20 | 0.02246 | 0.00267 | 0.00717 | 0.00046 |
| octdl | cascade_gain20 | 0.01916 | 0.00193 | 0.00591 | 0.00079 |
| octdl | cascade_hash_matched | 0.02098 | 0.00283 | 0.00716 | 0.00079 |
| cavri_h5 | specialist | 0.08373 | 0.01038 | 0.02909 | 0.08190 |
| cavri_h5 | fusion | 0.08373 | 0.01038 | 0.02909 | 0.08190 |
| cavri_h5 | cascade_budget20 | 0.08373 | 0.01038 | 0.02909 | 0.08190 |
| cavri_h5 | cascade_gain20 | 0.08373 | 0.01038 | 0.02909 | 0.08190 |
| cavri_h5 | cascade_hash_matched | 0.08373 | 0.01038 | 0.02909 | 0.08190 |
| srinivasan | specialist | 0.12046 | 0.02646 | 0.01833 | 0.00227 |
| srinivasan | fusion | 0.12476 | 0.02709 | 0.01833 | 0.00188 |
| srinivasan | cascade_budget20 | 0.12029 | 0.02617 | 0.01833 | 0.00188 |
| srinivasan | cascade_gain20 | 0.12064 | 0.02637 | 0.01833 | 0.00227 |
| srinivasan | cascade_hash_matched | 0.12125 | 0.02656 | 0.01833 | 0.00227 |

**Figure 4:**
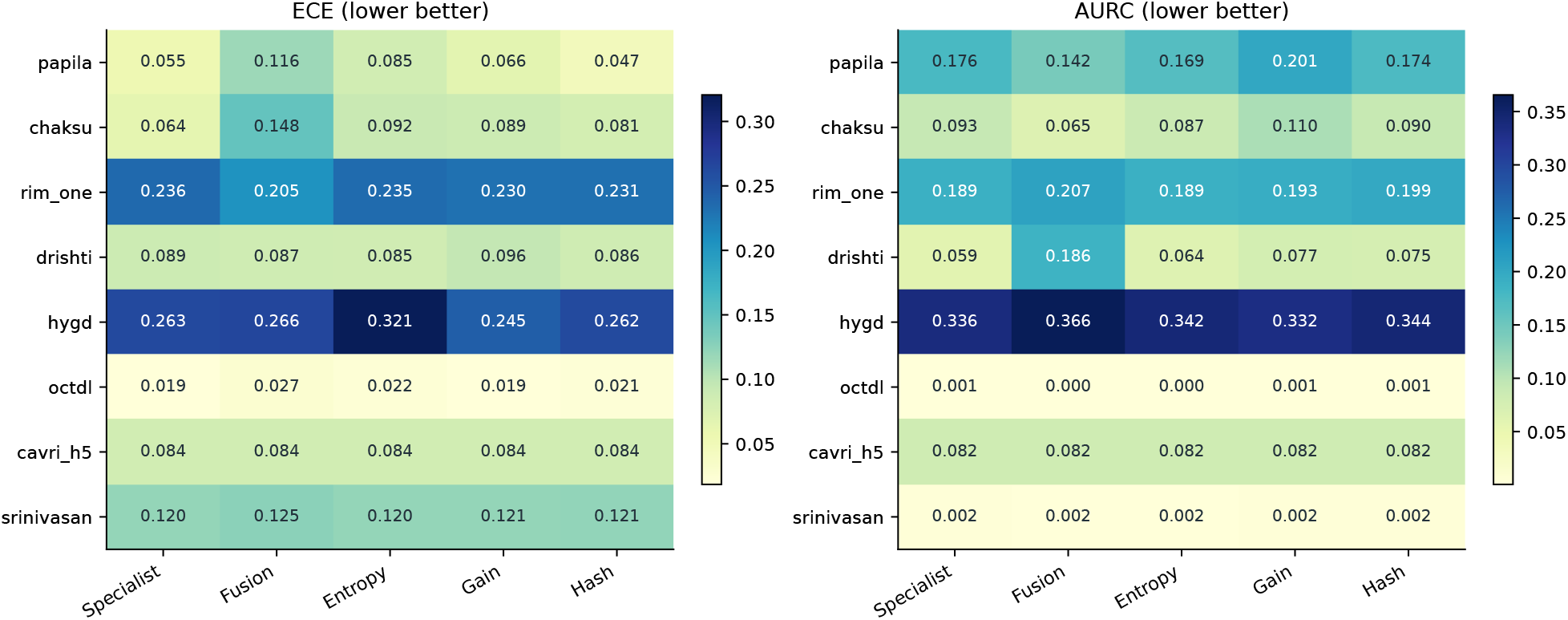
Complete external ECE and AURC matrix for the original 2B profile. The color scale does not imply statistical significance.

**Table 2:**
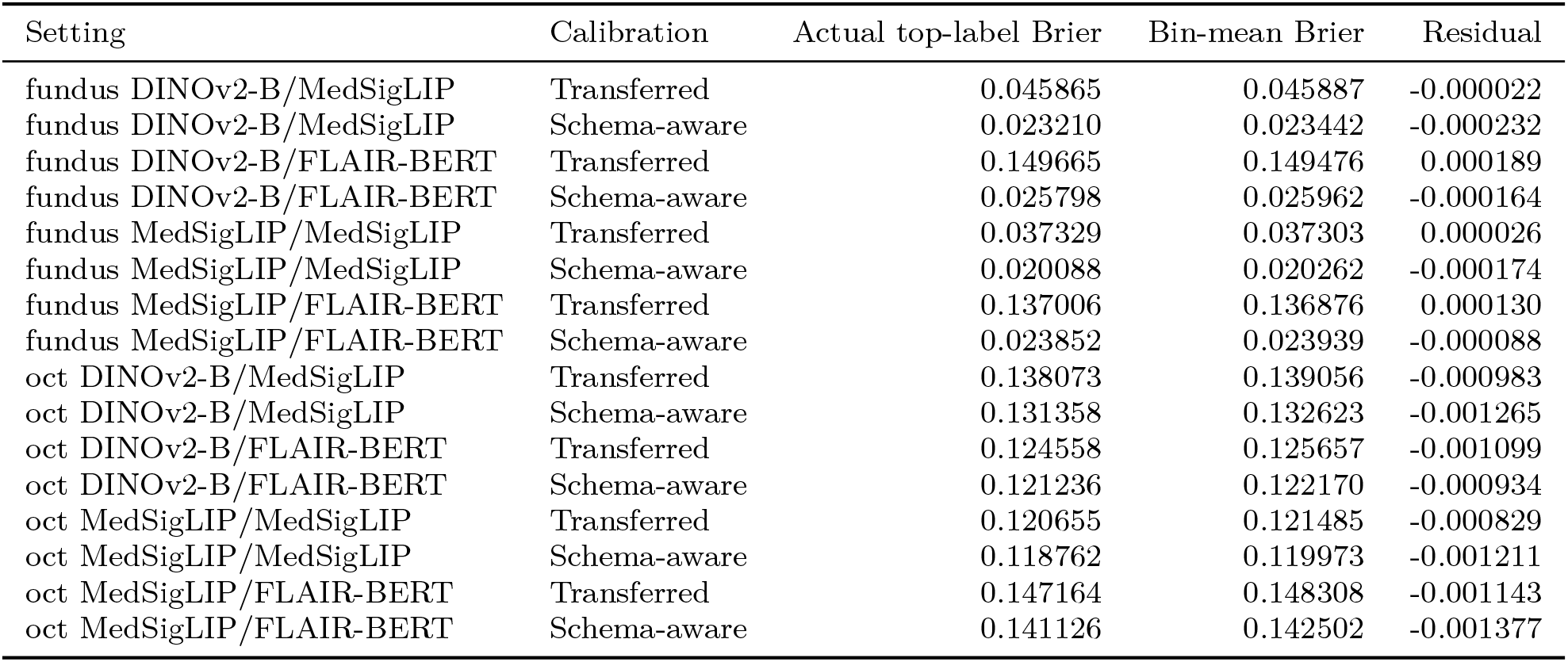
Binning audit. Bin-mean Brier equals reliability minus resolution plus uncertainty to numerical precision. The reported residual prevents treating a binned approximation as an exact decomposition of unbinned confidence.

| Setting | Calibration | Actual top-label Brier | Bin-mean Brier | Residual |
| --- | --- | --- | --- | --- |
| fundus DINOv2-B/MedSigLIP | Transferred | 0.045865 | 0.045887 | -0.000022 |
| fundus DINOv2-B/MedSigLIP | Schema-aware | 0.023210 | 0.023442 | -0.000232 |
| fundus DINOv2-B/FLAIR-BERT | Transferred | 0.149665 | 0.149476 | 0.000189 |
| fundus DINOv2-B/FLAIR-BERT | Schema-aware | 0.025798 | 0.025962 | -0.000164 |
| fundus MedSigLIP/MedSigLIP | Transferred | 0.037329 | 0.037303 | 0.000026 |
| fundus MedSigLIP/MedSigLIP | Schema-aware | 0.020088 | 0.020262 | -0.000174 |
| fundus MedSigLIP/FLAIR-BERT | Transferred | 0.137006 | 0.136876 | 0.000130 |
| fundus MedSigLIP/FLAIR-BERT | Schema-aware | 0.023852 | 0.023939 | -0.000088 |
| oct DINOv2-B/MedSigLIP | Transferred | 0.138073 | 0.139056 | -0.000983 |
| oct DINOv2-B/MedSigLIP | Schema-aware | 0.131358 | 0.132623 | -0.001265 |
| oct DINOv2-B/FLAIR-BERT | Transferred | 0.124558 | 0.125657 | -0.001099 |
| oct DINOv2-B/FLAIR-BERT | Schema-aware | 0.121236 | 0.122170 | -0.000934 |
| oct MedSigLIP/MedSigLIP | Transferred | 0.120655 | 0.121485 | -0.000829 |
| oct MedSigLIP/MedSigLIP | Schema-aware | 0.118762 | 0.119973 | -0.001211 |
| oct MedSigLIP/FLAIR-BERT | Transferred | 0.147164 | 0.148308 | -0.001143 |
| oct MedSigLIP/FLAIR-BERT | Schema-aware | 0.141126 | 0.142502 | -0.001377 |

### 3.1 0.8B generalist profile

| Resource | Policy | AUROC | Brier | Accuracy | Calls (%) |
| --- | --- | --- | --- | --- | --- |
| PAPILA | Specialist | 0.8240 | 0.3795 | 0.7167 | 0.00 |
| PAPILA | Generalist calibrated | 0.4949 | 0.5163 | 0.3976 | 100.00 |
| PAPILA | Uniform | 0.7409 | 0.2818 | 0.8024 | 100.00 |
| PAPILA | Fusion | 0.8123 | 0.3591 | 0.7706 | 100.00 |
| PAPILA | Entropy threshold | 0.8215 | 0.3561 | 0.7706 | 36.41 |
| PAPILA | Entropy budget | 0.8232 | 0.3652 | 0.7683 | 19.88 |
| PAPILA | Gain budget | 0.8276 | 0.3765 | 0.7262 | 19.88 |
| PAPILA | Hash matched | 0.8202 | 0.3744 | 0.7270 | 19.88 |
| Chaksu | Specialist | 0.8143 | 0.2842 | 0.8064 | 0.00 |
| Chaksu | Generalist calibrated | 0.6265 | 0.3483 | 0.8617 | 100.00 |
| Chaksu | Uniform | 0.7817 | 0.2198 | 0.8615 | 100.00 |
| Chaksu | Fusion | 0.8070 | 0.2673 | 0.8505 | 100.00 |
| Chaksu | Entropy threshold | 0.8100 | 0.2606 | 0.8505 | 32.22 |
| Chaksu | Entropy budget | 0.8110 | 0.2671 | 0.8460 | 20.00 |
| Chaksu | Gain budget | 0.8098 | 0.2913 | 0.8064 | 20.00 |
| Chaksu | Hash matched | 0.8113 | 0.2803 | 0.8161 | 20.00 |
| RIM-ONE | Specialist | 0.7318 | 0.5290 | 0.6674 | 0.00 |
| RIM-ONE | Generalist calibrated | 0.3824 | 0.4885 | 0.6082 | 100.00 |
| RIM-ONE | Uniform | 0.5013 | 0.5506 | 0.6495 | 100.00 |
| RIM-ONE | Fusion | 0.7170 | 0.5148 | 0.6674 | 100.00 |
| RIM-ONE | Entropy threshold | 0.7318 | 0.5299 | 0.6674 | 7.08 |
| RIM-ONE | Entropy budget | 0.7316 | 0.5297 | 0.6674 | 20.00 |
| RIM-ONE | Gain budget | 0.7308 | 0.5266 | 0.6680 | 11.00 |
| RIM-ONE | Hash matched | 0.7186 | 0.5265 | 0.6674 | 11.00 |
| Drishti | Specialist | 0.9046 | 0.2517 | 0.8185 | 0.00 |
| Drishti | Generalist calibrated | 0.7055 | 0.7215 | 0.3069 | 100.00 |
| Drishti | Uniform | 0.8857 | 0.7415 | 0.3828 | 100.00 |
| Drishti | Fusion | 0.9020 | 0.3812 | 0.6865 | 100.00 |
| Drishti | Entropy threshold | 0.9094 | 0.2740 | 0.7921 | 20.46 |
| Drishti | Entropy budget | 0.9118 | 0.2726 | 0.7921 | 19.80 |
| Drishti | Gain budget | 0.9127 | 0.2757 | 0.7954 | 19.80 |
| Drishti | Hash matched | 0.8942 | 0.2687 | 0.8020 | 19.80 |
| HYGD | Specialist | 0.7715 | 0.6183 | 0.5387 | 0.00 |
| HYGD | Generalist calibrated | 0.7118 | 0.6629 | 0.2687 | 100.00 |
| HYGD | Uniform | 0.8075 | 1.0468 | 0.2763 | 100.00 |
| HYGD | Fusion | 0.7927 | 0.6125 | 0.4903 | 100.00 |
| HYGD | Entropy threshold | 0.7739 | 0.6483 | 0.4907 | 21.82 |
| HYGD | Entropy budget | 0.7735 | 0.6469 | 0.4930 | 19.95 |
| HYGD | Gain budget | 0.7642 | 0.6080 | 0.5378 | 19.95 |
| HYGD | Hash matched | 0.7687 | 0.6158 | 0.5305 | 19.95 |
| OCTDL | Specialist | 0.9993 | 0.0280 | 0.9817 | 0.00 |
| OCTDL | Generalist calibrated | 0.9891 | 0.1231 | 0.9430 | 100.00 |
| OCTDL | Uniform | 0.9993 | 0.0395 | 0.9859 | 100.00 |
| OCTDL | Fusion | 0.9993 | 0.0280 | 0.9824 | 100.00 |
| OCTDL | Entropy threshold | 0.9994 | 0.0274 | 0.9824 | 23.50 |
| OCTDL | Entropy budget | 0.9994 | 0.0274 | 0.9824 | 19.96 |
| OCTDL | Gain budget | 0.9993 | 0.0280 | 0.9817 | 19.96 |
| OCTDL | Hash matched | 0.9993 | 0.0281 | 0.9817 | 19.96 |
| CAVRI-H5 | Specialist | 0.9770 | 0.2967 | 0.8003 | 0.00 |
| CAVRI-H5 | Generalist calibrated | 0.9032 | 0.5511 | 0.5759 | 100.00 |
| CAVRI-H5 | Uniform | 0.9781 | 0.3035 | 0.7940 | 100.00 |
| CAVRI-H5 | Fusion | 0.9770 | 0.2967 | 0.8003 | 100.00 |
| CAVRI-H5 | Entropy threshold | 0.9770 | 0.2967 | 0.8003 | 18.01 |
| CAVRI-H5 | Entropy budget | 0.9770 | 0.2967 | 0.8003 | 19.87 |
| CAVRI-H5 | Gain budget | 0.9770 | 0.2967 | 0.8003 | 19.87 |
| CAVRI-H5 | Hash matched | 0.9770 | 0.2967 | 0.8003 | 19.87 |
| Srinivasan | Specialist | 1.0000 | 0.0807 | 0.9667 | 0.00 |
| Srinivasan | Generalist calibrated | 1.0000 | 0.2840 | 0.9000 | 100.00 |
| Srinivasan | Uniform | 1.0000 | 0.1337 | 0.9444 | 100.00 |
| Srinivasan | Fusion | 1.0000 | 0.0805 | 0.9667 | 100.00 |
| Srinivasan | Entropy threshold | 1.0000 | 0.0802 | 0.9667 | 31.25 |
| Srinivasan | Entropy budget | 1.0000 | 0.0803 | 0.9667 | 19.99 |
| Srinivasan | Gain budget | 1.0000 | 0.0805 | 0.9667 | 19.99 |
| Srinivasan | Hash matched | 1.0000 | 0.0806 | 0.9667 | 19.99 |

### 3.2 2B generalist profile

| Resource | Policy | AUROC | Brier | Accuracy | Calls (%) |
| --- | --- | --- | --- | --- | --- |
| PAPILA | Specialist | 0.8240 | 0.3795 | 0.7167 | 0.00 |
| PAPILA | Generalist calibrated | 0.7624 | 0.4283 | 0.7000 | 100.00 |
| PAPILA | Uniform | 0.8267 | 0.2610 | 0.8230 | 100.00 |
| PAPILA | Fusion | 0.8316 | 0.3464 | 0.7794 | 100.00 |
| PAPILA | Entropy threshold | 0.8266 | 0.3510 | 0.7794 | 36.41 |
| PAPILA | Entropy budget | 0.8258 | 0.3622 | 0.7730 | 19.88 |
| PAPILA | Gain budget | 0.8261 | 0.3871 | 0.7175 | 19.88 |
| PAPILA | Hash matched | 0.8214 | 0.3723 | 0.7278 | 19.88 |
| Chaksu | Specialist | 0.8143 | 0.2842 | 0.8064 | 0.00 |
| Chaksu | Generalist calibrated | 0.7731 | 0.3040 | 0.8572 | 100.00 |
| Chaksu | Uniform | 0.8223 | 0.2158 | 0.8652 | 100.00 |
| Chaksu | Fusion | 0.8219 | 0.2641 | 0.8528 | 100.00 |
| Chaksu | Entropy threshold | 0.8148 | 0.2585 | 0.8523 | 32.22 |
| Chaksu | Entropy budget | 0.8145 | 0.2656 | 0.8475 | 20.00 |
| Chaksu | Gain budget | 0.8148 | 0.2923 | 0.8072 | 20.00 |
| Chaksu | Hash matched | 0.8132 | 0.2796 | 0.8169 | 20.00 |
| RIM-ONE | Specialist | 0.7318 | 0.5290 | 0.6674 | 0.00 |
| RIM-ONE | Generalist calibrated | 0.5824 | 0.4982 | 0.5918 | 100.00 |
| RIM-ONE | Uniform | 0.6167 | 0.4843 | 0.6515 | 100.00 |
| RIM-ONE | Fusion | 0.7203 | 0.5041 | 0.6674 | 100.00 |
| RIM-ONE | Entropy threshold | 0.7318 | 0.5301 | 0.6674 | 7.08 |
| RIM-ONE | Entropy budget | 0.7313 | 0.5288 | 0.6674 | 20.00 |
| RIM-ONE | Gain budget | 0.7193 | 0.5242 | 0.6674 | 13.68 |
| RIM-ONE | Hash matched | 0.7142 | 0.5239 | 0.6674 | 13.68 |
| Drishti | Specialist | 0.9046 | 0.2517 | 0.8185 | 0.00 |
| Drishti | Generalist calibrated | 0.8387 | 0.6347 | 0.3564 | 100.00 |
| Drishti | Uniform | 0.8922 | 0.6069 | 0.4653 | 100.00 |
| Drishti | Fusion | 0.9022 | 0.4091 | 0.6700 | 100.00 |
| Drishti | Entropy threshold | 0.9048 | 0.2789 | 0.7921 | 20.46 |
| Drishti | Entropy budget | 0.9075 | 0.2768 | 0.7921 | 19.80 |
| Drishti | Gain budget | 0.9091 | 0.2813 | 0.7888 | 19.80 |
| Drishti | Hash matched | 0.8943 | 0.2723 | 0.7987 | 19.80 |
| HYGD | Specialist | 0.7715 | 0.6183 | 0.5387 | 0.00 |
| HYGD | Generalist calibrated | 0.7409 | 0.7204 | 0.3039 | 100.00 |
| HYGD | Uniform | 0.7928 | 0.9335 | 0.2917 | 100.00 |
| HYGD | Fusion | 0.7979 | 0.6208 | 0.4844 | 100.00 |
| HYGD | Entropy threshold | 0.7753 | 0.6519 | 0.4876 | 21.82 |
| HYGD | Entropy budget | 0.7750 | 0.6501 | 0.4885 | 19.95 |
| HYGD | Gain budget | 0.7670 | 0.5994 | 0.5400 | 19.95 |
| HYGD | Hash matched | 0.7707 | 0.6181 | 0.5296 | 19.95 |
| OCTDL | Specialist | 0.9993 | 0.0280 | 0.9817 | 0.00 |
| OCTDL | Generalist calibrated | 0.9929 | 0.0566 | 0.9641 | 100.00 |
| OCTDL | Uniform | 0.9994 | 0.0265 | 0.9859 | 100.00 |
| OCTDL | Fusion | 0.9994 | 0.0263 | 0.9824 | 100.00 |
| OCTDL | Entropy threshold | 0.9994 | 0.0255 | 0.9824 | 23.50 |
| OCTDL | Entropy budget | 0.9994 | 0.0255 | 0.9824 | 19.96 |
| OCTDL | Gain budget | 0.9993 | 0.0283 | 0.9817 | 19.96 |
| OCTDL | Hash matched | 0.9992 | 0.0277 | 0.9817 | 19.96 |
| CAVRI-H5 | Specialist | 0.9770 | 0.2967 | 0.8003 | 0.00 |
| CAVRI-H5 | Generalist calibrated | 0.9781 | 0.3622 | 0.7405 | 100.00 |
| CAVRI-H5 | Uniform | 0.9867 | 0.2960 | 0.7947 | 100.00 |
| CAVRI-H5 | Fusion | 0.9770 | 0.2967 | 0.8003 | 100.00 |
| CAVRI-H5 | Entropy threshold | 0.9770 | 0.2967 | 0.8003 | 18.01 |
| CAVRI-H5 | Entropy budget | 0.9770 | 0.2967 | 0.8003 | 19.87 |
| CAVRI-H5 | Gain budget | 0.9770 | 0.2967 | 0.8003 | 19.87 |
| CAVRI-H5 | Hash matched | 0.9770 | 0.2967 | 0.8003 | 19.87 |
| Srinivasan | Specialist | 1.0000 | 0.0807 | 0.9667 | 0.00 |
| Srinivasan | Generalist calibrated | 1.0000 | 0.1429 | 0.9000 | 100.00 |
| Srinivasan | Uniform | 1.0000 | 0.1305 | 0.9000 | 100.00 |
| Srinivasan | Fusion | 1.0000 | 0.0814 | 0.9667 | 100.00 |
| Srinivasan | Entropy threshold | 1.0000 | 0.0796 | 0.9667 | 31.25 |
| Srinivasan | Entropy budget | 1.0000 | 0.0798 | 0.9667 | 19.99 |
| Srinivasan | Gain budget | 1.0000 | 0.0803 | 0.9667 | 19.99 |
| Srinivasan | Hash matched | 1.0000 | 0.0808 | 0.9667 | 19.99 |

### 3.3 4B generalist profile

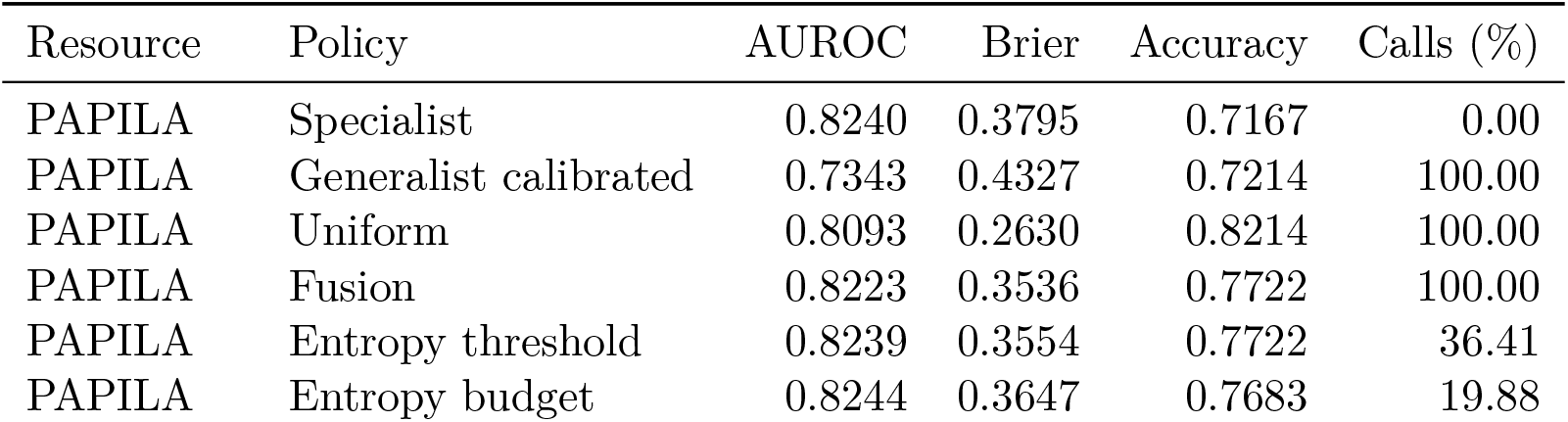
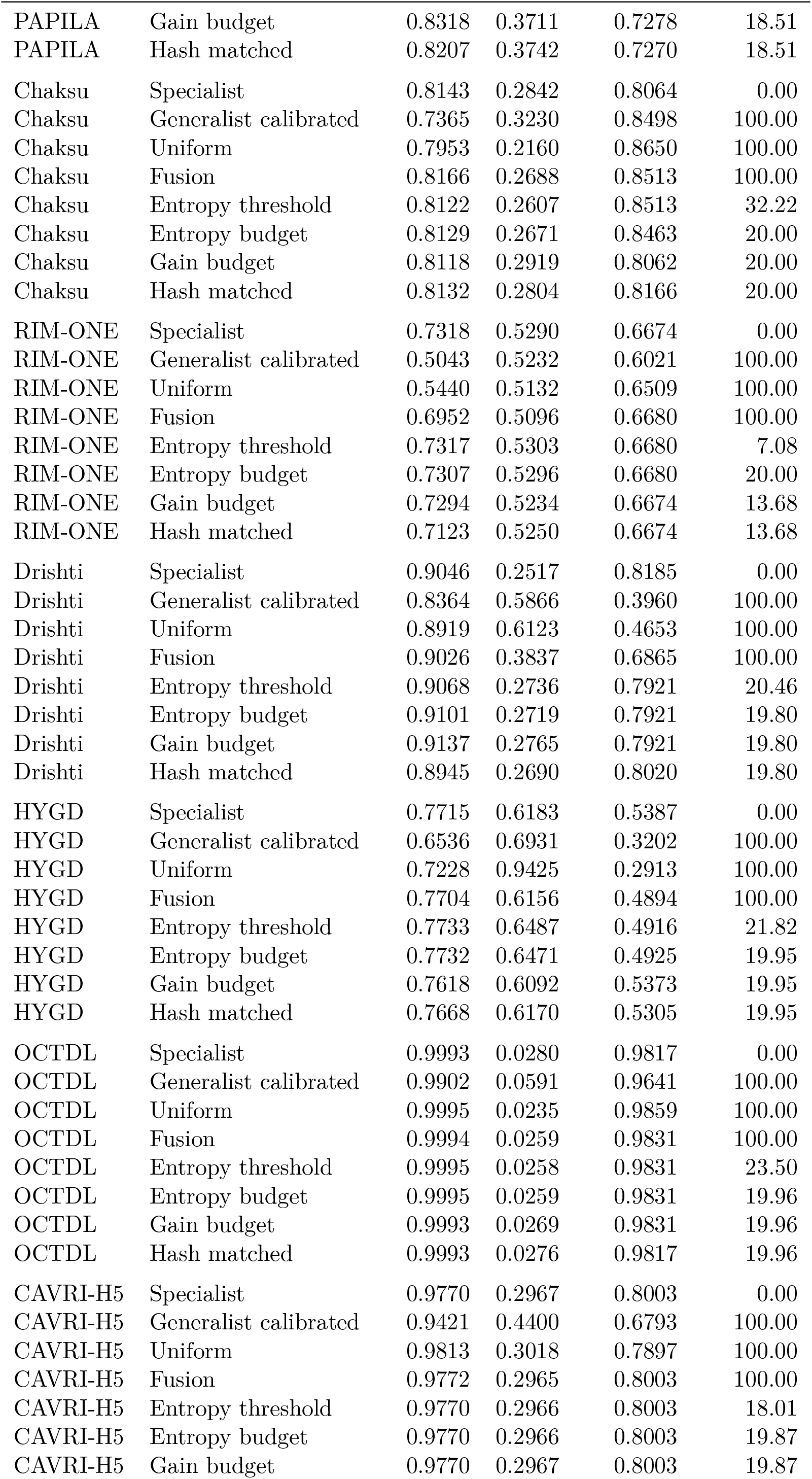
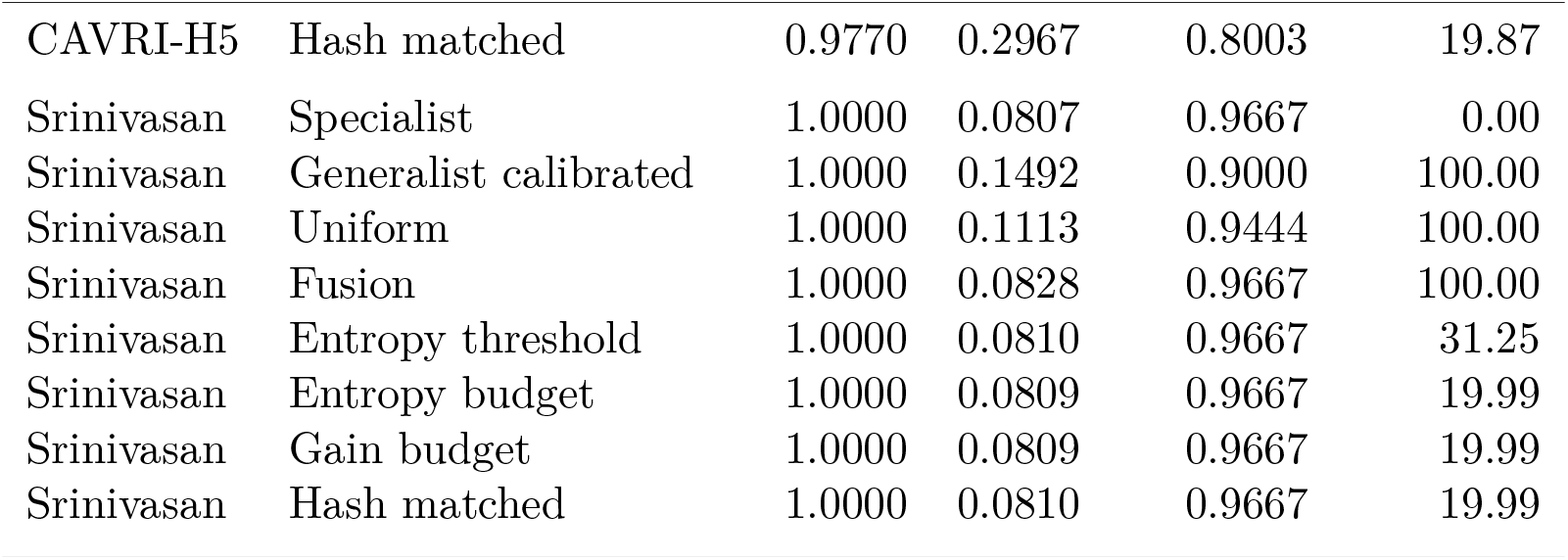

| Resource | Policy | AUROC | Brier | Accuracy | Calls (%) |
| --- | --- | --- | --- | --- | --- |
| PAPILA | Specialist | 0.8240 | 0.3795 | 0.7167 | 0.00 |
| PAPILA | Generalist calibrated | 0.7343 | 0.4327 | 0.7214 | 100.00 |
| PAPILA | Uniform | 0.8093 | 0.2630 | 0.8214 | 100.00 |
| PAPILA | Fusion | 0.8223 | 0.3536 | 0.7722 | 100.00 |
| PAPILA | Entropy threshold | 0.8239 | 0.3554 | 0.7722 | 36.41 |
| PAPILA | Entropy budget | 0.8244 | 0.3647 | 0.7683 | 19.88 |
| PAPILA | Gain budget | 0.8318 | 0.3711 | 0.7278 | 18.51 |
| PAPILA | Hash matched | 0.8207 | 0.3742 | 0.7270 | 18.51 |
| Chaksu | Specialist | 0.8143 | 0.2842 | 0.8064 | 0.00 |
| Chaksu | Generalist calibrated | 0.7365 | 0.3230 | 0.8498 | 100.00 |
| Chaksu | Uniform | 0.7953 | 0.2160 | 0.8650 | 100.00 |
| Chaksu | Fusion | 0.8166 | 0.2688 | 0.8513 | 100.00 |
| Chaksu | Entropy threshold | 0.8122 | 0.2607 | 0.8513 | 32.22 |
| Chaksu | Entropy budget | 0.8129 | 0.2671 | 0.8463 | 20.00 |
| Chaksu | Gain budget | 0.8118 | 0.2919 | 0.8062 | 20.00 |
| Chaksu | Hash matched | 0.8132 | 0.2804 | 0.8166 | 20.00 |
| RIM-ONE | Specialist | 0.7318 | 0.5290 | 0.6674 | 0.00 |
| RIM-ONE | Generalist calibrated | 0.5043 | 0.5232 | 0.6021 | 100.00 |
| RIM-ONE | Uniform | 0.5440 | 0.5132 | 0.6509 | 100.00 |
| RIM-ONE | Fusion | 0.6952 | 0.5096 | 0.6680 | 100.00 |
| RIM-ONE | Entropy threshold | 0.7317 | 0.5303 | 0.6680 | 7.08 |
| RIM-ONE | Entropy budget | 0.7307 | 0.5296 | 0.6680 | 20.00 |
| RIM-ONE | Gain budget | 0.7294 | 0.5234 | 0.6674 | 13.68 |
| RIM-ONE | Hash matched | 0.7123 | 0.5250 | 0.6674 | 13.68 |
| Drishti | Specialist | 0.9046 | 0.2517 | 0.8185 | 0.00 |
| Drishti | Generalist calibrated | 0.8364 | 0.5866 | 0.3960 | 100.00 |
| Drishti | Uniform | 0.8919 | 0.6123 | 0.4653 | 100.00 |
| Drishti | Fusion | 0.9026 | 0.3837 | 0.6865 | 100.00 |
| Drishti | Entropy threshold | 0.9068 | 0.2736 | 0.7921 | 20.46 |
| Drishti | Entropy budget | 0.9101 | 0.2719 | 0.7921 | 19.80 |
| Drishti | Gain budget | 0.9137 | 0.2765 | 0.7921 | 19.80 |
| Drishti | Hash matched | 0.8945 | 0.2690 | 0.8020 | 19.80 |
| HYGD | Specialist | 0.7715 | 0.6183 | 0.5387 | 0.00 |
| HYGD | Generalist calibrated | 0.6536 | 0.6931 | 0.3202 | 100.00 |
| HYGD | Uniform | 0.7228 | 0.9425 | 0.2913 | 100.00 |
| HYGD | Fusion | 0.7704 | 0.6156 | 0.4894 | 100.00 |
| HYGD | Entropy threshold | 0.7733 | 0.6487 | 0.4916 | 21.82 |
| HYGD | Entropy budget | 0.7732 | 0.6471 | 0.4925 | 19.95 |
| HYGD | Gain budget | 0.7618 | 0.6092 | 0.5373 | 19.95 |
| HYGD | Hash matched | 0.7668 | 0.6170 | 0.5305 | 19.95 |
| OCTDL | Specialist | 0.9993 | 0.0280 | 0.9817 | 0.00 |
| OCTDL | Generalist calibrated | 0.9902 | 0.0591 | 0.9641 | 100.00 |
| OCTDL | Uniform | 0.9995 | 0.0235 | 0.9859 | 100.00 |
| OCTDL | Fusion | 0.9994 | 0.0259 | 0.9831 | 100.00 |
| OCTDL | Entropy threshold | 0.9995 | 0.0258 | 0.9831 | 23.50 |
| OCTDL | Entropy budget | 0.9995 | 0.0259 | 0.9831 | 19.96 |
| OCTDL | Gain budget | 0.9993 | 0.0269 | 0.9831 | 19.96 |
| OCTDL | Hash matched | 0.9993 | 0.0276 | 0.9817 | 19.96 |
| CAVRI-H5 | Specialist | 0.9770 | 0.2967 | 0.8003 | 0.00 |
| CAVRI-H5 | Generalist calibrated | 0.9421 | 0.4400 | 0.6793 | 100.00 |
| CAVRI-H5 | Uniform | 0.9813 | 0.3018 | 0.7897 | 100.00 |
| CAVRI-H5 | Fusion | 0.9772 | 0.2965 | 0.8003 | 100.00 |
| CAVRI-H5 | Entropy threshold | 0.9770 | 0.2966 | 0.8003 | 18.01 |
| CAVRI-H5 | Entropy budget | 0.9770 | 0.2966 | 0.8003 | 19.87 |
| CAVRI-H5 | Gain budget | 0.9770 | 0.2967 | 0.8003 | 19.87 |
| CAVRI-H5 | Hash matched | 0.9770 | 0.2967 | 0.8003 | 19.87 |
| Srinivasan | Specialist | 1.0000 | 0.0807 | 0.9667 | 0.00 |
| Srinivasan | Generalist calibrated | 1.0000 | 0.1492 | 0.9000 | 100.00 |
| Srinivasan | Uniform | 1.0000 | 0.1113 | 0.9444 | 100.00 |
| Srinivasan | Fusion | 1.0000 | 0.0828 | 0.9667 | 100.00 |
| Srinivasan | Entropy threshold | 1.0000 | 0.0810 | 0.9667 | 31.25 |
| Srinivasan | Entropy budget | 1.0000 | 0.0809 | 0.9667 | 19.99 |
| Srinivasan | Gain budget | 1.0000 | 0.0809 | 0.9667 | 19.99 |
| Srinivasan | Hash matched | 1.0000 | 0.0810 | 0.9667 | 19.99 |

### 3.4 Primary paired uncertainty and budget controls

These are 500-draw paired connected-group percentile intervals, conditional on fitted states and shared across seeds. Srinivasan is resampled at subject level after averaging original four-class probabilities and then restricting to DME/normal. Other endpoints retain the grouping defined for each resource. No multiplicity correction or adjustment for repeated inspection is applied. Complete 96 primary and 30 control contrasts, including accuracy, are supplied in the aggregate JSON files.

**Full fusion versus specialist.**

| Resource | AUROC difference [95% interval] | Brier difference [95% interval] |
| --- | --- | --- |
| PAPILA | +0.0077 [−0.0016, +0.0197] | −0.0331 [−0.0489, −0.0155] |
| Chaksu | +0.0076 [+0.0017, +0.0140] | −0.0201 [−0.0274, −0.0123] |
| RIM-ONE | −0.0115 [−0.0240, −0.0001] | −0.0249 [−0.0306, −0.0198] |
| Drishti | −0.0025 [−0.0234, +0.0131] | +0.1573 [+0.1002, +0.2289] |
| HYGD | +0.0263 [+0.0118, +0.0421] | +0.0024 [−0.0201, +0.0254] |
| OCTDL | +0.0001 [−0.0002, +0.0004] | −0.0018 [−0.0051, +0.0011] |
| CAVRI-H5 | +0.0000 [+0.0000, +0.0000] | −0.0000 [−0.0000, +0.0000] |
| Srinivasan | +0.0000 [+0.0000, +0.0000] | +0.0007 [−0.0011, +0.0021] |

**Gain routing versus specialist.**

| Resource | AUROC difference [95% interval] | Brier difference [95% interval] |
| --- | --- | --- |
| PAPILA | +0.0021 [−0.0077, +0.0134] | +0.0076 [+0.0015, +0.0136] |
| Chaksu | +0.0005 [−0.0049, +0.0068] | +0.0081 [+0.0062, +0.0099] |
| RIM-ONE | −0.0125 [−0.0260, +0.0012] | −0.0049 [−0.0071, −0.0027] |
| Drishti | +0.0045 [−0.0202, +0.0308] | +0.0296 [−0.0069, +0.0689] |
| HYGD | −0.0045 [−0.0191, +0.0119] | −0.0189 [−0.0278, −0.0107] |
| OCTDL | −0.0001 [−0.0002, +0.0000] | +0.0003 [−0.0004, +0.0011] |
| CAVRI-H5 | +0.0000 [+0.0000, +0.0000] | +0.0000 [−0.0000, +0.0000] |
| Srinivasan | +0.0000 [+0.0000, +0.0000] | −0.0003 [−0.0008, +0.0000] |

**Gain routing versus matched hash budget.**

| Resource | AUROC difference [95% interval] | Brier difference [95% interval] |
| --- | --- | --- |
| PAPILA | +0.0046 [−0.0067, +0.0193] | +0.0148 [+0.0079, +0.0211] |
| Chaksu | +0.0016 [−0.0054, +0.0090] | +0.0126 [+0.0101, +0.0154] |
| RIM-ONE | +0.0051 [−0.0096, +0.0202] | +0.0002 [−0.0026, +0.0027] |
| Drishti | +0.0147 [−0.0161, +0.0505] | +0.0090 [−0.0311, +0.0453] |
| HYGD | −0.0037 [−0.0172, +0.0101] | −0.0187 [−0.0258, −0.0114] |
| OCTDL | +0.0000 [−0.0002, +0.0003] | +0.0006 [−0.0003, +0.0016] |
| CAVRI-H5 | +0.0000 [+0.0000, +0.0000] | +0.0000 [−0.0000, +0.0000] |
| Srinivasan | +0.0000 [+0.0000, +0.0000] | −0.0004 [−0.0007, −0.0002] |

**Historical stronger control.** The earlier DINOv2-B hard-label linear adapter additionally receives source-development labels, so this is a contextual comparison rather than the newly matched external-only supervision control.

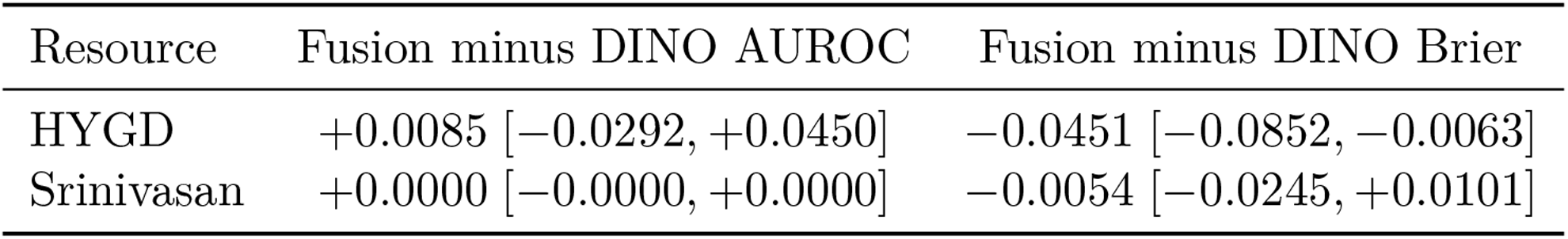

| Resource | Fusion minus DINO AUROC | Fusion minus DINO Brier |
| --- | --- | --- |
| HYGD | +0.0085 [−0.0292, +0.0450] | −0.0451 [−0.0852, −0.0063] |
| Srinivasan | +0.0000 [−0.0000, +0.0000] | −0.0054 [−0.0245, +0.0101] |

### 3.5 Executable scope and text-resource audit

The reusable probability-level interface validates task and endpoint names, aligned class dimensions, finite normalized expert outputs, and image-call caps. A generalist callback receives only selected indices. A zero-weight generalist is skipped while retaining any changed specialist weights and class intercepts. Replay checks cover 45 dataset/size/policy combinations and reject an unsupported pure-text clinical endpoint. The supplied two-profile bundle uses seed 2027 as an example, not a best-seed selection.

FLAIR-adapted clinical BERT encodes candidate descriptions in the original controlled experiments. This does not constitute trained patient-text diagnosis. The local VOLMO assessment/plan JSON contains 40 records, each with five prompt and answer variants; its README count of 89 does not match this file. The records combine images and clinical narratives. At least one inspected narrative explicitly contains a diagnostic finding, so a pure-text evaluation requires case-level input/target leakage and image-dependence auditing and a defensible closed-label mapping. These records are neither added to training nor reported as a third clinically validated modality in this release. StartLux is based on a Qwen architecture; an unmodified Qwen model has not been separately evaluated here.

## 4 Exploratory encoder and language-prior extension

This extension was designed after inspecting the previously evaluated external resources. It is not an untouched external validation. The original source fit, development, calibration, routing, and test partitions, three seeds (2027–2029), 30-epoch maximum, five-epoch patience, and development-NLL checkpoint selection are retained. Four additional frozen visual representations (RETFound-DINOv3, DINOv3-L, C-RADIOv4-SO400M, and RETFound-Green) are compared with the original encoders. OphthaBERT v2, Qwen3-Embedding-0.6B, and MedEmbed-large-v0.1 replace only the candidate-description encoder. Seven new image/text combinations each have fixed, identity, and text heads on two source tasks and three seeds (126 fits); hard-target and reader-target linear controls add 36 fits. Source calibration is transferred unchanged, with no external fitting, temperature selection, or model selection. These source-transfer arms differ from the clinically adapted fusion models in the main study.

All visual inputs derive from the same stored 224-pixel images. DINOv3 models and C-RADIO use 224-pixel network inputs; RETFound-Green uses the existing bicubic 392-pixel loader and cached source features verified by live replay (minimum cosine similarity above 0.9999). This is a controlled information-input contract, not a claim to exhaust each model at its optimal native resolution. RETFound-DINOv3 overlays the retinal teacher weights on the corresponding generic DINOv3 model; the absent register-token tensor retains the generic initialization. The available RETFound-DINOv3 checkpoint documents fundus pretraining, not OCT pretraining. C-RADIO is a single distilled encoder; its SAM-derived teacher supervision does not add a segmentation model to runtime. OphthaBERT uses attention-mask mean pooling, Qwen3 the last non-padding token, and MedEmbed its released CLS pooling. Tokenization, pooling, dimensions, local file hashes, and upstream revisions are recorded. Candidate encoders have different dimensions, so cross-tower head counts also differ; identity controls match capacity within each tower. No independent patient-record classification task is claimed. Models and loading contracts follow their public releases [1–6].

Eight cohorts retain their native endpoints. Exact duplicate images and exact source overlaps are excluded; HYGD uses 737 unique images in 283 patient/pixel components. Srinivasan averages original four-class probabilities within subjects before conditioning on DME/normal, yielding the existing 30-subject primary comparison. The core extension has 720 external rows and the additional linear evaluation 234. Every fixed model, head, seed, and cohort is retained. Paired intervals use 500 shared cluster resamples and average metrics across separately fitted seeds, not probabilities across a seed ensemble. They are exploratory, unadjusted for multiple comparisons, and conditional on fitted models.

**Table 10:** Text-conditioned source-transfer AUROC; mean of three fixed-seed metrics. No cohort-specific selection.

| Image / text | PAPILA | CHAKSU | RIM-ONE | Drishti | HYGD | OCTDL | CAVRI | Srin. |
| --- | --- | --- | --- | --- | --- | --- | --- | --- |
| RETFound-DINOv3 / MedSigLIP | 0.8949 | 0.8048 | 0.6823 | 0.9088 | 0.7402 | 0.9639 | 0.8625 | 0.9600 |
| DINOv3-L / MedSigLIP | 0.8347 | 0.7998 | 0.8550 | 0.9166 | 0.6430 | 0.9984 | 0.9679 | 1.0000 |
| C-RADIOv4 / MedSigLIP | 0.8361 | 0.7832 | 0.7743 | 0.8802 | 0.5134 | 0.9984 | 0.9527 | 0.9985 |
| RETFound-Green / MedSigLIP | 0.7836 | 0.7847 | 0.8118 | 0.8949 | 0.5954 | 0.9949 | 0.9680 | 1.0000 |
| DINOv2-B / OphthaBERT v2 | 0.8014 | 0.7743 | 0.7736 | 0.9104 | 0.7432 | 0.9989 | 0.9653 | 1.0000 |
| DINOv2-B / Qwen3-Emb 0.6B | 0.7968 | 0.7724 | 0.7665 | 0.9149 | 0.7162 | 0.9973 | 0.9645 | 1.0000 |
| DINOv2-B / MedEmbed-L | 0.7924 | 0.7706 | 0.7597 | 0.9154 | 0.6941 | 0.9984 | 0.9647 | 1.0000 |
| DINOv2-B / MedSigLIP | 0.8015 | 0.7797 | 0.7369 | 0.9184 | 0.6593 | 0.9989 | 0.9645 | 1.0000 |
| MedSigLIP / MedSigLIP | 0.8140 | 0.8731 | 0.6929 | 0.8874 | 0.6951 | 0.9980 | 0.9725 | 1.0000 |
| DINOv2-B / FLAIR-BERT | 0.8108 | 0.7830 | 0.7493 | 0.9243 | 0.6571 | 0.9981 | 0.9681 | 1.0000 |

The retinal overlay improves HYGD discrimination over the generic architecture control, but this does not establish superiority to the original strongest linear classifier. The OphthaBERT text– identity AUROC difference is small with an interval crossing zero, while its Brier difference favors the identity control. The large variation in probability loss, along with negative OCT transfer for the fundus-pretrained retinal model, supports explicit modality and endpoint profiles. These results do not demonstrate universal improvement, independent clinical benefit, or a new ophthalmic text foundation model.

**Figure 5:**
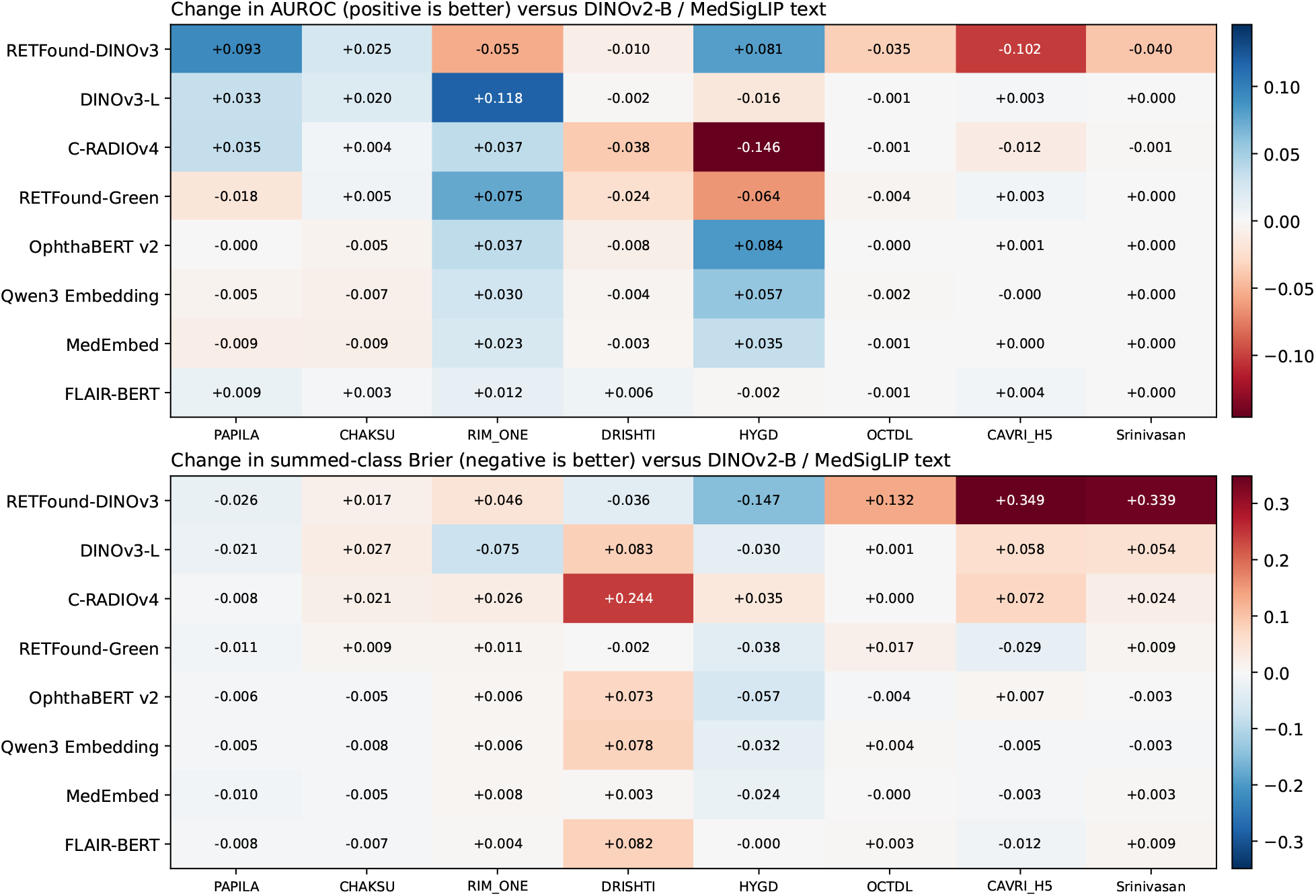
External encoder changes relative to DINOv2-B images and MedSigLIP text, averaged over three seeds. Benefits differ across cohorts and proper-score versus ranking outcomes. All models use the original source calibration.

**Table 11:** Text-conditioned source-transfer summed-class Brier; mean of three fixed-seed metrics. No cohort-specific selection.

| Image / text | PAPILA | CHAKSU | RIM-ONE | Drishti | HYGD | OCTDL | CAVRI | Srin. |
| --- | --- | --- | --- | --- | --- | --- | --- | --- |
| RETFound-DINOv3 / MedSigLIP | 0.2802 | 0.2455 | 0.7010 | 0.7676 | 1.1532 | 0.1600 | 0.6705 | 0.4173 |
| DINOv3-L / MedSigLIP | 0.2850 | 0.2554 | 0.5800 | 0.8863 | 1.2703 | 0.0289 | 0.3799 | 0.1314 |
| C-RADIOv4 / MedSigLIP | 0.2983 | 0.2494 | 0.6807 | 1.0479 | 1.3361 | 0.0282 | 0.3938 | 0.1013 |
| RETFound-Green / MedSigLIP | 0.2957 | 0.2379 | 0.6653 | 0.8016 | 1.2631 | 0.0450 | 0.2931 | 0.0870 |
| DINOv2-B / OphthaBERT v2 | 0.3005 | 0.2235 | 0.6602 | 0.8768 | 1.2432 | 0.0235 | 0.3289 | 0.0751 |
| DINOv2-B / Qwen3-Emb 0.6B | 0.3012 | 0.2206 | 0.6612 | 0.8810 | 1.2683 | 0.0319 | 0.3167 | 0.0746 |
| DINOv2-B / MedEmbed-L | 0.2964 | 0.2241 | 0.6627 | 0.8064 | 1.2762 | 0.0276 | 0.3186 | 0.0810 |
| DINOv2-B / MedSigLIP | 0.3064 | 0.2287 | 0.6547 | 0.8035 | 1.3006 | 0.0278 | 0.3218 | 0.0778 |
| MedSigLIP / MedSigLIP | 0.2960 | 0.2424 | 0.6814 | 0.8002 | 1.2316 | 0.0386 | 0.3104 | 0.0950 |
| DINOv2-B / FLAIR-BERT | 0.2985 | 0.2213 | 0.6587 | 0.8850 | 1.3005 | 0.0312 | 0.3095 | 0.0866 |

**Table 12:** Hard-target linear AUROC under the same frozen-source evaluation. Reader-target and all proper-score results are retained in the aggregate CSV.

| Linear image backbone | PAPILA | CHAKSU | RIM-ONE | Drishti | HYGD | OCTDL | CAVRI | Srin. |
| --- | --- | --- | --- | --- | --- | --- | --- | --- |
| RETFound-DINOv3 | 0.8775 | 0.7008 | 0.6225 | 0.8908 | 0.7712 | 0.9653 | 0.8521 | 0.9748 |
| DINOv3-L | 0.7822 | 0.7425 | 0.9002 | 0.8825 | 0.6934 | 0.9977 | 0.9682 | 0.9956 |
| C-RADIOv4 | 0.7895 | 0.7538 | 0.7096 | 0.8679 | 0.5771 | 0.9980 | 0.9646 | 1.0000 |
| RETFound-Green | 0.7551 | 0.7324 | 0.7710 | 0.9127 | 0.7537 | 0.9950 | 0.9681 | 0.9941 |
| DINOv2-B | 0.7975 | 0.7012 | 0.6577 | 0.8836 | 0.7893 | 0.9996 | 0.9647 | 1.0000 |
| MedSigLIP | 0.8227 | 0.7144 | 0.7901 | 0.8868 | 0.6780 | 0.9954 | 0.9722 | 1.0000 |

**Table 13:** Frozen-source wording robustness. Canonical averages three seeds; held-out averages the same seeds and three predetermined descriptions, using unchanged canonical temperature.

| Task / text | Canonical Brier | Held-out Brier | Difference | Canonical AUC | Held-out AUC |
| --- | --- | --- | --- | --- | --- |
| fundus / MedSigLIP | 0.0476 | 0.0917 | 0.0441 | 0.9213 | 0.9210 |
| fundus / FLAIR-BERT | 0.0472 | 0.2993 | 0.2522 | 0.9185 | 0.6396 |
| fundus / OphthaBERT v2 | 0.0462 | 0.0504 | 0.0042 | 0.9236 | 0.9236 |
| fundus / Qwen3-Emb 0.6B | 0.0467 | 0.0511 | 0.0043 | 0.9209 | 0.9209 |
| fundus / MedEmbed-L | 0.0477 | 0.0487 | 0.0010 | 0.9199 | 0.9200 |
| oct / MedSigLIP | 0.2726 | 0.2872 | 0.0146 | 0.9871 | 0.9844 |
| oct / FLAIR-BERT | 0.2530 | 0.2565 | 0.0035 | 0.9906 | 0.9866 |
| oct / OphthaBERT v2 | 0.2675 | 0.2939 | 0.0263 | 0.9885 | 0.9823 |
| oct / Qwen3-Emb 0.6B | 0.2590 | 0.2588 | -0.0002 | 0.9878 | 0.9867 |
| oct / MedEmbed-L | 0.2693 | 0.2747 | 0.0054 | 0.9873 | 0.9852 |

**Table 14:** Paired source-transfer contrasts. Within-tower identity comparisons were added after the encoder-level results; no causal semantic interpretation is inferred.

| Cohort | First minus second | AUROC difference [95% CI] | Brier difference [95% CI] |
| --- | --- | --- | --- |
| hygd | RETFound-DINOv3 vs DINOv3-L | +0.0972 [+0.0519, +0.1414] | -0.1171 [-0.1651, -0.0693] |
| hygd | RETFound-DINOv3 vs DINOv2-B | +0.0809 [+0.0324, +0.1247] | -0.1474 [-0.1945, -0.1035] |
| hygd | C-RADIOv4 vs DINOv2-B | -0.1460 [-0.1964, -0.0951] | +0.0355 [+0.0034, +0.0596] |
| hygd | RETFound-Green vs DINOv2-B | -0.0639 [-0.1096, -0.0239] | -0.0375 [-0.0727, -0.0086] |
| hygd | OphthaBERT v2 vs MedSigLIP | +0.0838 [+0.0620, +0.1046] | -0.0574 [-0.0713, -0.0448] |
| hygd | MedEmbed-L vs OphthaBERT v2 | -0.0491 [-0.0629, -0.0333] | +0.0330 [+0.0235, +0.0439] |
| hygd | Qwen3-Emb 0.6B vs OphthaBERT v2 | -0.0269 [-0.0380, -0.0165] | +0.0251 [+0.0177, +0.0346] |
| hygd | OphthaBERT v2 vs ID | +0.0071 [-0.0055, +0.0196] | +0.0128 [+0.0023, +0.0223] |
| hygd | RETFound-DINOv3 text vs ID | +0.0073 [-0.0005, +0.0153] | -0.0037 [-0.0115, +0.0056] |
| srinivasan | RETFound-DINOv3 vs DINOv3-L | -0.0400 [-0.1045, +0.0000] | +0.2858 [+0.1492, +0.4536] |
| srinivasan | RETFound-DINOv3 vs DINOv2-B | -0.0400 [-0.1045, +0.0000] | +0.3395 [+0.1675, +0.5216] |
| srinivasan | C-RADIOv4 vs DINOv2-B | -0.0015 [-0.0089, +0.0000] | +0.0235 [-0.0135, +0.0645] |
| srinivasan | RETFound-Green vs DINOv2-B | +0.0000 [+0.0000, +0.0000] | +0.0092 [-0.0155, +0.0424] |
| srinivasan | OphthaBERT v2 vs MedSigLIP | +0.0000 [+0.0000, +0.0000] | -0.0027 [-0.0098, +0.0042] |
| srinivasan | MedEmbed-L vs OphthaBERT v2 | +0.0000 [+0.0000, +0.0000] | +0.0058 [-0.0036, +0.0162] |
| srinivasan | Qwen3-Emb 0.6B vs OphthaBERT v2 | +0.0000 [+0.0000, +0.0000] | -0.0006 [-0.0151, +0.0151] |
| srinivasan | OphthaBERT v2 vs ID | +0.0000 [+0.0000, +0.0000] | +0.0028 [-0.0038, +0.0097] |
| srinivasan | RETFound-DINOv3 text vs ID | +0.0104 [-0.0145, +0.0409] | -0.0100 [-0.0272, +0.0079] |

**Table 15:** Strong linear-control intervals, added to assess whether nonlinear candidate-head gains survive a conventional baseline.

| Cohort | Linear contrast | Target | AUROC difference [95% CI] | Brier difference [95% CI] |
| --- | --- | --- | --- | --- |
| hygd | RETFound-DINOv3 vs DINOv2-B | hard | -0.0181 [-0.0650, +0.0313] | -0.0890 [-0.1257, -0.0513] |
| hygd | RETFound-DINOv3 vs DINOv3-L | hard | +0.0778 [+0.0340, +0.1234] | -0.0322 [-0.0730, +0.0075] |
| hygd | C-RADIOv4 vs DINOv2-B | hard | -0.2123 [-0.2577, -0.1695] | +0.0342 [+0.0108, +0.0538] |
| hygd | RETFound-Green vs DINOv2-B | hard | -0.0356 [-0.0841, +0.0131] | -0.0752 [-0.1026, -0.0534] |
| hygd | RETFound-DINOv3 vs DINOv2-B | reader_soft | +0.0034 [-0.0462, +0.0517] | -0.0802 [-0.1173, -0.0431] |
| srinivasan | RETFound-DINOv3 vs DINOv2-B | hard | -0.0252 [-0.0722, +0.0000] | +0.3566 [+0.1841, +0.5541] |
| srinivasan | RETFound-DINOv3 vs DINOv3-L | hard | -0.0207 [-0.0565, +0.0000] | +0.2866 [+0.1365, +0.4541] |
| srinivasan | C-RADIOv4 vs DINOv2-B | hard | +0.0000 [+0.0000, +0.0000] | +0.0113 [-0.0114, +0.0353] |
| srinivasan | RETFound-Green vs DINOv2-B | hard | -0.0059 [-0.0305, +0.0000] | +0.0714 [+0.0227, +0.1250] |

A separate MedGemma 1.5 4B mechanism pilot uses the same first 32 label-blind hash-selected images per cohort as the archived StartLux controls (256 images; 768 MedGemma calls). Canonical, reversed-option, and no-image inputs use the frozen candidate descriptions. MedGemma scores the first-answer A/B/C/D tokens and normalizes only over those options; it uses its native image processor on the same 224-pixel cache. Reversed outputs are mapped back to semantic classes. Mean total variation (half the summed absolute probability difference) measures option-order sensitivity and image-ablation sensitivity. The restricted token scores are not calibrated clinical risks, and larger image sensitivity is not evidence of greater diagnostic accuracy. This pilot supplies no full-cohort performance claim or matched wall-clock speedup. MedGemma is loaded in BF16 with batch one; observed peak allocated memory is 8.14 GiB under a 13 GiB allocator cap. The model follows the official release [7].

**Table 16:**
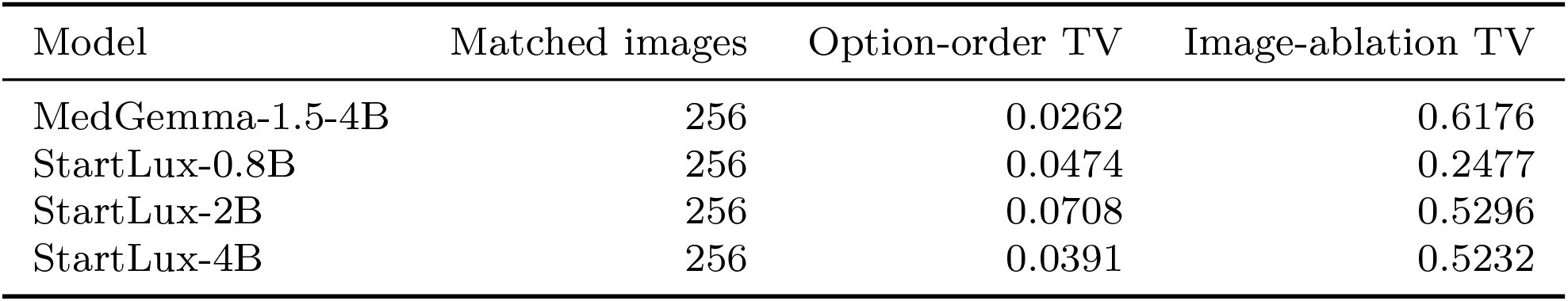
Medical-VLM mechanism pilot on the same 256 images; all three generalist sizes retained. No outcome superiority is inferred from these sensitivity measures.

## 5 Medical priors, transfer and expanded expert banks

All experiments in this section are exploratory and were performed after inspection of the existing cohorts. MedKCO exposure flags apply to its scores and to every fusion containing its features. Source-training splits remain isolated for readout fitting, but this does not remove pretrained-model exposure.

**Table 17:** Released recipe evidence, not a checkpoint-specific pixel-membership certificate.

| Resource | MedKCO exposure evidence |
| --- | --- |
| AIROGS / source OCT | Listed in official pretraining recipe/manifests |
| PAPILA / Chaksu / Drishti | Listed in CFP pretraining recipe/manifests |
| Srinivasan | 495 path matches; all 45 subjects represented |
| HYGD / RIM-ONE / CAVRI | No named-manifest match; unseen status unproven |
| OCTDL | Downstream recipe listed; unseen status unproven |

The new visual assets use strict backbone loading and weight hashes. MerMED is used as a released model resource; its repository publication claim is not independently certified here. The word System One describes bounded non-autoregressive specialist scoring, not a biological cognitive mechanism or a guarantee of zero reasoning cost. StartLux remains an optional separately evaluated expert. No new images, patients, clinical reader observations, or guaranteed risk bounds were synthesized.

## 6 Earlier worst-cohort adaptation extension

### 6.1 Worst-cohort fitting exposes the limits of a universal repair

We fit 24 additional constrained adapters with the same six experts, supervision, regularizers, and bias bounds, replacing average cohort loss by maximum cohort loss. No new hyperparameter sweep or HYGD/Srinivasan fitting is performed. This extension follows inspection of all eight cohorts and is therefore exploratory. On HYGD, mean Brier decreases from 0.7377 to 0.5626 (paired difference-0.1751, 95% cluster interval [-0.2053,-0.1427]), while AUROC changes only from 0.7128 to 0.7129 and accuracy rises from 50.25% to 58.07%. Brier also decreases on Drishti, RIM-ONE, and Srinivasan, but increases on PAPILA, Cháks.u, OCTDL, and CAVRI-H5. Thus the new objective improves neither every cohort nor every metric. These are post-inspection comparisons against the previous fusion adapter, not evidence that the robust adapter dominates all matched single-expert controls. Figure 7 reports every cohort contrast. The probability-loss gain does not establish better screening. With each adapter’s threshold fixed using the original source threshold-selection set, HYGD sensitivity changes from 25.31% to 25.93% (paired difference 0.62 percentage points; 95% interval [−0.25, 1.47]), while specificity changes from 96.62% to 95.94%. Under an illustrative 5% prevalence scenario, missed cases change from 37.35 to 37.04 per 1,000 assessments and unnecessary referrals from 32.15 to 38.58. The paired missed-case difference is −0.31 per 1,000 (95% interval [−0.73, 0.12]). These projections assume unchanged sensitivity and specificity; they are not observed population outcomes.

**Figure 6:**
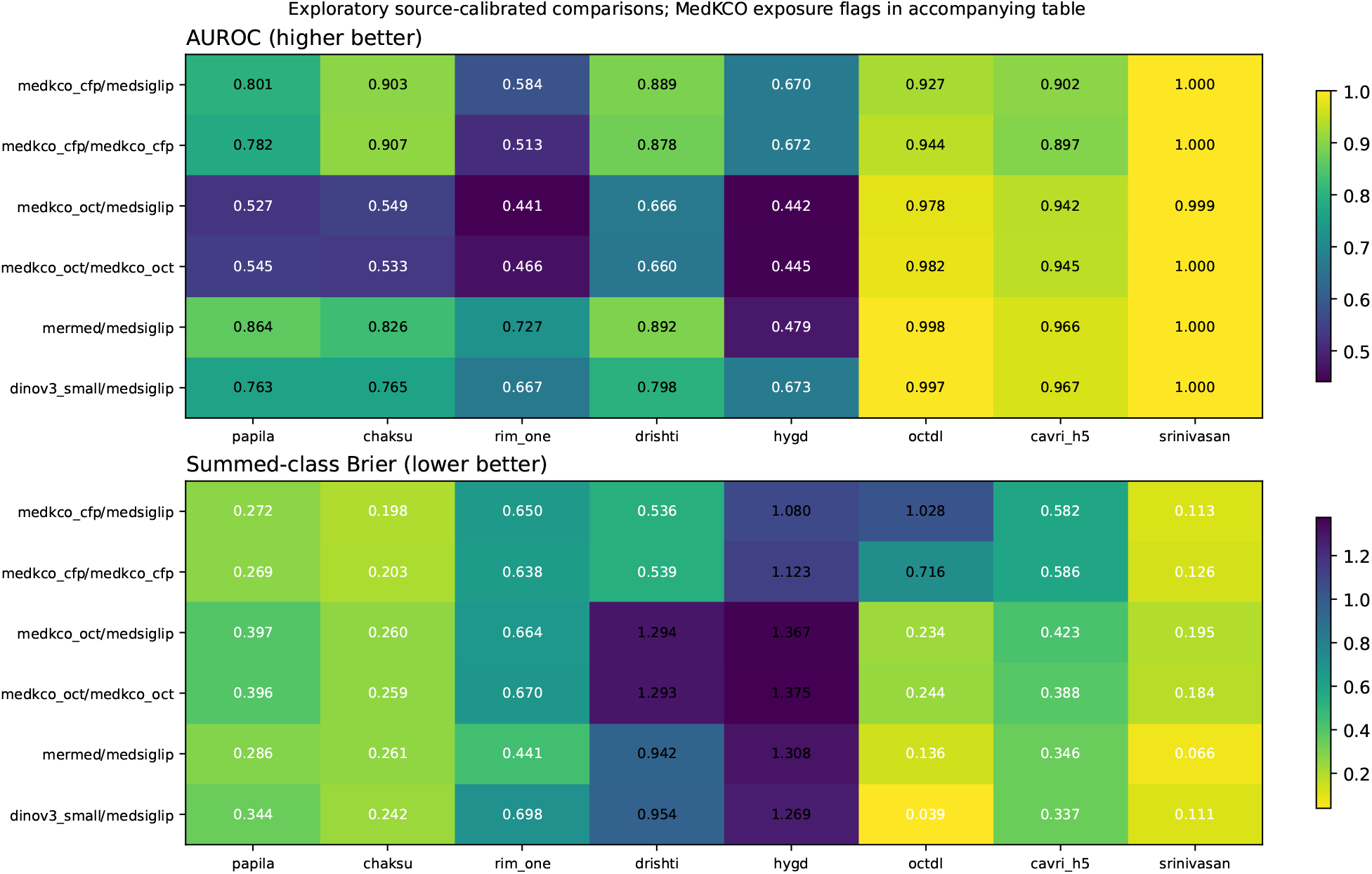
All medical-encoder text-head outcomes. Colors describe absolute performance rather than significance. MedKCO exposure flags in the accompanying table apply throughout; Srinivasan is not an unseen-patient MedKCO validation.

**Table 18:**
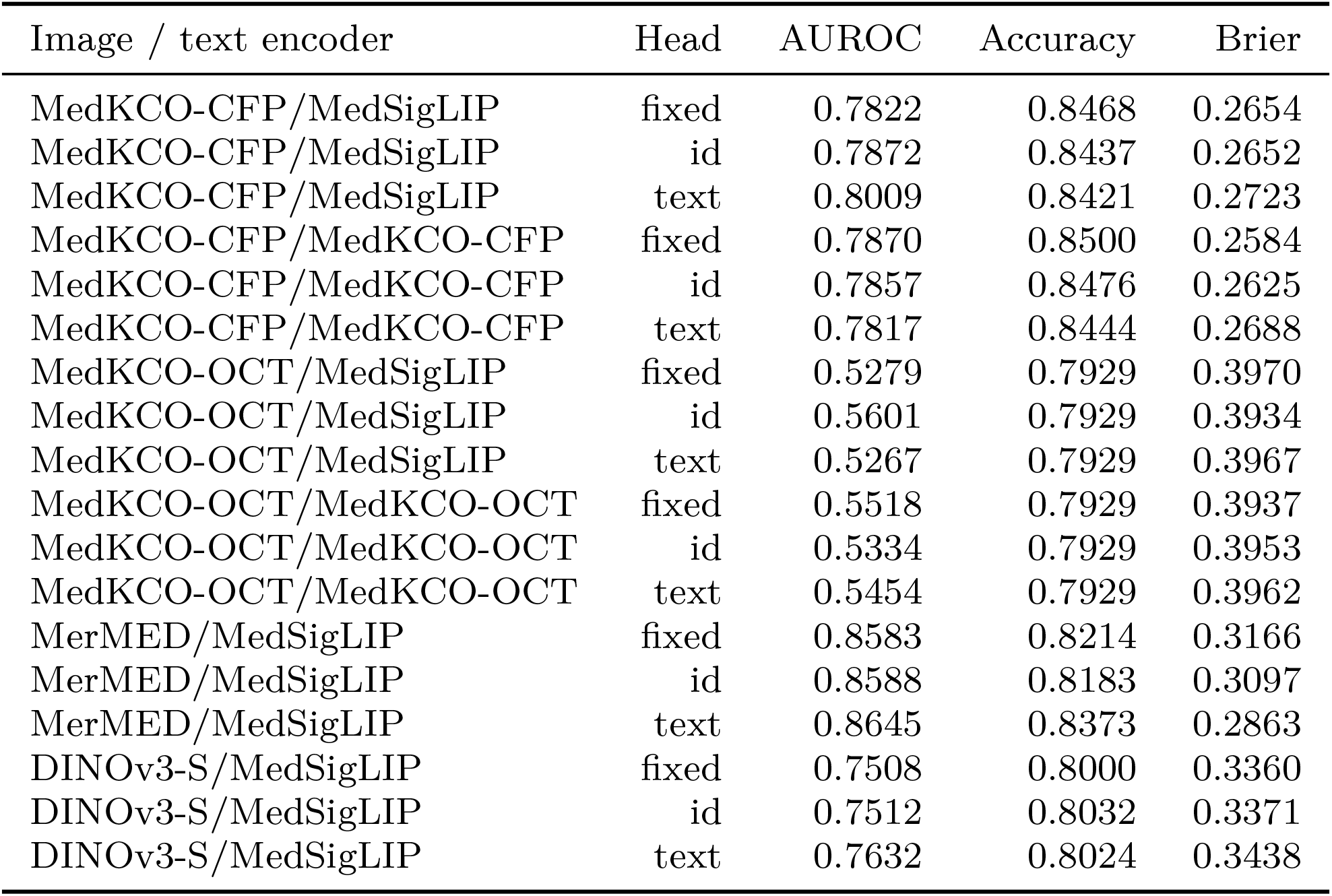
papila: three-seed means, recipe-exposure flagged. Fixed/identity/text heads share the source split; identity controls match text-head capacity.

**Table 19:** chaksu: three-seed means, recipe-exposure flagged. Fixed/identity/text heads share the source split; identity controls match text-head capacity.

| Image / text encoder | Head | AUROC | Accuracy | Brier |
| --- | --- | --- | --- | --- |
| MedKCO-CFP/MedSigLIP | fixed | 0.9065 | 0.8762 | 0.1974 |
| MedKCO-CFP/MedSigLIP | id | 0.9075 | 0.8772 | 0.1966 |
| MedKCO-CFP/MedSigLIP | text | 0.9035 | 0.8754 | 0.1977 |
| MedKCO-CFP/MedKCO-CFP | fixed | 0.9067 | 0.8762 | 0.1968 |
| MedKCO-CFP/MedKCO-CFP | id | 0.9074 | 0.8774 | 0.1979 |
| MedKCO-CFP/MedKCO-CFP | text | 0.9074 | 0.8739 | 0.2027 |
| MedKCO-OCT/MedSigLIP | fixed | 0.5513 | 0.8600 | 0.2574 |
| MedKCO-OCT/MedSigLIP | id | 0.5354 | 0.8602 | 0.2574 |
| MedKCO-OCT/MedSigLIP | text | 0.5493 | 0.8602 | 0.2596 |
| MedKCO-OCT/MedKCO-OCT | fixed | 0.5326 | 0.8602 | 0.2586 |
| MedKCO-OCT/MedKCO-OCT | id | 0.5401 | 0.8602 | 0.2578 |
| MedKCO-OCT/MedKCO-OCT | text | 0.5327 | 0.8602 | 0.2586 |
| MerMED/MedSigLIP | fixed | 0.8065 | 0.8605 | 0.2636 |
| MerMED/MedSigLIP | id | 0.8130 | 0.8607 | 0.2628 |
| MerMED/MedSigLIP | text | 0.8263 | 0.8605 | 0.2615 |
| DINOv3-S/MedSigLIP | fixed | 0.7555 | 0.8600 | 0.2454 |
| DINOv3-S/MedSigLIP | id | 0.7605 | 0.8625 | 0.2393 |
| DINOv3-S/MedSigLIP | text | 0.7655 | 0.8635 | 0.2416 |

**Table 20:** rim_one: three-seed means, pretraining absence unverified. Fixed/identity/text heads share the source split; identity controls match text-head capacity.

| Image / text encoder | Head | AUROC | Accuracy | Brier |
| --- | --- | --- | --- | --- |
| MedKCO-CFP/MedSigLIP | fixed | 0.5245 | 0.6192 | 0.6450 |
| MedKCO-CFP/MedSigLIP | id | 0.5224 | 0.6309 | 0.6237 |
| MedKCO-CFP/MedSigLIP | text | 0.5840 | 0.6433 | 0.6502 |
| MedKCO-CFP/MedKCO-CFP | fixed | 0.5077 | 0.6364 | 0.6355 |
| MedKCO-CFP/MedKCO-CFP | id | 0.4840 | 0.6289 | 0.6473 |
| MedKCO-CFP/MedKCO-CFP | text | 0.5134 | 0.6460 | 0.6378 |
| MedKCO-OCT/MedSigLIP | fixed | 0.4528 | 0.6454 | 0.6639 |
| MedKCO-OCT/MedSigLIP | id | 0.4150 | 0.6454 | 0.6608 |
| MedKCO-OCT/MedSigLIP | text | 0.4408 | 0.6454 | 0.6635 |
| MedKCO-OCT/MedKCO-OCT | fixed | 0.4301 | 0.6454 | 0.6672 |
| MedKCO-OCT/MedKCO-OCT | id | 0.4406 | 0.6454 | 0.6603 |
| MedKCO-OCT/MedKCO-OCT | text | 0.4659 | 0.6454 | 0.6704 |
| MerMED/MedSigLIP | fixed | 0.7752 | 0.6515 | 0.5075 |
| MerMED/MedSigLIP | id | 0.7886 | 0.6536 | 0.4836 |
| MerMED/MedSigLIP | text | 0.7274 | 0.6378 | 0.4412 |
| DINOv3-S/MedSigLIP | fixed | 0.6768 | 0.6454 | 0.6980 |
| DINOv3-S/MedSigLIP | id | 0.6806 | 0.6454 | 0.6958 |
| DINOv3-S/MedSigLIP | text | 0.6669 | 0.6454 | 0.6983 |

**Table 21:** drishti: three-seed means, recipe-exposure flagged. Fixed/identity/text heads share the source split; identity controls match text-head capacity.

| Image / text encoder | Head | AUROC | Accuracy | Brier |
| --- | --- | --- | --- | --- |
| MedKCO-CFP/MedSigLIP | fixed | 0.8826 | 0.6766 | 0.5208 |
| MedKCO-CFP/MedSigLIP | id | 0.8889 | 0.6799 | 0.5082 |
| MedKCO-CFP/MedSigLIP | text | 0.8886 | 0.6601 | 0.5362 |
| MedKCO-CFP/MedKCO-CFP | fixed | 0.8820 | 0.6667 | 0.5205 |
| MedKCO-CFP/MedKCO-CFP | id | 0.8849 | 0.6766 | 0.5105 |
| MedKCO-CFP/MedKCO-CFP | text | 0.8777 | 0.6700 | 0.5390 |
| MedKCO-OCT/MedSigLIP | fixed | 0.6919 | 0.3069 | 1.2864 |
| MedKCO-OCT/MedSigLIP | id | 0.6929 | 0.3069 | 1.2885 |
| MedKCO-OCT/MedSigLIP | text | 0.6664 | 0.3069 | 1.2937 |
| MedKCO-OCT/MedKCO-OCT | fixed | 0.6790 | 0.3069 | 1.2710 |
| MedKCO-OCT/MedKCO-OCT | id | 0.6859 | 0.3069 | 1.2932 |
| MedKCO-OCT/MedKCO-OCT | text | 0.6596 | 0.3069 | 1.2925 |
| MerMED/MedSigLIP | fixed | 0.8859 | 0.3465 | 1.0631 |
| MerMED/MedSigLIP | id | 0.8903 | 0.3663 | 0.9917 |
| MerMED/MedSigLIP | text | 0.8923 | 0.3993 | 0.9424 |
| DINOv3-S/MedSigLIP | fixed | 0.7955 | 0.4059 | 0.9655 |
| DINOv3-S/MedSigLIP | id | 0.7906 | 0.3861 | 0.9776 |
| DINOv3-S/MedSigLIP | text | 0.7978 | 0.4158 | 0.9541 |

**Table 22:** hygd: three-seed means, pretraining absence unverified. Fixed/identity/text heads share the source split; identity controls match text-head capacity.

| Image / text encoder | Head | AUROC | Accuracy | Brier |
| --- | --- | --- | --- | --- |
| MedKCO-CFP/MedSigLIP | fixed | 0.6691 | 0.3976 | 1.0972 |
| MedKCO-CFP/MedSigLIP | id | 0.6800 | 0.4102 | 1.0785 |
| MedKCO-CFP/MedSigLIP | text | 0.6703 | 0.4089 | 1.0803 |
| MedKCO-CFP/MedKCO-CFP | fixed | 0.6762 | 0.4075 | 1.0770 |
| MedKCO-CFP/MedKCO-CFP | id | 0.6825 | 0.4080 | 1.0839 |
| MedKCO-CFP/MedKCO-CFP | text | 0.6715 | 0.3885 | 1.1226 |
| MedKCO-OCT/MedSigLIP | fixed | 0.4389 | 0.2673 | 1.3653 |
| MedKCO-OCT/MedSigLIP | id | 0.4458 | 0.2673 | 1.3635 |
| MedKCO-OCT/MedSigLIP | text | 0.4423 | 0.2673 | 1.3669 |
| MedKCO-OCT/MedKCO-OCT | fixed | 0.4360 | 0.2673 | 1.3651 |
| MedKCO-OCT/MedKCO-OCT | id | 0.4521 | 0.2673 | 1.3672 |
| MedKCO-OCT/MedKCO-OCT | text | 0.4451 | 0.2673 | 1.3755 |
| MerMED/MedSigLIP | fixed | 0.4748 | 0.2822 | 1.3332 |
| MerMED/MedSigLIP | id | 0.4931 | 0.2840 | 1.3367 |
| MerMED/MedSigLIP | text | 0.4794 | 0.2926 | 1.3075 |
| DINOv3-S/MedSigLIP | fixed | 0.6649 | 0.2763 | 1.2774 |
| DINOv3-S/MedSigLIP | id | 0.6813 | 0.2804 | 1.2664 |
| DINOv3-S/MedSigLIP | text | 0.6731 | 0.2727 | 1.2693 |

**Table 23:** octdl: three-seed means, pretraining absence unverified. Fixed/identity/text heads share the source split; identity controls match text-head capacity.

| Image / text encoder | Head | AUROC | Accuracy | Brier |
| --- | --- | --- | --- | --- |
| MedKCO-CFP/MedSigLIP | fixed | 0.9279 | 0.4557 | 0.9367 |
| MedKCO-CFP/MedSigLIP | id | 0.9443 | 0.6195 | 0.6344 |
| MedKCO-CFP/MedSigLIP | text | 0.9266 | 0.4015 | 1.0284 |
| MedKCO-CFP/MedKCO-CFP | fixed | 0.9406 | 0.4557 | 0.9041 |
| MedKCO-CFP/MedKCO-CFP | id | 0.9407 | 0.5211 | 0.8023 |
| MedKCO-CFP/MedKCO-CFP | text | 0.9441 | 0.5598 | 0.7159 |
| MedKCO-OCT/MedSigLIP | fixed | 0.9573 | 0.7890 | 0.3271 |
| MedKCO-OCT/MedSigLIP | id | 0.9829 | 0.8411 | 0.2191 |
| MedKCO-OCT/MedSigLIP | text | 0.9778 | 0.8432 | 0.2343 |
| MedKCO-OCT/MedKCO-OCT | fixed | 0.9812 | 0.8333 | 0.2440 |
| MedKCO-OCT/MedKCO-OCT | id | 0.9802 | 0.8235 | 0.2748 |
| MedKCO-OCT/MedKCO-OCT | text | 0.9822 | 0.8319 | 0.2436 |
| MerMED/MedSigLIP | fixed | 0.9988 | 0.8896 | 0.1651 |
| MerMED/MedSigLIP | id | 0.9979 | 0.8565 | 0.2243 |
| MerMED/MedSigLIP | text | 0.9978 | 0.9079 | 0.1358 |
| DINOv3-S/MedSigLIP | fixed | 0.9974 | 0.9754 | 0.0395 |
| DINOv3-S/MedSigLIP | id | 0.9972 | 0.9740 | 0.0397 |
| DINOv3-S/MedSigLIP | text | 0.9971 | 0.9733 | 0.0391 |

**Table 24:**
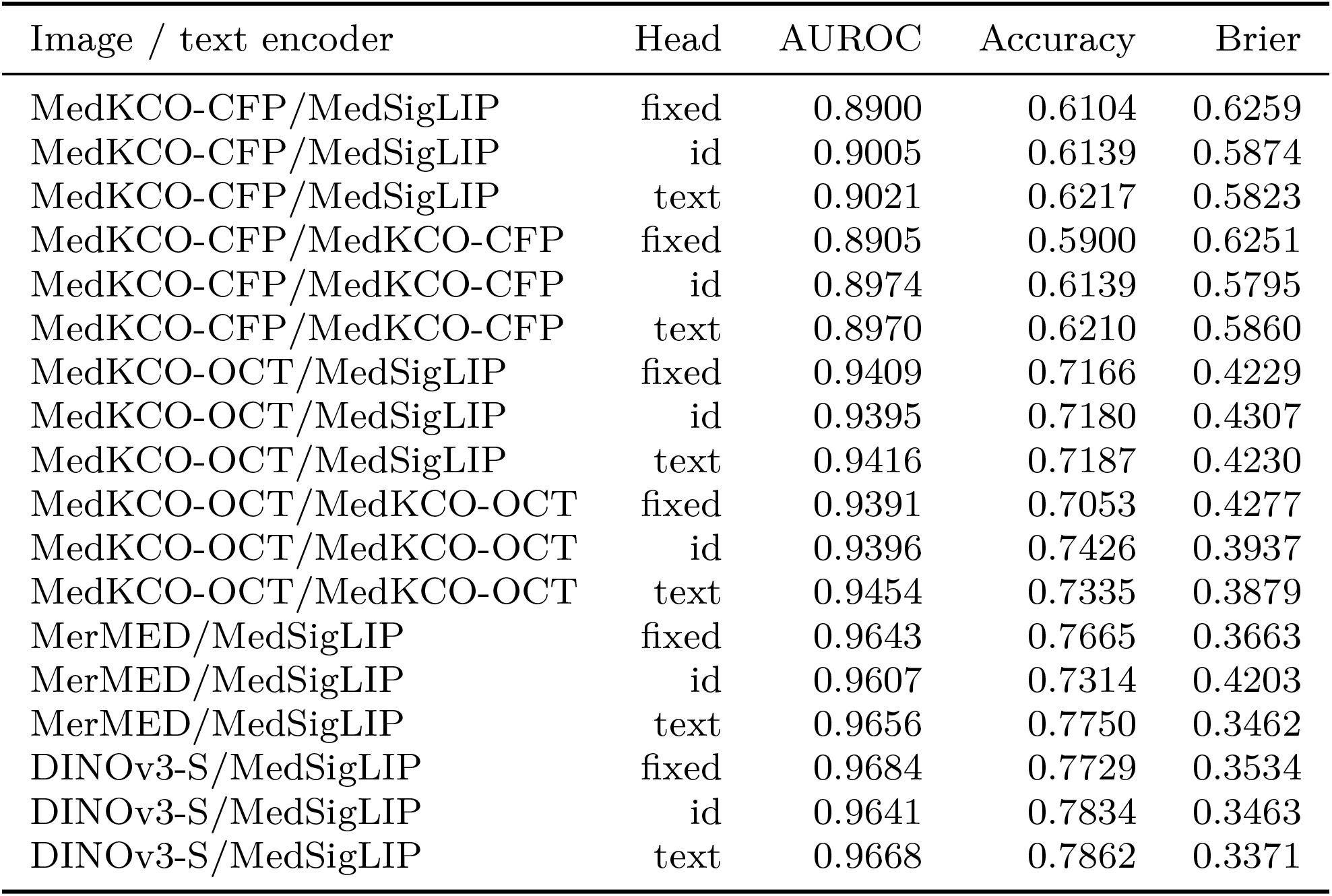
cavri_h5: three-seed means, pretraining absence unverified. Fixed/identity/text heads share the source split; identity controls match text-head capacity.

| Image / text encoder | Head | AUROC | Accuracy | Brier |
| --- | --- | --- | --- | --- |
| MedKCO-CFP/MedSigLIP | fixed | 0.8900 | 0.6104 | 0.6259 |
| MedKCO-CFP/MedSigLIP | id | 0.9005 | 0.6139 | 0.5874 |
| MedKCO-CFP/MedSigLIP | text | 0.9021 | 0.6217 | 0.5823 |
| MedKCO-CFP/MedKCO-CFP | fixed | 0.8905 | 0.5900 | 0.6251 |
| MedKCO-CFP/MedKCO-CFP | id | 0.8974 | 0.6139 | 0.5795 |
| MedKCO-CFP/MedKCO-CFP | text | 0.8970 | 0.6210 | 0.5860 |
| MedKCO-OCT/MedSigLIP | fixed | 0.9409 | 0.7166 | 0.4229 |
| MedKCO-OCT/MedSigLIP | id | 0.9395 | 0.7180 | 0.4307 |
| MedKCO-OCT/MedSigLIP | text | 0.9416 | 0.7187 | 0.4230 |
| MedKCO-OCT/MedKCO-OCT | fixed | 0.9391 | 0.7053 | 0.4277 |
| MedKCO-OCT/MedKCO-OCT | id | 0.9396 | 0.7426 | 0.3937 |
| MedKCO-OCT/MedKCO-OCT | text | 0.9454 | 0.7335 | 0.3879 |
| MerMED/MedSigLIP | fixed | 0.9643 | 0.7665 | 0.3663 |
| MerMED/MedSigLIP | id | 0.9607 | 0.7314 | 0.4203 |
| MerMED/MedSigLIP | text | 0.9656 | 0.7750 | 0.3462 |
| DINOv3-S/MedSigLIP | fixed | 0.9684 | 0.7729 | 0.3534 |
| DINOv3-S/MedSigLIP | id | 0.9641 | 0.7834 | 0.3463 |
| DINOv3-S/MedSigLIP | text | 0.9668 | 0.7862 | 0.3371 |

**Table 25:**
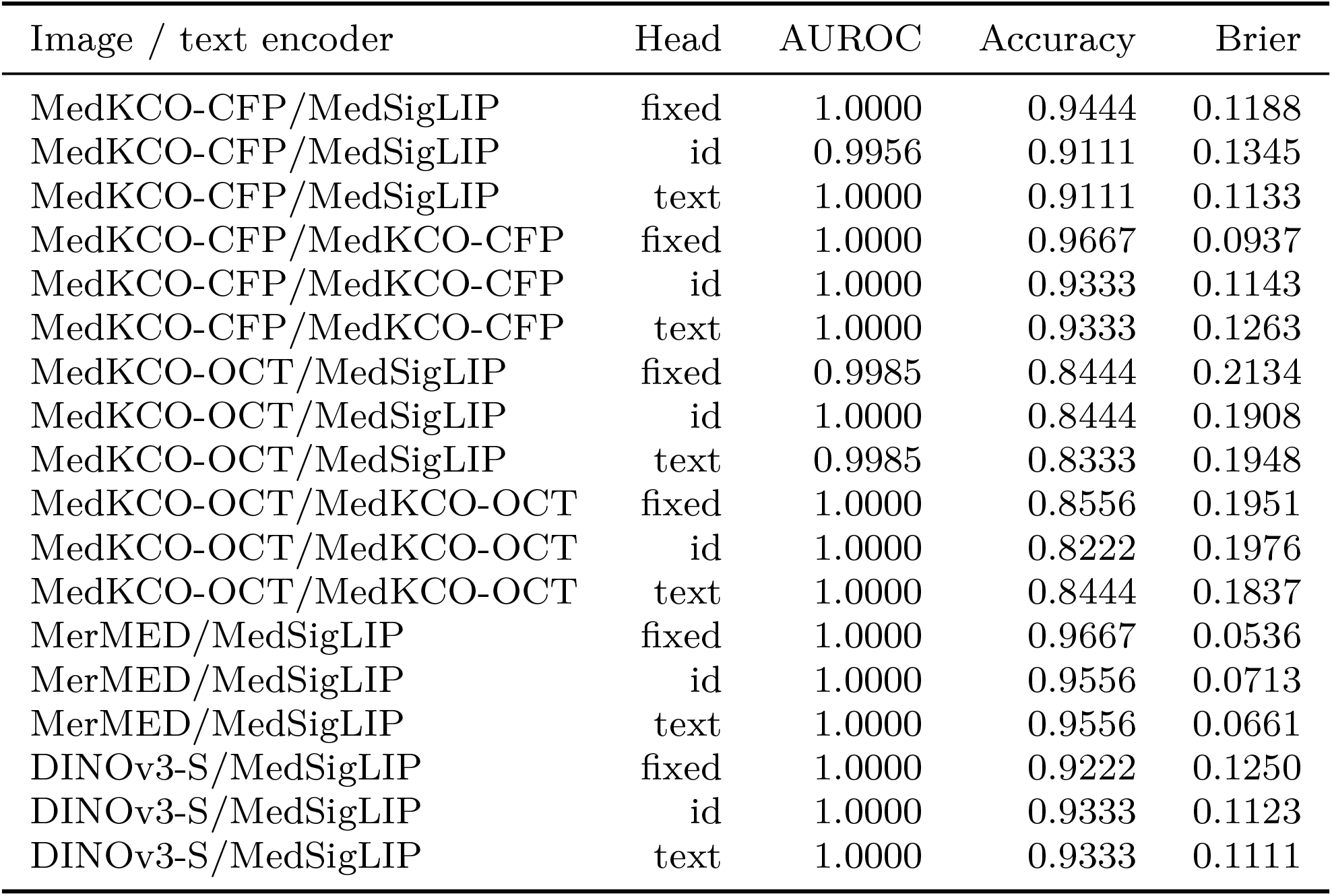
srinivasan: three-seed means, recipe-exposure flagged. Fixed/identity/text heads share the source split; identity controls match text-head capacity.

**Table 26:** Native fixed category names versus official medical descriptions. These have no source readout training; all MedKCO exposure flags still apply.

| Cohort | Image model | Names AUROC | Knowledge AUROC | Names Brier | Knowledge Brier |
| --- | --- | --- | --- | --- | --- |
| papila | MedKCO-CFP | 0.7383 | 0.7338 | 0.7269 | 0.7377 |
| papila | MedKCO-OCT | 0.4498 | 0.5752 | 0.5537 | 0.4107 |
| chaksu | MedKCO-CFP | 0.9066 | 0.9076 | 0.6865 | 0.6629 |
| chaksu | MedKCO-OCT | 0.5009 | 0.4892 | 0.5731 | 0.2787 |
| rim_one | MedKCO-CFP | 0.4664 | 0.4564 | 0.9527 | 0.9108 |
| rim_one | MedKCO-OCT | 0.5790 | 0.3973 | 0.5220 | 0.7075 |
| drishti | MedKCO-CFP | 0.8677 | 0.8691 | 0.3430 | 0.3440 |
| drishti | MedKCO-OCT | 0.5431 | 0.5829 | 0.4688 | 1.3829 |
| hygd | MedKCO-CFP | 0.7417 | 0.7400 | 0.6017 | 0.6315 |
| hygd | MedKCO-OCT | 0.5476 | 0.6018 | 0.4586 | 1.4606 |
| octdl | MedKCO-CFP | 0.3952 | 0.3588 | 0.6893 | 0.7659 |
| octdl | MedKCO-OCT | 0.9032 | 0.9499 | 0.3646 | 0.2424 |
| cavri_h5 | MedKCO-CFP | 0.5645 | 0.4840 | 0.7675 | 0.7979 |
| cavri_h5 | MedKCO-OCT | 0.7422 | 0.7548 | 0.9349 | 0.9307 |
| srinivasan | MedKCO-CFP | 0.2578 | 0.2578 | 0.6229 | 0.6086 |
| srinivasan | MedKCO-OCT | 1.0000 | 1.0000 | 0.4376 | 0.2902 |

**Table 27:**
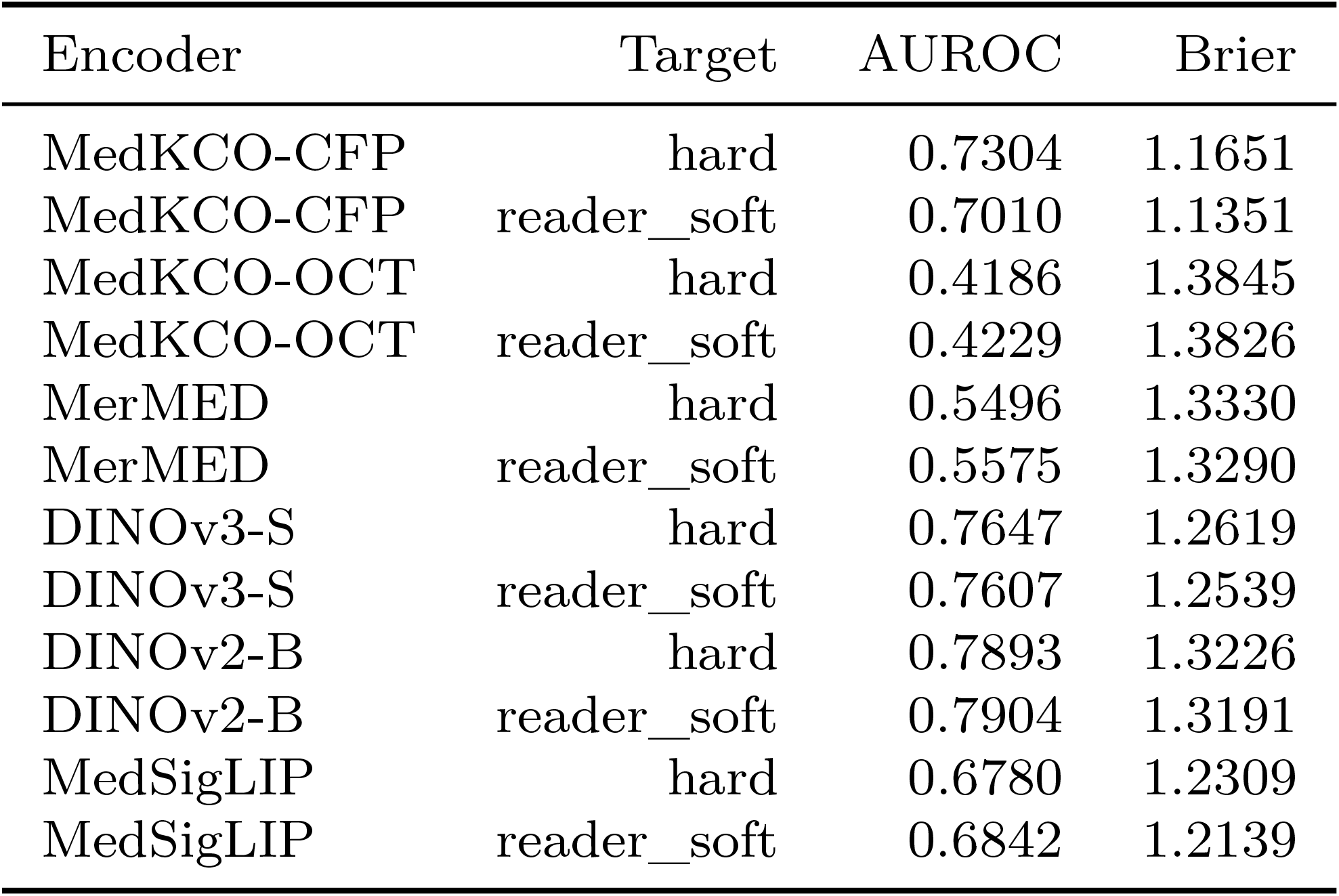
hygd conventional linear controls, three seeds. Complete per-cohort aggregate CSVs accompany the code.

| Encoder | Target | AUROC | Brier |
| --- | --- | --- | --- |
| MedKCO-CFP | hard | 0.7304 | 1.1651 |
| MedKCO-CFP | reader_soft | 0.7010 | 1.1351 |
| MedKCO-OCT | hard | 0.4186 | 1.3845 |
| MedKCO-OCT | reader_soft | 0.4229 | 1.3826 |
| MerMED | hard | 0.5496 | 1.3330 |
| MerMED | reader_soft | 0.5575 | 1.3290 |
| DINOv3-S | hard | 0.7647 | 1.2619 |
| DINOv3-S | reader_soft | 0.7607 | 1.2539 |
| DINOv2-B | hard | 0.7893 | 1.3226 |
| DINOv2-B | reader_soft | 0.7904 | 1.3191 |
| MedSigLIP | hard | 0.6780 | 1.2309 |
| MedSigLIP | reader_soft | 0.6842 | 1.2139 |

**Table 28:** octdl conventional linear controls, three seeds. Complete per-cohort aggregate CSVs accompany the code.

| Encoder | Target | AUROC | Brier |
| --- | --- | --- | --- |
| MedKCO-CFP | hard | 0.9716 | 0.6586 |
| MedKCO-OCT | hard | 0.9741 | 0.1972 |
| MerMED | hard | 0.9979 | 0.0947 |
| DINOv3-S | hard | 0.9966 | 0.0322 |
| DINOv2-B | hard | 0.9996 | 0.0183 |
| MedSigLIP | hard | 0.9954 | 0.0616 |

**Table 29:** cavri_h5 conventional linear controls, three seeds. Complete per-cohort aggregate CSVs accompany the code.

| Encoder | Target | AUROC | Brier |
| --- | --- | --- | --- |
| MedKCO-CFP | hard | 0.9246 | 0.4989 |
| MedKCO-OCT | hard | 0.9244 | 0.4757 |
| MerMED | hard | 0.9703 | 0.3157 |
| DINOv3-S | hard | 0.9650 | 0.3891 |
| DINOv2-B | hard | 0.9647 | 0.3296 |
| MedSigLIP | hard | 0.9722 | 0.3295 |

**Table 30:** srinivasan conventional linear controls, three seeds. Complete per-cohort aggregate CSVs accompany the code.

| Encoder | Target | AUROC | Brier |
| --- | --- | --- | --- |
| MedKCO-CFP | hard | 1.0000 | 0.1065 |
| MedKCO-OCT | hard | 1.0000 | 0.1963 |
| MerMED | hard | 1.0000 | 0.0985 |
| DINOv3-S | hard | 1.0000 | 0.1846 |
| DINOv2-B | hard | 1.0000 | 0.0996 |
| MedSigLIP | hard | 1.0000 | 0.1346 |

**Table 31:** Medical replacement with StartLux 2B. Raw-image call counts are distinct from referral rates; additional specialist encoders have their own cost. A zero learned generalist weight can make calls unproductive. All prior exposure flags apply.

| Cohort | Arm | AUROC | Brier | Call fraction |
| --- | --- | --- | --- | --- |
| papila | specialist | 0.8056 | 0.2340 | 0.000 |
| papila | fusion | 0.8162 | 0.2706 | 1.000 |
| papila | cascade_gain20 | 0.7800 | 0.2392 | 0.199 |
| papila | cascade_hash_matched | 0.7921 | 0.2410 | 0.199 |
| chaksu | specialist | 0.9070 | 0.1570 | 0.000 |
| chaksu | fusion | 0.9033 | 0.1764 | 1.000 |
| chaksu | cascade_gain20 | 0.8998 | 0.1584 | 0.200 |
| chaksu | cascade_hash_matched | 0.8825 | 0.1612 | 0.200 |
| rim_one | specialist | 0.7252 | 0.9928 | 0.000 |
| rim_one | fusion | 0.7035 | 0.7626 | 1.000 |
| rim_one | cascade_gain20 | 0.7208 | 0.9913 | 0.018 |
| rim_one | cascade_hash_matched | 0.7203 | 0.9891 | 0.018 |
| drishti | specialist | 0.8909 | 0.4398 | 0.000 |
| drishti | fusion | 0.8885 | 0.4793 | 1.000 |
| drishti | cascade_gain20 | 0.8843 | 0.4345 | 0.198 |
| drishti | cascade_hash_matched | 0.8770 | 0.4465 | 0.198 |
| hygd | specialist | 0.6022 | 0.9245 | 0.000 |
| hygd | fusion | 0.6990 | 0.7871 | 1.000 |
| hygd | cascade_gain20 | 0.6025 | 0.9114 | 0.199 |
| hygd | cascade_hash_matched | 0.6208 | 0.8955 | 0.199 |
| octdl | specialist | 0.9982 | 0.0542 | 0.000 |
| octdl | fusion | 0.9985 | 0.0464 | 1.000 |
| octdl | cascade_gain20 | 0.9984 | 0.0492 | 0.200 |
| octdl | cascade_hash_matched | 0.9980 | 0.0524 | 0.200 |
| cavri_h5 | specialist | 0.9761 | 0.2799 | 0.000 |
| cavri_h5 | fusion | 0.9761 | 0.2799 | 1.000 |
| cavri_h5 | cascade_gain20 | 0.9761 | 0.2799 | 0.199 |
| cavri_h5 | cascade_hash_matched | 0.9761 | 0.2799 | 0.199 |
| srinivasan | specialist | 1.0000 | 0.1530 | 0.000 |
| srinivasan | fusion | 1.0000 | 0.1520 | 1.000 |
| srinivasan | cascade_gain20 | 1.0000 | 0.1492 | 0.200 |
| srinivasan | cascade_hash_matched | 1.0000 | 0.1526 | 0.200 |

**Table 32:** Original-plus-medical augmentation with StartLux 2B. Raw-image call counts are distinct from referral rates; additional specialist encoders have their own cost. A zero learned generalist weight can make calls unproductive. All prior exposure flags apply.

| Cohort | Arm | AUROC | Brier | Call fraction |
| --- | --- | --- | --- | --- |
| papila | specialist | 0.8305 | 0.2807 | 0.000 |
| papila | fusion | 0.8307 | 0.2818 | 1.000 |
| papila | cascade_gain20 | 0.8305 | 0.2807 | 0.107 |
| papila | cascade_hash_matched | 0.8300 | 0.2810 | 0.107 |
| chaksu | specialist | 0.8620 | 0.1981 | 0.000 |
| chaksu | fusion | 0.8666 | 0.1955 | 1.000 |
| chaksu | cascade_gain20 | 0.8619 | 0.1983 | 0.116 |
| chaksu | cascade_hash_matched | 0.8623 | 0.1978 | 0.116 |
| rim_one | specialist | 0.6843 | 0.6268 | 0.000 |
| rim_one | fusion | 0.6831 | 0.5986 | 1.000 |
| rim_one | cascade_gain20 | 0.6839 | 0.6269 | 0.007 |
| rim_one | cascade_hash_matched | 0.6836 | 0.6267 | 0.007 |
| drishti | specialist | 0.9108 | 0.3897 | 0.000 |
| drishti | fusion | 0.9111 | 0.4122 | 1.000 |
| drishti | cascade_gain20 | 0.9071 | 0.3903 | 0.198 |
| drishti | cascade_hash_matched | 0.9081 | 0.3933 | 0.198 |
| hygd | specialist | 0.7434 | 0.7364 | 0.000 |
| hygd | fusion | 0.7474 | 0.7300 | 1.000 |
| hygd | cascade_gain20 | 0.7463 | 0.7353 | 0.172 |
| hygd | cascade_hash_matched | 0.7426 | 0.7352 | 0.172 |
| octdl | specialist | 0.9996 | 0.0256 | 0.000 |
| octdl | fusion | 0.9997 | 0.0248 | 1.000 |
| octdl | cascade_gain20 | 0.9997 | 0.0250 | 0.200 |
| octdl | cascade_hash_matched | 0.9996 | 0.0253 | 0.200 |
| cavri_h5 | specialist | 0.9789 | 0.2955 | 0.000 |
| cavri_h5 | fusion | 0.9789 | 0.2955 | 1.000 |
| cavri_h5 | cascade_gain20 | 0.9789 | 0.2955 | 0.199 |
| cavri_h5 | cascade_hash_matched | 0.9789 | 0.2955 | 0.199 |
| srinivasan | specialist | 1.0000 | 0.0934 | 0.000 |
| srinivasan | fusion | 1.0000 | 0.0925 | 1.000 |
| srinivasan | cascade_gain20 | 1.0000 | 0.0934 | 0.200 |
| srinivasan | cascade_hash_matched | 1.0000 | 0.0932 | 0.200 |

**Table 33:** hygd: 500 paired connected-group draws, unadjusted exploratory intervals.

| Contrast (first minus second) | AUROC difference [95% CI] | Brier difference [95% CI] |
| --- | --- | --- |
| Matched vs swapped image prior | +0.2280 [+0.1616, +0.3059] | -0.2866 [-0.3635, -0.2210] |
| Native vs common text | +0.0013 [-0.0071, +0.0107] | +0.0423 [+0.0306, +0.0519] |
| Text vs matched identities | -0.0110 [-0.0193, -0.0010] | +0.0387 [+0.0281, +0.0479] |
| MerMED vs DINOv2-B | -0.1800 [-0.2187, -0.1442] | +0.0069 [-0.0181, +0.0333] |
| DINOv3-S vs DINOv2-B | +0.0138 [-0.0255, +0.0542] | -0.0313 [-0.0542, -0.0074] |
| Knowledge vs category names | -0.0017 [-0.0048, +0.0013] | +0.0298 [+0.0211, +0.0383] |

**Table 34:** octdl: 500 paired connected-group draws, unadjusted exploratory intervals.

| Contrast (first minus second) | AUROC difference [95% CI] | Brier difference [95% CI] |
| --- | --- | --- |
| Matched vs swapped image prior | +0.0511 [+0.0202, +0.0805] | -0.7941 [-0.8805, -0.6865] |
| Native vs common text | +0.0045 [+0.0012, +0.0080] | +0.0093 [+0.0011, +0.0173] |
| Text vs matched identities | +0.0020 [-0.0003, +0.0046] | -0.0313 [-0.0450, -0.0198] |
| MerMED vs DINOv2-B | -0.0011 [-0.0034, +0.0006] | +0.1080 [+0.0762, +0.1471] |
| DINOv3-S vs DINOv2-B | -0.0018 [-0.0046, +0.0003] | +0.0113 [-0.0081, +0.0296] |
| Knowledge vs category names | +0.0466 [+0.0301, +0.0651] | -0.1222 [-0.1557, -0.0913] |

**Table 35:** cavri_h5: 500 paired connected-group draws, unadjusted exploratory intervals.

| Contrast (first minus second) | AUROC difference [95% CI] | Brier difference [95% CI] |
| --- | --- | --- |
| Matched vs swapped image prior | +0.0395 [+0.0226, +0.0568] | -0.1593 [-0.2172, -0.0976] |
| Native vs common text | +0.0038 [+0.0003, +0.0075] | -0.0352 [-0.0485, -0.0210] |
| Text vs matched identities | +0.0058 [+0.0016, +0.0097] | -0.0058 [-0.0214, +0.0104] |
| MerMED vs DINOv2-B | +0.0010 [-0.0096, +0.0123] | +0.0244 [-0.0247, +0.0746] |
| DINOv3-S vs DINOv2-B | +0.0023 [-0.0065, +0.0111] | +0.0153 [-0.0204, +0.0528] |
| Knowledge vs category names | +0.0126 [+0.0036, +0.0214] | -0.0042 [-0.0280, +0.0208] |

**Table 36:** Expert-bank and routing comparisons, 500 paired group draws. Additional experts do not guarantee improvement.

| Cohort | Contrast | AUROC difference [95% CI] | Brier difference [95% CI] |
| --- | --- | --- | --- |
| hygd | Expanded vs original fusion | -0.0504 [-0.0667, -0.0351] | +0.1092 [+0.0867, +0.1307] |
| hygd | Expanded vs original specialist | -0.0281 [-0.0417, -0.0138] | +0.1181 [+0.0973, +0.1427] |
| hygd | Gain vs matched hash gate | +0.0037 [+0.0011, +0.0067] | +0.0001 [-0.0008, +0.0011] |
| hygd | Replacement vs original fusion | -0.0989 [-0.1322, -0.0704] | +0.1663 [+0.1293, +0.2003] |
| octdl | Expanded vs original fusion | +0.0003 [-0.0001, +0.0008] | -0.0015 [-0.0059, +0.0035] |
| octdl | Expanded vs original specialist | +0.0003 [-0.0001, +0.0009] | -0.0024 [-0.0081, +0.0040] |
| octdl | Gain vs matched hash gate | +0.0000 [-0.0000, +0.0002] | -0.0002 [-0.0014, +0.0007] |
| octdl | Replacement vs original fusion | -0.0008 [-0.0029, +0.0006] | +0.0201 [+0.0081, +0.0318] |
| cavri_h5 | Expanded vs original fusion | +0.0019 [+0.0003, +0.0041] | -0.0011 [-0.0016, -0.0006] |
| cavri_h5 | Expanded vs original specialist | +0.0019 [+0.0003, +0.0041] | -0.0011 [-0.0016, -0.0006] |
| cavri_h5 | Gain vs matched hash gate | +0.0000 [+0.0000, +0.0000] | +0.0000 [-0.0000, +0.0000] |
| cavri_h5 | Replacement vs original fusion | -0.0010 [-0.0090, +0.0082] | -0.0168 [-0.0511, +0.0160] |

**Table 37:** Batch-one BF16 resident-image encoding on a shared RTX 5090, 20 warmups and 100 repeats. Includes on-device preprocessing; excludes disk, transfers, text encoding, decision head and routing. MedKCO upsamples to 512 and its memory includes the unused text branch. Shared-device timing does not establish exclusive-hardware throughput or LLM speedup.

| Encoder | Median (ms) | 10–90th percentile (ms) | Peak reserved GiB |
| --- | --- | --- | --- |
| DINOv3-S | 8.01 | 7.71–9.02 | 0.12 |
| MerMED | 4.95 | 3.51–5.24 | 0.41 |
| MedKCO-CFP | 3.57 | 3.44–3.84 | 0.61 |
| MedKCO-OCT | 3.58 | 3.35–3.89 | 0.61 |

**Table 38:** Exploratory maximum-cohort versus equal-cohort adaptation. Brier sums class losses; lower is better. Both methods use the same experts and permitted training data. Old-cohort results use leave-one-cohort-out fitting; HYGD and Srinivasan are post-inspection exploratory evaluations.

| Cohort | Equal Brier | Robust Brier | Equal AUROC | Robust AUROC |
| --- | --- | --- | --- | --- |
| cavri-h5 | 0.2710 | 0.2735 | 0.9791 | 0.9793 |
| chaksu | 0.2233 | 0.3238 | 0.8322 | 0.8260 |
| drishti | 0.3202 | 0.2217 | 0.9218 | 0.9226 |
| hygd | 0.7377 | 0.5626 | 0.7128 | 0.7129 |
| octdl | 0.0282 | 0.0318 | 0.9991 | 0.9991 |
| papila | 0.2280 | 0.2616 | 0.8403 | 0.8417 |
| rim-one | 0.5271 | 0.4490 | 0.7919 | 0.7970 |
| srinivasan | 0.0733 | 0.0606 | 1.0000 | 1.0000 |

**Figure 7:**
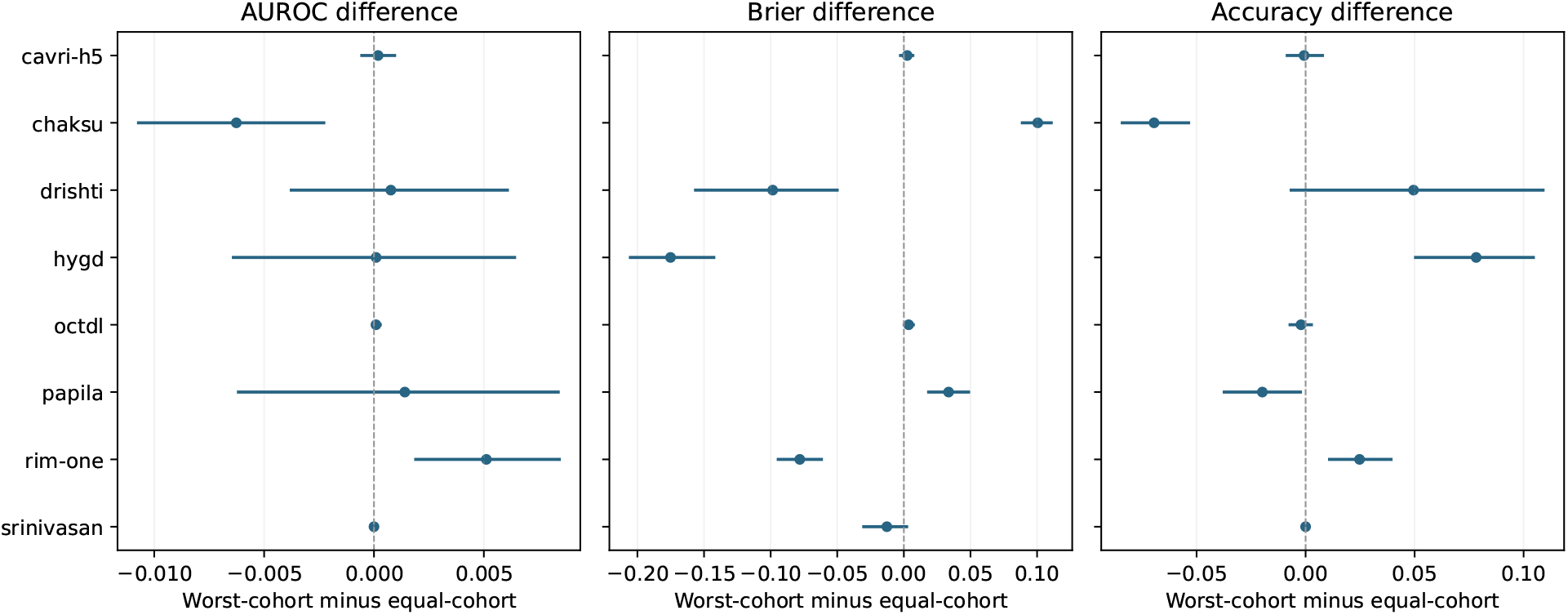
All cohort contrasts for maximum-cohort versus equal-cohort adaptation. Dots average paired differences across three seeds; bars are 95% intervals from 500 paired connected-group bootstrap draws. Lower Brier and higher AUROC or accuracy are favorable. Intervals are exploratory and not multiplicity-adjusted.

## 7 General-purpose visual Decision comparisons

These models receive the same stored 224-pixel ophthalmic images and closed candidate descriptions. Zero-shot supervision differs from every trained OphJev comparator. HYGD metrics use unique images; Srinivasan averages original four-class probabilities by subject before conditioning. OCTDL is conditional DME/normal; unsupported CAVRI labels are excluded. Table entries are AUROC / summed-class Brier. Inference uses the documented PyTorch compatibility path, not the vendor optimized runtime. Candidate-order, wording, no-image, and different-image interventions use the first 32 image identifiers by a fixed hash ordering per cohort. Outputs retain stable class keys; probability sensitivity does not by itself establish image grounding or correctness. The shuffled-image control may substitute an image from the same subject and is consequently a weak visual perturbation, not a patient-independent randomization.

**Table 39:** Complete eight-cohort zero-shot Decision matrix. No cohort is selected for presentation.

| Cohort | 0.8B | 2B | 4B |
| --- | --- | --- | --- |
| cavri-h5 | 0.8965 / 0.5465 | 0.9763 / 0.3936 | 0.9429 / 0.4421 |
| chaksu | 0.6265 / 0.3165 | 0.7731 / 0.4948 | 0.7365 / 0.4761 |
| drishti | 0.7055 / 0.5936 | 0.8387 / 0.3500 | 0.8364 / 0.3631 |
| hygd | 0.7118 / 0.7192 | 0.7409 / 0.5007 | 0.6536 / 0.5264 |
| octdl | 0.9891 / 0.1230 | 0.9929 / 0.0747 | 0.9902 / 0.0593 |
| papila | 0.4949 / 0.3876 | 0.7624 / 0.5232 | 0.7343 / 0.4826 |
| rim-one | 0.3824 / 0.4743 | 0.5824 / 0.5875 | 0.5043 / 0.5811 |
| srinivasan | 1.0000 / 0.3050 | 1.0000 / 0.2672 | 1.0000 / 0.1815 |

**Table 40:** Exploratory StartLux minus matched-adaptation DINOv2-B linear contrasts across all three reported metrics. The comparator has ophthalmic supervision; these are contextual differences, not isolated architectural effects. Intervals use 500 paired patient/pixel-cluster draws without multiplicity correction.

| Cohort | Size | Metric | Difference | 95% interval |
| --- | --- | --- | --- | --- |
| hygd | 0.8B | auroc | -0.0776 | [-0.1380, -0.0093] |
| hygd | 0.8B | brier | 0.0534 | [-0.0070, 0.1087] |
| hygd | 0.8B | accuracy | -0.2465 | [-0.2865, -0.2019] |
| hygd | 2B | auroc | -0.0485 | [-0.1030, 0.0081] |
| hygd | 2B | brier | -0.1652 | [-0.2301, -0.1026] |
| hygd | 2B | accuracy | 0.0208 | [-0.0246, 0.0719] |
| hygd | 4B | auroc | -0.1358 | [-0.1883, -0.0772] |
| hygd | 4B | brier | -0.1395 | [-0.2060, -0.0758] |
| hygd | 4B | accuracy | -0.0213 | [-0.0681, 0.0304] |
| srinivasan | 0.8B | auroc | 0.0000 | [0.0000, 0.0000] |
| srinivasan | 0.8B | brier | 0.2182 | [0.1596, 0.2912] |
| srinivasan | 0.8B | accuracy | -0.2333 | [-0.4000, -0.1000] |
| srinivasan | 2B | auroc | 0.0000 | [0.0000, 0.0000] |
| srinivasan | 2B | brier | 0.1805 | [0.0877, 0.2872] |
| srinivasan | 2B | accuracy | -0.2000 | [-0.3667, -0.0667] |
| srinivasan | 4B | auroc | 0.0000 | [0.0000, 0.0000] |
| srinivasan | 4B | brier | 0.0947 | [0.0279, 0.1747] |
| srinivasan | 4B | accuracy | -0.1333 | [-0.2667, -0.0333] |

**Table 41:** Complete presentation and image intervention probes on 32 hash-selected images per model and cohort. Columns report argmax changes after a paraphrase or reversed candidate order, and mean probability total variation after removing or substituting the image. These descriptive changes are not diagnostic performance.

| Cohort | Size | Wording flip | Order flip | No-image TV | Swap TV |
| --- | --- | --- | --- | --- | --- |
| cavri-h5 | 0.8B | 56.2% | 12.5% | 0.309 | 0.163 |
| cavri-h5 | 2B | 40.6% | 9.4% | 0.492 | 0.212 |
| cavri-h5 | 4B | 18.8% | 6.2% | 0.516 | 0.245 |
| chaksu | 0.8B | 0.0% | 0.0% | 0.201 | 0.025 |
| chaksu | 2B | 25.0% | 25.0% | 0.505 | 0.091 |
| chaksu | 4B | 31.2% | 21.9% | 0.514 | 0.096 |
| drishti | 0.8B | 0.0% | 6.2% | 0.249 | 0.028 |
| drishti | 2B | 12.5% | 18.8% | 0.579 | 0.110 |
| drishti | 4B | 18.8% | 3.1% | 0.596 | 0.117 |
| hygd | 0.8B | 0.0% | 0.0% | 0.161 | 0.026 |
| hygd | 2B | 12.5% | 31.2% | 0.409 | 0.080 |
| hygd | 4B | 18.8% | 9.4% | 0.414 | 0.098 |
| octdl | 0.8B | 62.5% | 6.2% | 0.309 | 0.182 |
| octdl | 2B | 21.9% | 6.2% | 0.622 | 0.292 |
| octdl | 4B | 12.5% | 6.2% | 0.615 | 0.292 |
| papila | 0.8B | 0.0% | 0.0% | 0.227 | 0.031 |
| papila | 2B | 15.6% | 34.4% | 0.521 | 0.115 |
| papila | 4B | 18.8% | 15.6% | 0.498 | 0.081 |
| rim-one | 0.8B | 0.0% | 3.1% | 0.232 | 0.034 |
| rim-one | 2B | 0.0% | 12.5% | 0.636 | 0.180 |
| rim-one | 4B | 6.2% | 9.4% | 0.569 | 0.193 |
| srinivasan | 0.8B | 37.5% | 3.1% | 0.294 | 0.114 |
| srinivasan | 2B | 31.2% | 12.5% | 0.473 | 0.198 |
| srinivasan | 4B | 12.5% | 6.2% | 0.464 | 0.211 |

Checkpoint revisions (0.8B, 2B, and 4B respectively):

bd4f76a600e23227547fee7bfc1825e12a32764c

3e2e456409a7fe44be69eee231566f7830872ea2

9302aedb7f7bd889994336f2ae919dffbf5dbee1

Weight downloads are checksum verified; full weights and native image data are not redistributed.

## 8 Materials and methods

### 8.1 Study design and scope

The unified generalist–specialist extension is described above and in the main Methods. The preceding study comprises a candidate-readout experiment, a separate target–loss experiment, a conventional-baseline and model-cascade analysis, a frozen evaluation on six external cohorts, and a subsequent fusion/adaptation extension with two newly acquired held-out cohorts. Each reports all specified arms across three seeds (2027, 2028, and 2029). The candidate experiment fixes its descriptions, training settings, and intervention definitions before evaluating its final fits. The data partitions had been inspected in earlier development, so this is an exploratory retrospective benchmark, not a prospectively registered confirmatory study. The original two experiments have different readout architectures and must not be pooled as a single matched comparison.

The unit of prediction is a fundus photograph or OCT image. There is no paired fundus–OCT training, no fusion of the two image types, no longitudinal clinical outcome prediction, and no new human reader study. OCT provides a second modality and candidate schema; only the fundus data support a reader-response analysis. The Srinivasan extension additionally aggregates image probabilities to subject-level diagnostic endpoints. The external OCT extension also assesses an unsupported category without training a new class.

### 8.2 Fundus images and glaucoma-screening targets

We use the locally available AIROGS photographs aligned with the released JustRAIGS annotations [8, 9]. Of 101,442 available photographs, 101,423 match the annotation table. The released final endpoint is non-referable versus referable glaucomatous optic neuropathy (NRG/RG), with 3,270 RG cases. NRG is a referral category and is not synonymous with a healthy eye or a normal optic disc. Candidate descriptions retain this distinction.

Let *y_i_* ∈ {0, 1} be the released final label and *r_i_*_1_*, r_i_*_2_ the G1/G2 reader labels. For the 96,786 cases where both readers give a binary NRG/RG label, the empirical reader target is

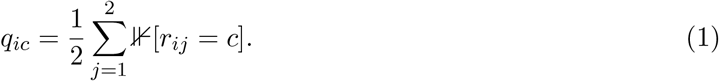

Missing and ungradable responses are excluded from this binary target rather than mapped to NRG. The selectively observed G3 adjudication is never counted as an exchangeable third reader. The released final label is used directly; we do not reconstruct the adjudication rule. Clinical metrics use the final label, whereas reader metrics use *q_i_* on the two-binary-reader mask. Matched target comparisons use this same eligible training mask for both supervision arms.

Fellow-eye links define connected groups for fundus partitioning. These are grouping proxies, not independently verified patient identities. Exact processed-image hashes are also checked; all matched fundus images have distinct hashes. No claim is made that hashing detects different views of the same eye or all near-duplicate images.

### 8.3 OCT images and duplicate-aware partitioning

We use the local 224-pixel OCTMNIST mirror, with CNV, diabetic macular edema (DME), drusen, and normal classes, derived from the public OCT classification resource [10, 11]. The mirror contains 109,309 rows: 97,477 training, 10,832 validation, and 1,000 test images. Our audit identifies 1,334 exact processed-image hash groups shared between the original training and validation partitions. Removing 1,535 training rows whose hashes occur in reserved validation or test data leaves 95,942 training candidates. No exact test–training or test–validation overlap is detected. The test set contains 975 distinct hashes and equal class counts.

Hash groups remain intact when training data are divided into fitting and development partitions and validation data into calibration and threshold-selection partitions. The fixed public test set is preserved. Training and validation are class-imbalanced, whereas the public test set is balanced. This prior shift can affect absolute calibration even without a wording change; our wording contrasts hold the test images fixed. We retain within-partition duplicates and label conflicts and account for test duplicates in uncertainty estimates. Patient identifiers are unavailable in the local mirror; image-hash isolation therefore must not be described as patient-disjoint evaluation. Existing analyses of leakage motivate this distinction [12].

### 8.4 Five disjoint roles for data

Each run stores the exact indices and sample identifiers for five group-isolated partitions: fitting, development, calibration, threshold selection, and test. Fitting updates the head and preprocessing statistics. Development selects the checkpoint. Calibration estimates probability mappings. Threshold selection defines a specificity operating point. Test data are used only to report that run’s performance. Pairwise group-overlap assertions are evaluated for every seed. For OCT, seeds change the internal group partitions and training randomness while leaving the test set fixed; these are not three independent clinical cohorts.

**Figure 8:**
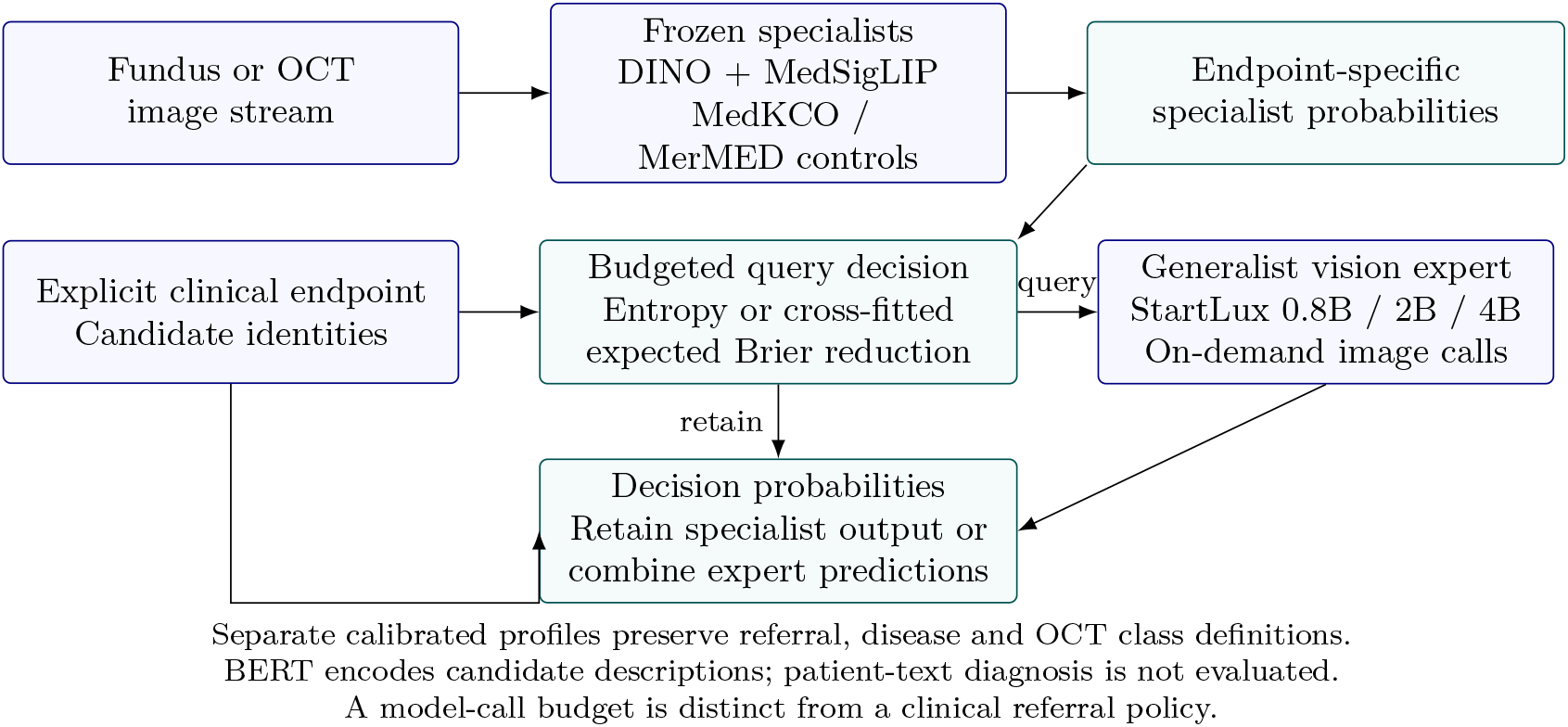
OphJev separates the candidate interface from the target of calibration. Frozen image and text representations feed a trainable shared candidate scorer. Candidate descriptions specify the closed label set; they are not clinical reports. Fundus probabilities are evaluated against either the released clinical label or the empirical G1/G2 response distribution, using separately fitted temperatures. OCT has only the clinical-label evaluation. Held-out wording, parameter-matched random identities, and input interventions examine which properties follow from language, architecture, or calibration. The routing row connects schema changes to actual model-upgrade workload and downstream referral outcomes; the gray row shows the original disjoint data roles. The final row summarizes the later fusion/adaptation extension, distinguishing inspected development cohorts, locked new-cohort evaluation, and post-hoc controls.

### 8.5 Representations and candidate scoring

The primary candidate experiment uses frozen DINOv2-B and MedSigLIP image representations. Additional backbone results, where included, use the same stored-image budget and are specified in the result tables. Images were first resized and center-cropped to 224 pixels. Some cached encoders subsequently resize these stored images to their expected input size; this does not recover native high-resolution detail. The study is a controlled readout comparison, not a claim to reproduce each foundation model’s best native-resolution performance. Per-encoder pooling, input size, feature dimension, and weight hashes are retained with the caches.

For image feature *v_i_* and candidate description *d_c_*, a frozen text tower gives *t_c_*. Trainable two-layer projections with LayerNorm map them to *h_i_* = *g_v_*(*v_i_*) and *u_c_* = *g_t_*(*t_c_*). A shared function scores each candidate:

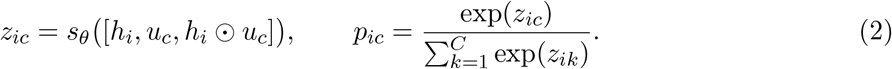

The options share parameters and differ through their inputs. The model outputs a normalized probability vector over the supplied closed candidate set. The question and candidate description are concatenated before text encoding; the evaluated question is fixed within each task. This protocol tests alternative descriptions of known classes, not general question answering, arbitrary new diseases, or unseen clinical tasks.

The fixed classifier predicts class-index logits from the image representation, using the same image projection and a hidden width chosen to match the candidate-head parameter budget within 0.1%. The nonsemantic identity control uses frozen random candidate vectors with the same dimension, projection, and shared scorer as the corresponding text arm. This strictly matches trainable capacity within each text-tower comparison while removing the language representation. MedSigLIP text and the FLAIR-adapted clinical BERT have different feature dimensions; comparisons across towers report that difference and use tower-specific random controls.

### 8.6 Wording protocol and mechanism controls

Each task has three training schemas and three held-out schemas, all listed in the supplement. Training rotates the available training wording. Held-out descriptions are never used in the training consistency term or checkpoint selection. NRG descriptions refer to the referral criterion; CNV descriptions retain choroidal neovascularization rather than equating it with all neovascular agerelated macular degeneration. The wording was constructed to preserve the endpoint, but has not undergone an independent masked clinical equivalence study.

We distinguish four interventions. A *permutation* changes candidate order and restores outputs by candidate identity before evaluation. A *mismatch* assigns a different class description to a fixed clinical key and intentionally does not restore the correct mapping. *Zero-text* gives every candidate the same zero vector. Finally, *held-out wording* changes descriptions while retaining class identity. Permutation tests equivariance, mismatch tests dependence on candidate assignment, and zero-text tests dependence on candidate-distinguishing input. None alone demonstrates language understanding. In particular, Equation 2 predicts identical logits and uniform probabilities when all candidate vectors are identical; this is a structural consequence, not an image-only classifier baseline.

For every candidate control, the input identity is permuted together with its associated key. Fixed-classifier predictions are joined to candidate keys without corrupting their clinical meaning. Thus a control is not penalized for an artificial output-column error. Generalization to held-out descriptions, relative to matched random identities, is the informative test of the language representation within this closed-label design.

### 8.7 Training objectives and consistency intervention

The primary candidate heads minimize distributional cross-entropy, using *q_i_* for eligible fundus training cases and one-hot labels for OCT:

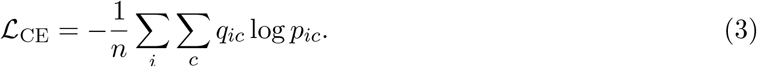

For consistency training, two training descriptions of the same candidate set produce *p*^(*a*)^ and *p*^(*b*)^.

The intervention adds a prespecified Jensen–Shannon penalty,

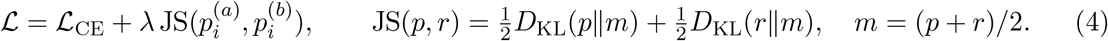

This encourages invariance across training wording; it provides no guarantee for unseen descriptions. Hyperparameters and exact implementation are recorded in the run protocol and Supplementary Methods. The visual and text backbones remain frozen. Consequently, the learned alignment is confined to the projection/scoring head and is not a new end-to-end retinal language pretraining result.

The separate target–loss experiment uses linear frozen-feature probes over DINOv2-S, RETFound-CFP, and RETFound-OCT representations of the same fundus images. It crosses final-label versus G1/G2 targets with cross-entropy alone versus cross-entropy plus 0.1 summed-class Brier loss, using three seeds and identical eligible training masks. All 36 selected checkpoints reach the fixed 30-epoch cap. These results describe this finite optimization budget, not converged equivalence or superiority of the objectives.

### 8.8 Calibration specifies both an endpoint and a schema

For logits *z*, temperature scaling produces *p_T_*= softmax(*z/T*) with positive scalar *T*, selected by calibration cross-entropy. A clinical temperature fits released hard labels. A reader temperature fits the empirical G1/G2 distribution on eligible calibration cases. These are different statistical objectives and their probabilities are labeled accordingly. The dedicated paired calibration-target analysis refits both temperatures on exactly the same eligible calibration cases and evaluates both on the same eligible test cases, isolating the target from sample composition.

Wording transfer first applies the *canonical* temperature unchanged to held-out descriptions. A separate *schema-specific recalibration* condition refits temperature on the same reserved calibration images and labels, now using logits from the new description. An additional affine condition fits a positive scalar slope and zero-sum class biases. Both use labeled calibration data for each new schema, although neither uses test labels or updates prediction weights. Their cost and information requirement distinguish them from zero-label wording robustness. A scalar temperature can alter confidence; it cannot reverse a wrong binary ranking or change multiclass argmax decisions. Affine class biases can change the operating point and multiclass decisions, so this stronger intervention is reported separately.

Clinical Brier loss sums over classes, ||*p*-*e_y_*||_2_^2^; in the binary task this equals twice the positive-class Brier score. Reader-distribution Brier loss is ||*p*-*q*||_2_^2^. Averaging one-hot Brier losses over the two observed readers gives

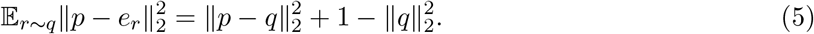

The second term is an observed ambiguity term independent of *p*. This algebraic identity does not require independence between readers and does not identify population-level uncertainty from two annotations.

### 8.9 Endpoints, operating points, and uncertainty

We report binary AUROC for fundus and macro one-versus-rest AUROC for OCT, together with average precision, accuracy, negative log-likelihood (NLL), summed-class Brier loss, and 15-bin equal-width top-confidence expected calibration error (ECE). Reader metrics comprise soft cross-entropy, summed-class Brier loss, and classwise ECE on the eligible mask. ECE depends on binning and prevalence, so proper-score results accompany it.

For fundus referral, a threshold is selected using only the threshold-selection partition to target at least 95% empirical specificity, then evaluated on test images without adjustment. We report the achieved test sensitivity and specificity, not a claim of exactly 95% test specificity. The local split and operating-point protocol differ from the official challenge evaluation, so these values are not a direct leaderboard comparison.

Results summarize every seed; standard deviations describe run-to-run variation rather than population confidence intervals. Paired uncertainty intervals resample fellow-eye proxy groups for fundus and exact-hash groups for OCT, preserving comparisons on the same test cases. These intervals condition on fitted models and calibration parameters. They do not account for model selection across projects, unknown patient clustering, pretraining overlap, or transport to another clinical population. Held-out wording results average all three specified descriptions; individual-schema results remain available.

Following the observed wording failures, we perform two explicitly post-hoc diagnostics. First, we compare the direction of the RG-minus-NRG contrast in frozen text space for each held-out description with the mean training contrast. Second, we flag a schema if any calibration-set one-versus-rest AUROC is below chance (binary: RG only), and then inspect its test performance. The 0.5 cutoff is fixed by chance discrimination rather than optimized on test labels. These analyses characterize failure modes and retrospective detectability; they do not provide an independently validated acceptance gate.

### 8.10 Conventional controls, actual model upgrades and timing

The follow-up analysis adds 18 linear probes over frozen DINOv2-B and MedSigLIP features, using the corresponding candidate runs’ eligible fitting mask, scaling, five partitions, optimizer settings, development-label checkpoint selection and calfit temperature scaling. Fundus probes compare final-label and G1/G2 distributional supervision. OCT uses one-hot supervision, for which the two targets coincide. These smaller-capacity controls are distinct from the parameter-matched fixed classifiers. Thirteen of 18 selected probes reach the 30-epoch cap; this is a matched-budget comparison, not an exhaustive optimization of linear models.

For model routing, DINOv2-S supplies a cheap probability vector *p_c_* and MedSigLIP supplies *p_s_*on exactly aligned cases. Both use the same head type, either fixed or MedSigLIP-text-conditioned. For an upgrade indicator *a*(*x*), the actual cascade prediction is

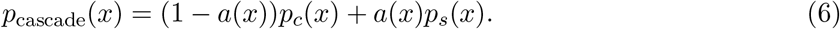

The strong model retains its real errors; neither a perfect model nor a hypothetical human is substituted. Policies score images by a stable label-independent random hash, predictive entropy, disease probability, or proximity to the cheap model’s clinical threshold. For each policy, calroute score quantiles define nominal upgrade budgets of 10, 20, 30 and 50%. Entropy at 30% is the designated descriptive primary setting; all settings are retained. A separate calroute threshold for each resulting mixture targets 95% specificity for RG. In OCT, the analogous abnormal-versus-normal endpoint is exploratory and is not a glaucoma referral decision. Both thresholds remain fixed on test images.

The cascade policies and budgets were recorded before computing their new test outcomes, after original single-model summaries had been inspected. These are exploratory follow-up analyses, not preregistered confirmation. Model upgrades and predicted clinical referrals are distinct events. Cost comparisons use observed test upgrade fractions, which can differ despite the same calibration budget. Corrected, introduced and shared argmax classification errors measure error complementarity; they are separate from referral false negatives. Paired cascade intervals use 1,000 group-bootstrap draws within each seed. OCT head comparisons use 2,000 shared hash-group resamples of the same test cases across three fitted seeds, avoiding artificial expansion to 3,000 independent images. These intervals condition on those particular trained models and are not multiplicity-adjusted.

The wording-routing intervention changes only the cheap candidate descriptions, retaining the strong model’s canonical interface. Canonical upgrade and clinical thresholds transfer unchanged. Separate conditions apply the old temperature or a schema-specific temperature or affine mapping fitted on calfit labels. This tests a fixed routing rule under schema change rather than selecting new test-set quantiles.

Single-image timing uses an RTX 5090, BF16 encoder inference, FP32 readouts, 20 warm-ups and 100 timed repetitions per method, with GPU synchronization and methods randomly interleaved within each encoder. It includes resident-image conversion, resizing, normalization, encoding, feature standardization, readout and calibrated softmax. It excludes disk reads, image decoding, host-to-device transfer, model loading and repeated text encoding; candidate embeddings are cached. Background jobs are recorded and not stopped. The component cost model is *C_c_* +*u*^*C_s_* for a cascade, using its actual test upgrade fraction *u*^, versus *C_s_* for the strong model alone. This descriptive estimate is separate from actual dynamic-cascade timing on 512 label-independently sampled fundus test images using seed 2027 models. Both encoders remain resident and the cheap model’s live entropy controls the upgrade branch. After 20 warm-ups, each of four paths is timed once per sampled image, in randomly interleaved order; this is distinct from the 100-repetition component benchmark.

### 8.11 Frozen transfer to six external cohorts

We extend the internal benchmark to six native public cohorts, reusing the existing checkpoints without external fitting, model selection, calibration, or threshold adjustment. DINOv2-S, DINOv2-B, and MedSigLIP yield 18 external feature matrices. The 420 model–cohort evaluations produce 500 three-seed summaries across heads, descriptions, and calibration states; these are repeated evaluations of existing fits, not 420 new training runs. The original training total remains 222. The main external comparison fixes the MedSigLIP visual backbone and contrasts its MedSigLIP candidate head with the matched fixed MLP and the conventional linear probe. All other completed conditions remain in the aggregate release.

#### Native endpoints

PAPILA provides bilateral photographs and clinical labels [13]. We retain 420 normal/glaucoma eyes from 210 patients for binary evaluation and hold 68 suspect eyes outside those primary metrics. Cháks.u supplies five image-based expert decisions for each of 1,345 photographs acquired with three cameras [14]. Its released decisions are normal versus glaucoma suspect, not independently confirmed clinical glaucoma. We use majority suspect status as the binary endpoint and the fraction of five suspect votes as a separate reader target. Removing seven exact-image repetitions leaves 1,338 images. RIM-ONE DL contributes 485 normal/glaucoma images and Drishti-GS contributes 101 [15, 16]. These native labels are related to, but not interchangeable with, the internal RG referral endpoint; we do not combine cohorts into one screening-performance estimate.

OCTDL contains additional retinal diagnoses that do not map unambiguously to the four internal classes [17]. Its DME/normal subset contains 474 unique images in 214 patient-plus-pixel components after deduplication. For discrimination, accuracy, and proper scores on this subset, we use the conditional probability *p*(DME)*/*[*p*(DME) + *p*(NORMAL)]. We additionally report unchanged four-class argmax accuracy on these images and probability mass assigned outside DME/normal. AMD images are not relabeled CNV or drusen. The frozen OCT abnormality threshold always uses the original four-class 1 − *p*(NORMAL) score, including for OCTDL; it is distinct from the conditional DME score.

CAVRI-H5 supplies 609 OCT images with CNV, DME, drusen, normal, and vitreomacular adhesion/traction (VMA–VMT) labels [18]. The matching four-class subset comprises 96 CNV, 99 DME, 137 drusen, and 142 normal images (*n* = 474); we preserve the original four-column probabilities throughout this analysis. The additional 135 VMA–VMT images form an unsupported-class stress test, not a fifth trained class. We compare their entropy with that of the 474 modeled-class images and apply the 95th-percentile entropy threshold from the original internal threshold-selection set without modification. This diagnostic tests whether unsupported inputs receive greater uncertainty; it does not train or validate a disease detector. The source describes one image per person and selected, diagnostically clear central scans, but the local archive supplies no patient-identity table. We therefore label its uncertainty estimates as image-level, not verified patient-level, intervals.

#### Integrity, grouping, and uncertainty

External images are decoded and checked against the internal source images by exact RGB hashes; no exact intersection is found. Within each external collection, the first manifest occurrence of each identical image is retained without consulting predictions. Patient identifiers and identical-pixel links jointly define resampling components where identities are supplied. OCTDL contains duplicates spanning different native patient identifiers, making this joint rule necessary. PAPILA and OCTDL use 210 and 214 such components, respectively; the other cohorts lack locally verified patient linkage. Hash absence cannot exclude recompressed or transformed copies, and image-level intervals may be too narrow if unrecorded within-person dependence remains.

All 609 CAVRI-H5 RGB hashes are distinct. Two drusen files in different provider partitions share a basename but have different pixels. We retain both images and repeat primary paired comparisons with this basename pair grouped, reducing the known-class resampling units from 474 to 473. The point estimates and all primary interval-direction conclusions are unchanged. The provider’s train/validation names do not define an OphJev training split: every CAVRI image is evaluated only after freezing the internal models.

External paired percentile intervals use 500 bootstrap draws, with the same sampled components applied to each comparator and all three seed fits. The statistic is the mean of the three per-seed metrics, not a probability ensemble. Intervals condition on the fitted checkpoints and calibrators; the exploratory matrix has no multiplicity adjustment. We also transfer 216 rows of existing DINOv2-S–MedSigLIP cascade conditions across the six cohorts. Upgrade thresholds, referral/abnormality thresholds, and canonical or schema-specific temperatures all come from internal data. In particular, the nominal 30% internal upgrade budget is not reset using external quantiles.

#### Pretraining exposure

Externality here refers to OphJev head training and internal calibration. A pinned audit of FLAIR’s preparation recipe lists PAPILA and Cháks.u in pretraining [19]. Accordingly, results using its pretrained BERT candidate vectors cannot be called backbone-unseen evaluation. The same directory’s fixed and random-identity controls do not consume pretrained FLAIR text vectors. Other public encoders also lack image-level certificates of nonexposure. These qualifications remain material even when exact overlap with our downstream training images is absent.

### 8.12 Source-only fusion of complementary frozen readouts

After inspecting the first six-cohort results, we evaluated whether complementary encoders and conventional heads could improve the candidate interface. This extension is exploratory method development. The six experts are MedSigLIP and DINOv2-B visual encoders, each paired with a hard-label linear probe, fixed MLP, or canonical MedSigLIP candidate-text head. Existing checkpoints remain frozen; no additional visual or language backbone is trained. The original 222 head fits are therefore reported separately from the small fusion and calibration fits below. Fusion uses canonical candidates only and does not establish robustness to the wording failures studied above.

Within each task and seed, a deterministic group hash splits the original development partition into 60% meta-fit groups and 40% meta-selection groups. These groups are separate from calibration and test groups, but the full development partition had already selected the base checkpoints. Reuse is disclosed rather than characterized as independent stacking validation. Expert temperatures *T_m_* minimize meta-fit NLL. Let *p_im_*= softmax(*z_im_/T_m_*) and *w* ∈ Δ^5^, where Δ^5^ is the nonnegative six-expert simplex. We compare the source-selected best single expert, uniform averaging, a convex mixture, an anchored mixture, and class-balanced fitting. Learned weights minimize meta-fit NLL plus 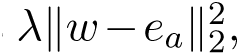 where *a* is the single expert with lowest meta-selection NLL and *λ* ∈ {0, 0.01, 0.1, 1}. The anchored arm additionally requires *w_a_* ≥ 0.5. Class-balanced fitting and its regularization selection use inverse class-frequency weights; other arms use ordinary mean NLL.

The regularization value for each arm is selected on meta-selection loss. A single fusion family is then selected by mean meta-selection NLL relative to the single-expert anchor across both tasks and three seeds, requiring no mean NLL worsening for either task. This source-only rule selects the anchored arm; it does not inspect new-cohort results. Each mixture receives a final positive scalar temperature *T*_src_ from the original calibration-fit partition. Referral/abnormality and entropy thresholds come from the original calibration-for-routing partition. Every family remains reported. The source-selected, newly calibrated single expert is distinct from the original v1 baselines, which retain their original calibration states in all final comparisons.

### 8.13 Endpoint-aware adaptation across development cohorts

For the next stage, the six already inspected external cohorts are explicitly designated development resources. Their original frozen-transfer results remain separate from the new adaptation analysis. Starting from the source-only anchored weights *w*_src_, we fit a simplex mixture and bounded class intercepts:

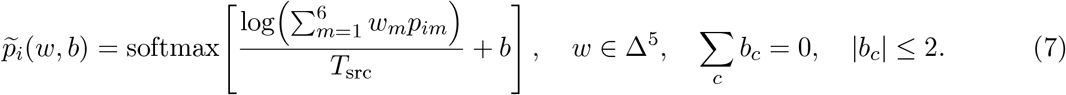

Eexpert and source-mixture temperatures remain frozen. Cohorts contribute equal total weight, and connected patient/pixel groups contribute equal total weight within each cohort. The objective adds fixed penalties 0.05||*w*-*w*_src_||*b*||_2_^2^ +0.01||*b*||_2_^2^ to this weighted NLL. Intercepts are fitted calibration parameters; they do not separately identify disease prevalence or causal clinical priors. SLSQP uses analytic gradients, a stable log-sum-exp likelihood, and the derivative of the declared probability floor. Numerical checks include partial-class likelihoods and extreme probabilities. Optimizer convergence does not establish a global optimum of the joint objective.

The source development partition remains in each fit. Fundus adaptation additionally uses PAPILA, RIM-ONE DL, and Drishti-GS clinical glaucoma labels. Cháks.u’s suspect labels never train this clinical adapter and remain a distinct transfer endpoint. This is supervised adaptation from referral grading toward diagnosis-oriented datasets, not an assumption that their labels are equivalent. OCT adaptation uses CAVRI-H5’s four modeled classes and OCTDL’s DME/normal subset. For OCTDL, the loss is conditional on the supported class pair,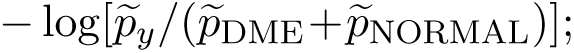 unsupported classes incur no fabricated label penalty. VMA–VMT examples never enter supervised fitting.

For every old-cohort report, that entire cohort is excluded from adapter fitting; no method or hyperparameter is selected from these leave-one-cohort-out (LOCO) outcomes. The earlier inspection of these cohorts still prevents calling LOCO a fresh blind validation. Cháks.u is always excluded, so its adapter equals the corresponding fit-all clinical adapter. Joint fusion and intercept adaptation is the prespecified primary arm. A bias-only arm holds *w* = *w*_src_, and the unchanged source ensemble is retained. There are 24 task/seed/fold states, comprising 18 LOCO and six fit-all states, with two learned adapters per state. All operating and entropy thresholds use source calibration-for-routing images; none uses the reported cohort. A separate source-test regression audit reports whether clinical adaptation damages the original screening task, without selecting a replacement state. Fit-all states are locked before the first HYGD or Srinivasan predictions are generated.

### 8.14 Two newly acquired held-out cohorts and patient-aware endpoints

HYGD version 1.1.0 provides 747 fundus photographs from 288 native patient identifiers with clinically assigned glaucomatous optic neuropathy (GON) labels [20–22]. Its GON-negative label does not establish that an eye is otherwise healthy. Labels reflect clinical assessment rather than solely photographic suspicion. No laterality is supplied. Exact-pixel deduplication retains 737 images representing 286 native identifiers; cross-identifier image copies yield 283 patient-plus-pixel correlation components. We evaluate native image labels using both ordinary image weights and inverse retained-image-count weights per native patient. The latter is a weighted pooled metric, not a mean of within-patient AUROCs. We do not average diagnoses across a conservatively linked component or invent eye identities. All eligible images remain included; machine-generated quality scores do not select cases or enter the ensemble.

The Srinivasan/Duke OCT release contains 3,231 B-scans from 45 volumetric subjects: 15 each with DME, dry AMD, and normal findings [23]. The primary endpoint is DME versus normal in 30 subjects. Within each subject, we average unchanged four-class probability vectors across exact-pixel-unique B-scans and only then condition on DME/normal. This differs from averaging already conditioned slice probabilities. Dry AMD is not relabeled CNV or drusen. We also report original four-class argmax and off-subset probability mass, pooled slice metrics with ordinary or equal-patient weights, and an exploratory 45-subject any-pathology endpoint using 1 – *p^_^*_NORMAL_. Slice labels inherit the source volume diagnosis. Thresholds transferred from individual source images are not assumed calibrated for volume averages.

Before inference, all new images undergo full decoding, identity alignment, and exact-pixel checks against the original source and six development cohorts. New-cohort predictions require matching frozen model, protocol, and script hashes. An immutable first-evaluation receipt and per-seed prediction receipts document the sequence. Neither new cohort contributes labels or predictions to model selection, adapter fitting, calibration, thresholds, or aggregation selection. This establishes separation from downstream optimization, not certified absence from foundation-model pretraining. Primary comparisons retain original MedSigLIP text, fixed, and matched linear heads alongside the source-only anchored ensemble. DINOv2-B experts, the additional hard-label fundus linear probe, and bias-only adaptation are also reported. We compute paired 500-draw percentile intervals by diagnosis-stratified resampling of patient-plus-pixel components, using the same draw across comparators and all three fixed seed fits. HYGD’s primary statistic uses unique images; Srinivasan’s uses subject-mean probabilities. Metrics are averaged across seeds rather than ensembling seed predictions. Intervals condition on the fixed models and observed diagnostic strata, have no multiplicity correction, and cannot remove uncertainty from the small 30-subject primary OCT sample. Argmax accuracy is kept separate from accuracy, sensitivity, and specificity at each method’s frozen source operating threshold.

#### Post-hoc controls with matched adaptation supervision

After inspecting the locked primary new-cohort outcomes, we fit a separate control grid to distinguish fusion from access to additional labeled development data. Each of the six single experts receives the same permitted cohorts, group weights, conditional/full endpoint loss, bounded zero-sum intercepts, and intercept penalty as the primary adapter. Expert temperatures and the common source-mixture temperature follow the frozen convention; mixture weights are fixed to the selected single component. This adds 144 small intercept fits across six experts and 24 task/seed/fold states. No new-cohort labels enter these fits, and the original primary model and predictions remain unchanged. These are post-hoc component controls, not a newly blinded comparison or an exhaustive search for each expert’s best calibration. Their full LOCO and new-cohort results, paired intervals, and source regressions are retained.

**Table 42:**
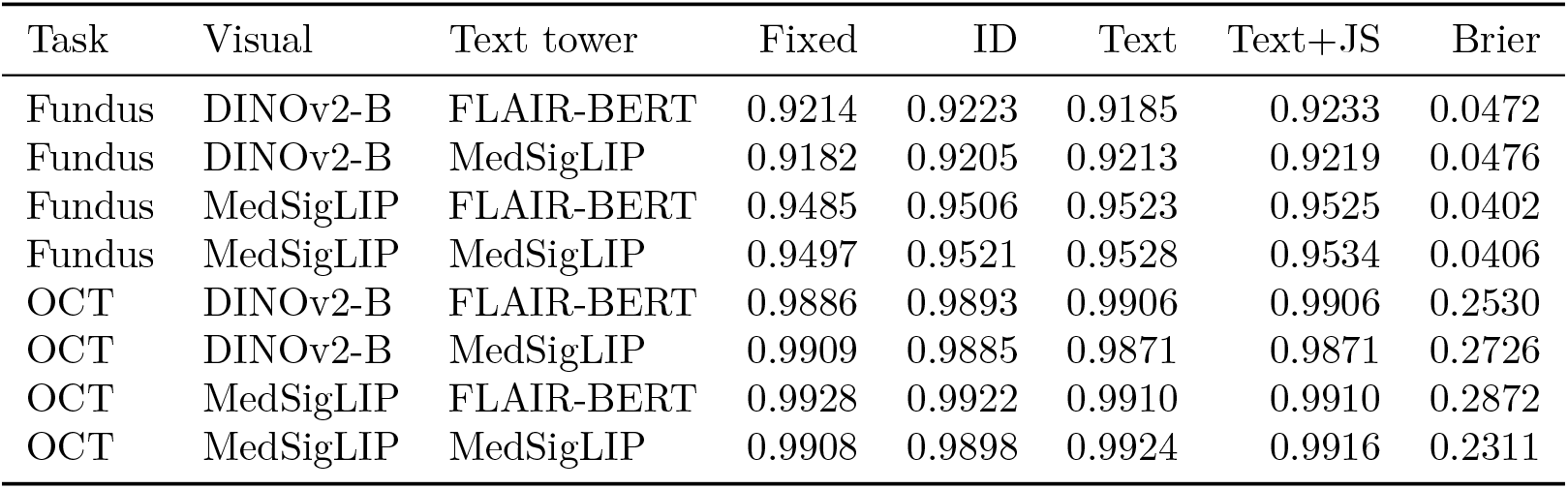
Primary comparison. AUROC is the mean of three seeds, under canonical wording and clinical temperature calibration. Fixed heads approximately match the parameter budget; random-ID and text heads match exactly within a text tower. Brier is reported for the text arm.

## 9 Results

### 9.1 Candidate readouts across two imaging modalities

The revised candidate experiment completed 168 fits: 96 in the two-visual-encoder, two-text-tower primary matrix and 72 in three additional encoder settings per modality with MedSigLIP text. Together with the separate 36 target–loss fits, this yields 204 fitted models reported under their respective protocols. The same fit evaluated under several schemas is counted once. In the primary matrix, the text head exceeded its parameter-matched random-ID control in mean canonical AUROC in 5 of 8 modality–visual-encoder–text-tower combinations; this is a descriptive count, not a significance test. Table 42 reports all primary comparisons, and the supplement reports every replication setting. Canonical classification performance alone does not establish that the text representation adds reliable wording transfer.

### 9.2 Absolute performance and conventional alternatives

With MedSigLIP vision and text, the candidate head attains mean fundus AUROC 0.9528 and OCT macro AUROC 0.9924. These are useful discrimination values, but do not establish clinical or cross-paper superiority. A constant fitting-label-prior baseline has fundus accuracy about 96.8%, AUROC 0.5 and AP about 0.032, illustrating why high accuracy alone is insufficient in this imbalanced task. A reader-supervised linear MedSigLIP probe attains fundus AUROC 0.9576, exceeding candidate text in every seed. At the calroute-selected operating point, candidate text and the matched fixed head have similar fundus sensitivity (76.75% versus 76.84%) and test specificity (94.62% versus 94.85%). False negatives are 7.51 and 7.47 per 1,000 images. Candidate conditioning does not improve this referral endpoint.

OCT provides a more favorable but conditional comparison. With MedSigLIP vision, candidate text reaches 85.03% accuracy and Brier 0.2311, versus 82.90% and 0.2632 for the matched fixed head, and 80.40% and 0.2988 for the linear probe. Mean paired accuracy differences are 2.13 percentage points (95% conditional CI, 1.23–3.12) and 4.63 points (3.00–6.32), respectively. Against the fixed head, the Brier difference is −0.0321 (−0.0420 to −0.0229). However, seed 2027 favors the fixed head by 1.8 accuracy points, and DINOv2-B does not reproduce the same language advantage. This supports a backbone-dependent mean improvement for these fitted models, not superiority on every retraining. AUROC near 0.99 should not be confused with near-perfect four-class accuracy. Table 43 includes all conventional and candidate controls with MedSigLIP text.

**Table 43:** Conventional controls under the same feature, split and optimization budget. Mean of three seeds; AP is average precision. Fundus accuracy alone is misleading because the RG prevalence is about 3.2%. OCT AP and AUROC are macro one-versus-rest. Linear hard and reader targets coincide in OCT. All candidate rows use MedSigLIP text.

| Task | Vision | Head | AUROC | AP | Accuracy | Brier |
| --- | --- | --- | --- | --- | --- | --- |
| Fundus | DINO-B | Matched fixed | 0.9182 | 0.4357 | 97.08% | 0.0469 |
| Fundus | DINO-B | Random candidate ID | 0.9205 | 0.4390 | 97.05% | 0.0471 |
| Fundus | DINO-B | Linear / final label | 0.9195 | 0.4691 | 97.14% | 0.0467 |
| Fundus | DINO-B | Linear / G1-G2 | 0.9235 | 0.4703 | 97.21% | 0.0453 |
| Fundus | DINO-B | Candidate text | 0.9213 | 0.4417 | 96.98% | 0.0476 |
| Fundus | DINO-B | Candidate text + JS | 0.9219 | 0.4491 | 97.09% | 0.0466 |
| Fundus | MedSigLIP | Matched fixed | 0.9497 | 0.5561 | 97.37% | 0.0404 |
| Fundus | MedSigLIP | Random candidate ID | 0.9521 | 0.5619 | 97.42% | 0.0401 |
| Fundus | MedSigLIP | Linear / final label | 0.9536 | 0.5939 | 97.49% | 0.0388 |
| Fundus | MedSigLIP | Linear / G1-G2 | 0.9576 | 0.5883 | 97.42% | 0.0396 |
| Fundus | MedSigLIP | Candidate text | 0.9528 | 0.5610 | 97.42% | 0.0406 |
| Fundus | MedSigLIP | Candidate text + JS | 0.9534 | 0.5683 | 97.36% | 0.0401 |
| Oct | DINO-B | Matched fixed | 0.9909 | 0.9767 | 83.23% | 0.2458 |
| Oct | DINO-B | Random candidate ID | 0.9885 | 0.9709 | 81.87% | 0.2690 |
| Oct | DINO-B | Linear / final label | 0.9904 | 0.9748 | 82.17% | 0.2531 |
| Oct | DINO-B | Candidate text | 0.9871 | 0.9685 | 81.83% | 0.2726 |
| Oct | DINO-B | Candidate text + JS | 0.9871 | 0.9684 | 81.87% | 0.2727 |
| Oct | MedSigLIP | Matched fixed | 0.9908 | 0.9768 | 82.90% | 0.2632 |
| Oct | MedSigLIP | Random candidate ID | 0.9898 | 0.9725 | 82.83% | 0.2645 |
| Oct | MedSigLIP | Linear / final label | 0.9918 | 0.9794 | 80.40% | 0.2988 |
| Oct | MedSigLIP | Candidate text | 0.9924 | 0.9795 | 85.03% | 0.2311 |
| Oct | MedSigLIP | Candidate text + JS | 0.9916 | 0.9773 | 83.53% | 0.2549 |

### 9.3 Wording transfer and calibration interventions

Fundus with DINOv2-B and MedSigLIP text changed from AUROC 0.9213 to 0.9210, ECE 0.0032 to 0.1205, and Brier 0.0476 to 0.0917 when the canonical calibrator was transferred to held-out wording. Fundus with MedSigLIP and MedSigLIP text changed from AUROC 0.9528 to 0.9527, ECE 0.0035 to 0.1033, and Brier 0.0406 to 0.0747 when the canonical calibrator was transferred to held-out wording. OCT with DINOv2-B and MedSigLIP text changed from AUROC 0.9871 to 0.9844, ECE 0.0957 to 0.0934, and Brier 0.2726 to 0.2872 when the canonical calibrator was transferred to held-out wording. OCT with MedSigLIP and MedSigLIP text changed from AUROC 0.9924 to 0.9898, ECE 0.0717 to 0.0703, and Brier 0.2311 to 0.2522 when the canonical calibrator was transferred to held-out wording.

Across all eight primary combinations, training-template consistency lowered mean held-out Brier in 3/8, schema-specific temperature in 4/8, and schema-specific affine calibration in 8/8 relative to the corresponding uncorrected text arm. These aggregate counts do not imply uniform improvement for each seed or description. In all 4 of the four primary OCT settings, refitted temperature increased mean held-out Brier; the calibration-to-test class-prior shift helps explain why fitting the reserved calibration distribution need not improve test calibration. For DINOv2-B/FLAIR-BERT fundus decisions, affine recalibration lowered mean held-out Brier from 0.2993 to 0.0516, but mean AUROC remained 0.6396; it did not recover the reversed ranking. The complete metric table includes both successful and unsuccessful interventions. The new-schema calibrators use labeled calibration cases and therefore are not zero-label fixes. Paired per-seed intervals average all three held-out descriptions within each test image before group resampling. All 168 fits passed the candidate-key permutation check, including the correctly handled controls (maximum absolute logit difference 0). Text removal produced uniform probabilities for every shared text scorer, exactly as predicted by its architecture. Mismatched descriptions were evaluated with the clinical keys held fixed; their results are provided in the aggregate intervention file. These checks establish correct handling and dependence of the input path; they do not identify general language understanding. Fresh CPU reconstruction of every saved checkpoint reproduced its sampled CUDA logits within a maximum absolute difference of 5.72e-06.

**Figure 9:**
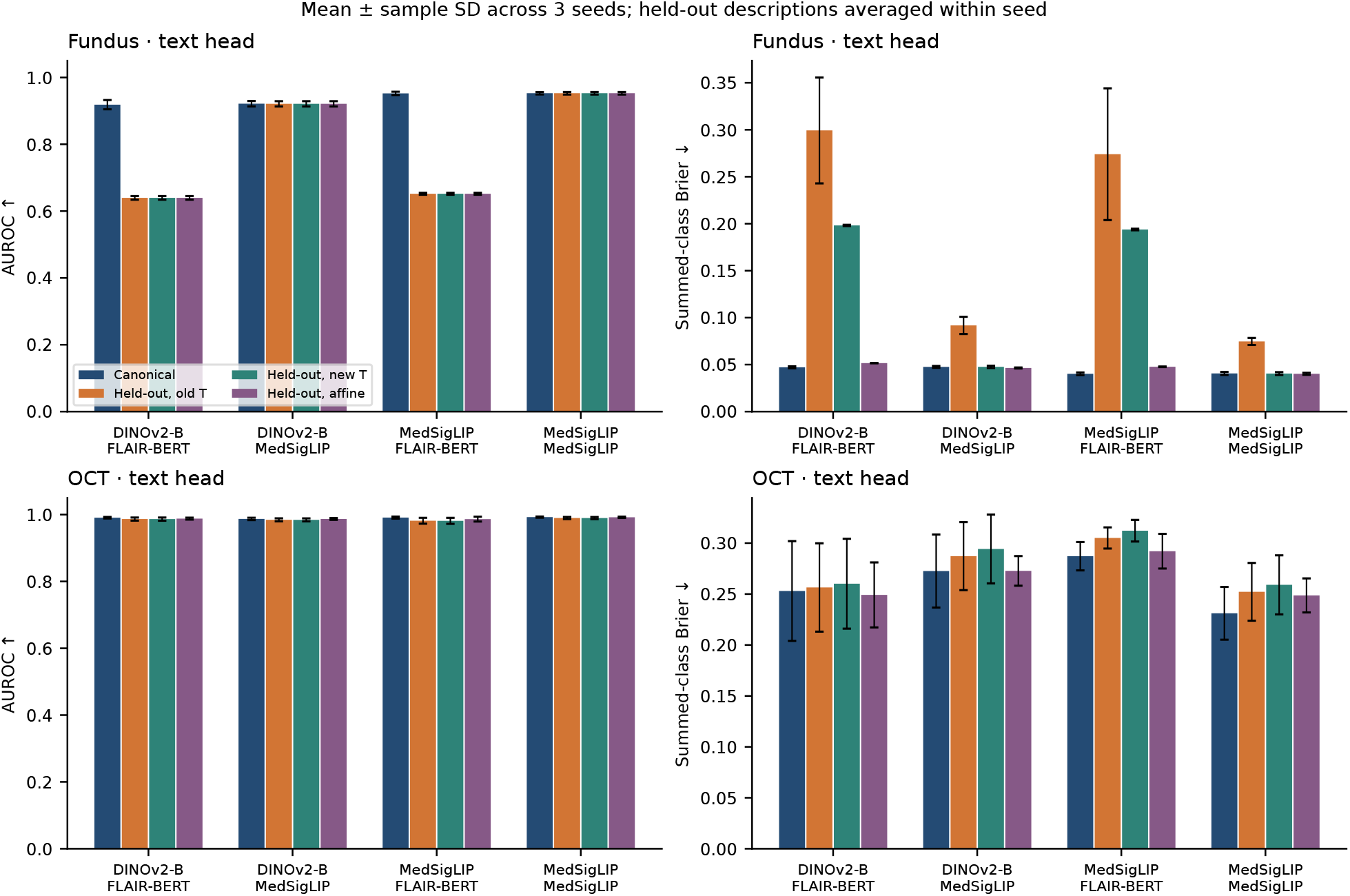
Wording transfer is evaluated separately from discrimination. Results use all three held-out descriptions and all three seeds. Canonical calibration is transferred unchanged unless schema-specific recalibration is indicated. Error bars and the precise aggregation unit are specified on the panels. A reduction in ECE without a corresponding proper-score improvement should not be interpreted as a comprehensive reliability gain.

### 9.4 Probability drift and referral-polarity reversal are distinct failures

With DINOv2-B images, the FLAIR-BERT-conditioned head changed from canonical AUROC 0.9185 to 0.0825 for held-out description 2. With MedSigLIP images, the FLAIR-BERT-conditioned head changed from canonical AUROC 0.9523 to 0.0513 for held-out description 2. The other two descriptions retained the class ordering much more closely. The reversal occurred in all six primary fundus text fits (two image encoders, three seeds) and persisted in their consistency-trained counterparts. This is a failure of the tested BERT-conditioned readout under that description, not an evaluation of the complete native FLAIR classifier or of all BERT models.

An exploratory representation analysis found a cosine of-0.121 between the failing description’s RG-minus-NRG vector and the mean training contrast in frozen FLAIR text space (Figure 10). The other two BERT descriptions and all three MedSigLIP descriptions had positive cosines to their respective mean training contrasts. This association suggests a candidate-polarity mechanism but does not isolate causality through the nonlinear trained scorer. Positive-slope recalibration cannot reverse a binary ranking, so a small calibration error after collapsing predictions toward prevalence would not constitute recovery of discrimination.

**Figure 10:**
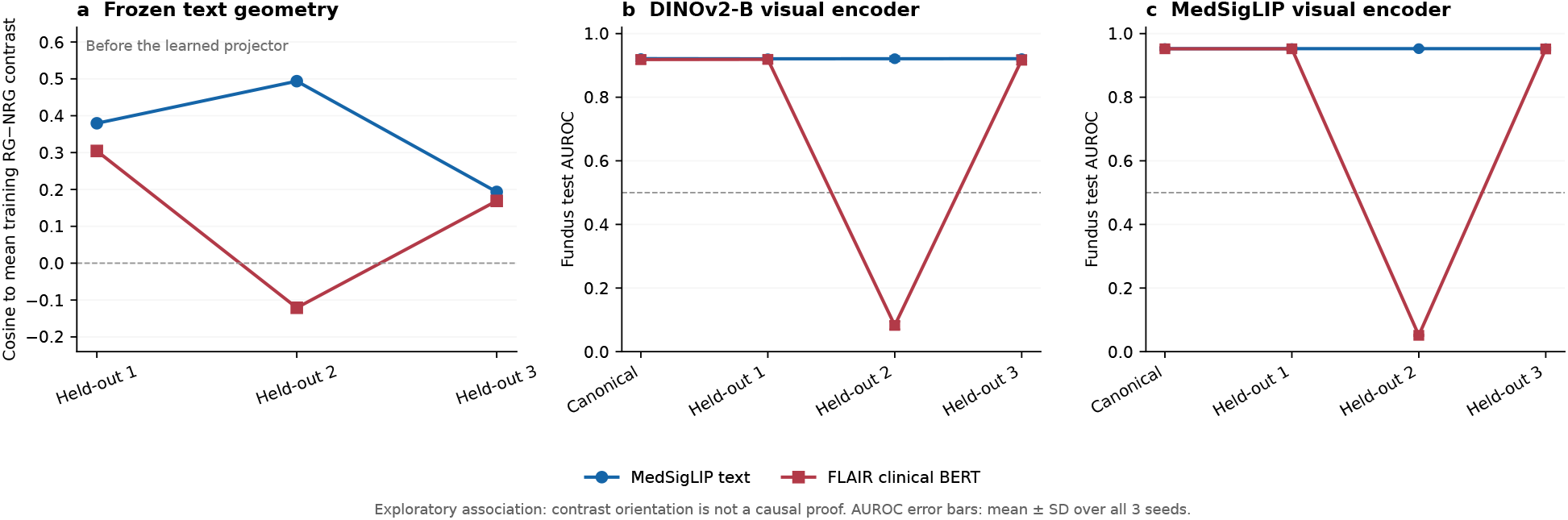
Exploratory diagnosis of a wording-specific polarity reversal. Left: cosine between each frozen RG-minus-NRG candidate contrast and the mean training contrast, computed separately for each text tower. Right: canonical and individual held-out AUROC for the trained fundus text heads, summarized over three seeds. The negative frozen-text contrast coincides with a reversal of the BERT-conditioned readout for description 2. This association does not establish causation through the nonlinear scorer. All three held-out descriptions are shown; none was changed after inspecting results.

A post-hoc check using only calibration labels flagged 12 of 192 primary text-head/schema evaluations for a below-chance class AUROC; 12 of these also showed the direction failure on the test partition. All flags concerned the same second held-out fundus description with the BERT tower, including both text training arms. This demonstrates retrospective detectability in the available calibration set; it is not a validated clinical rejection policy or an independent test of a preregistered detector.

### 9.5 Reader targets and proper-score objectives

In the separate 36-fit linear-probe experiment, G1/G2 supervision lowered the mean clinical Brier score on all three encoders (Table 44). Adding the 0.1 Brier term improved neither target arm in any of the nine encoder–seed settings. All selected checkpoints reached epoch 30, so this is a finite-budget result. The paired target effect was not uniformly distinguishable from zero for every encoder and seed; full intervals accompany the aggregate data.

Refitting both temperatures on the same two-binary-reader calibration cases isolates a change in calibration target from final labels to the empirical reader distribution. Reader Brier loss decreased for all nine saved G1/G2-supervised probes. Mean reader Brier changes were-0.001237,-0.001105, and-0.000547 for DINOv2-S, RETFound-CFP, and RETFound-OCT, respectively. On the same eligible test cases, mean clinical Brier changed by-0.000069,-0.000078, and +0.000348. The first two clinical changes are small; the intervals in Figure 11 are needed to judge their uncertainty. Thus an improvement against the reader distribution cannot be substituted for an improvement against the referral endpoint.

### 9.6 Referral operating points and reproducibility

DINOv2-B/MedSigLIP achieved mean sensitivity 0.640 at test specificity 0.949; applying the same canonical threshold unchanged to held-out descriptions gave mean specificity 0.398. DINOv2-B/FLAIR-BERT achieved mean sensitivity 0.628 at test specificity 0.951; applying the same canonical threshold unchanged to held-out descriptions gave mean specificity 0.541. MedSigLIP/MedSigLIP achieved mean sensitivity 0.768 at test specificity 0.946; applying the same canonical threshold unchanged to held-out descriptions gave mean specificity 0.505. MedSigLIP/FLAIR-BERT achieved mean sensitivity 0.772 at test specificity 0.948; applying the same canonical threshold unchanged to held-out descriptions gave mean specificity 0.615. These comparisons expose threshold transport separately from reselecting a threshold on new-schema calibration data. They are internal operating points and cannot be compared directly with the official JustRAIGS challenge leaderboard. Every new candidate fit retains its state dictionary, feature-scaling statistics, selected epoch, training history, calibration states, exact partitions, schema embeddings, and prediction arrays. Source and feature checksums identify the artifacts used in each run. Reproduction checks test group separation, candidate permutation, zero-text symmetry, and checkpoint replay. Resource measurements describe these cached-feature jobs and do not imply end-to-end clinical inference latency.

**Figure 11:**
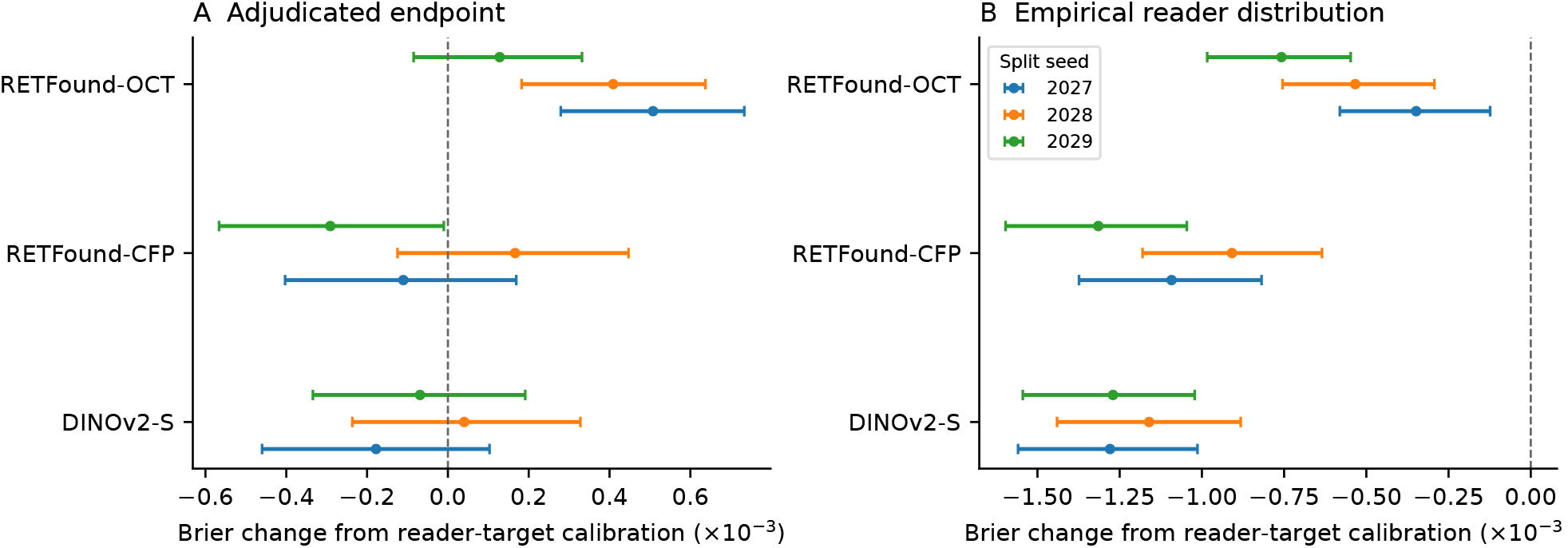
Changing the calibration target changes what a probability estimates. The same saved G1/G2-supervised linear probe is calibrated to either the final clinical endpoint or the reader distribution. Both temperatures are refitted on the same two-binary-reader calibration cases and evaluated on the same eligible test cases. Points show reader-target minus clinical-target calibration, with 2,000-draw paired fellow-eye cluster bootstrap intervals for each fixed model and seed. Negative values favor reader-target calibration for the indicated score. These are conditional, exploratory intervals, not independent-cohort confirmation.

**Table 44:** Separate target–loss study: mean clinical summed-class Brier over three seeds, evaluated on all clinical test cases. All probes use fundus images and the same eligible training mask. The final row uses an OCT-pretrained encoder on fundus images.

| Encoder | Final CE | Final CE+Brier | G1/G2 CE | G1/G2 CE+Brier |
| --- | --- | --- | --- | --- |
| DINOv2-S | 0.051368 | 0.051501 | 0.050503 | 0.050536 |
| RETFound-CFP | 0.054137 | 0.054270 | 0.053399 | 0.053511 |
| RETFound-OCT | 0.060359 | 0.060407 | 0.060169 | 0.060195 |

### 9.7 Model upgrades recover errors but retain a clinical trade-off

In the fundus text-head cascade, the 30% entropy budget upgrades 30.13% of test images. Sensitivity is 75.48%, versus 57.58% for DINOv2-S alone, 64.32% for random upgrades under the same calroute budget, and 76.75% for MedSigLIP alone. Test specificities are 94.69%, 94.69%, 94.67% and 94.62%, respectively. Compared with always using MedSigLIP, the entropy cascade adds 0.41 false negatives per 1,000 images (7.92 versus 7.51). No noninferiority margin was prespecified; this is not evidence of clinical equivalence. At the 50% budget, actual upgrades are 50.51%, sensitivity is 76.50%, and false negatives are 7.59 per 1,000 images. Table 45 reports the false-positive load.

For the binary fundus task, entropy and disease-score policies at the 30% and 50% budgets select identical cases in these runs; they are not independent confirmations. Fixed-head cascades, every budget, paired intervals and classification-error accounting are retained in the numerical release. In OCT, entropy at the nominal 30% budget actually upgrades 50.17% of images and yields 84.70% accuracy versus 85.03% for the strong model. Random routing upgrades 29.20%, so the two policies are not matched in actual test compute. The budget deviation accompanies the known validation– test composition shift; this experiment does not isolate a purely prevalence-driven causal effect.

**Table 45:** Actual model upgrades on the fundus test set. Mean of three seeds using MedSigLIP text for both encoders; nominal budgets are set on calroute. Each mixture selects its referral threshold on calroute to target 95% specificity; test specificity is reported. FN and FP are per 1,000 images. Cost is a component-based estimate, not measured cascade wall time.

| Policy (budget) | Upgrade | Sens. | Spec. | FN | FP | Cost ratio |
| --- | --- | --- | --- | --- | --- | --- |
| DINO-S only | 0.0% | 57.58% | 94.69% | 13.70 | 51.36 | 0.285 |
| Random (30%) | 29.8% | 64.32% | 94.67% | 11.52 | 51.62 | 0.582 |
| Entropy (30%) | 30.1% | 75.48% | 94.69% | 7.92 | 51.34 | 0.586 |
| Clinical boundary (30%) | 30.3% | 74.90% | 94.70% | 8.10 | 51.28 | 0.588 |
| Entropy (50%) | 50.5% | 76.50% | 94.65% | 7.59 | 51.74 | 0.790 |
| MedSigLIP only | 100.0% | 76.75% | 94.62% | 7.51 | 52.03 | 1.000 |

**Figure 12:**
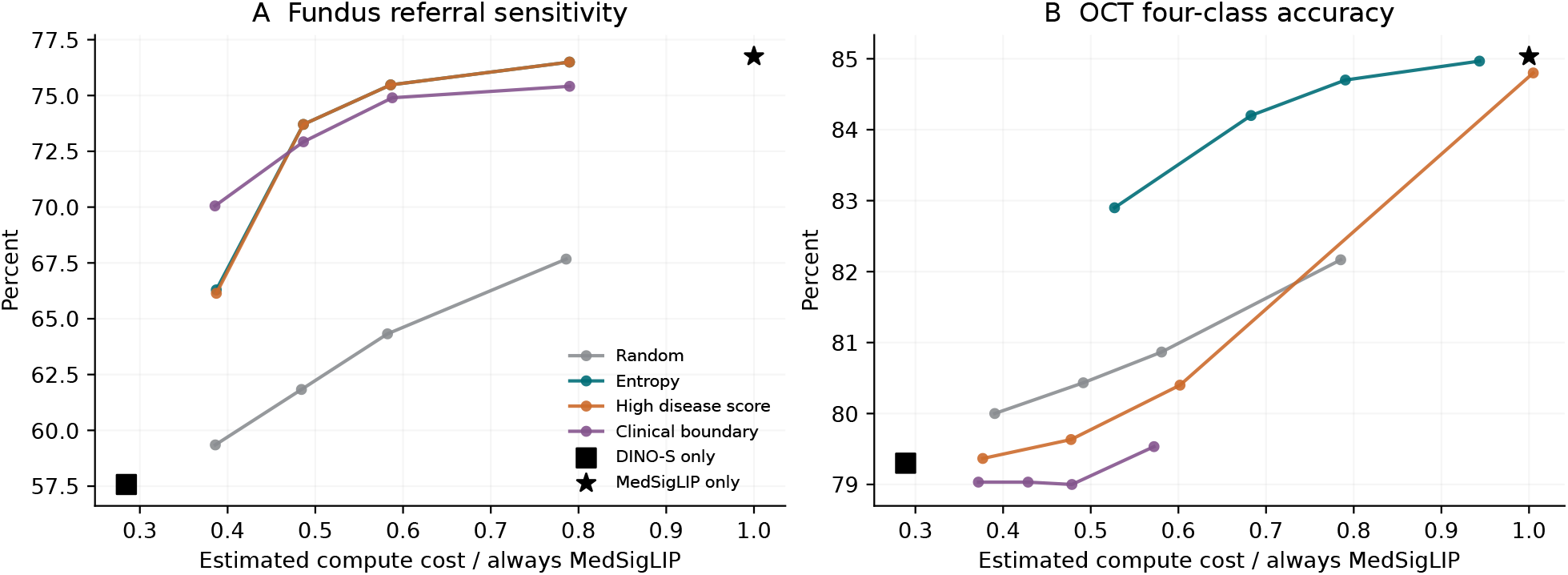
Real prediction cascades expose cost–performance trade-offs. Each curve includes all four calroute budgets, using three-seed mean test outcomes for text heads. Fundus referral thresholds are selected separately on calroute for each mixture; OCT shows four-class accuracy. Horizontal coordinates are component-cost estimates using measured single-image medians and actual test upgrade fractions, not direct cascade wall-clock measurements. The cheap model is always executed in a cascade. The two fundus entropy/disease curves substantially overlap.

### 9.8 Wording transfer changes the routing workload

For fundus entropy routing at the nominal 30% budget, changing only the cheap candidate descriptions increases upgrades from 30.13% to a mean 81.76% across the three held-out descriptions and three seeds. A temperature fitted for each new schema restores a mean 29.97%; affine recalibration yields 30.75%. These conditions reuse the canonical upgrade threshold and the strong model’s canonical predictions. Recalibration restores the behavior of an existing routing rule here; it does not merely choose a new test-set quantile.

At the same locked referral threshold, the uncorrected wording cascade changes sensitivity and specificity to 78.99% and 93.97%, versus the canonical 75.48% and 94.69%. After schema-specific temperature scaling they are 75.47% and 94.70%. The apparent increase in sensitivity before correction accompanies greater upgrade workload and more false-positive referrals. OCT shows additional transfer difficulty, including nominal–realized budget differences before the schema changes (Figure 13); fundus repair should not be generalized to all schemas or cohorts.

**Figure 13:**
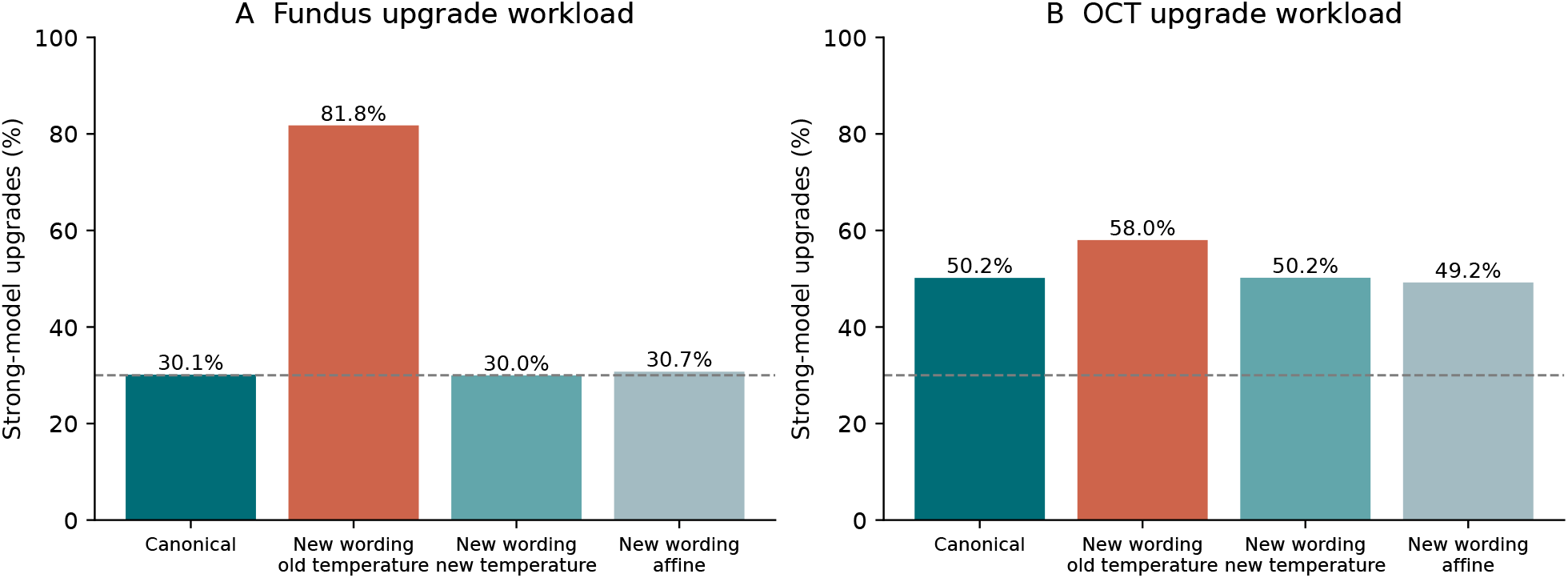
The calibration budget is not a deployment guarantee. Actual upgrades under a fixed entropy threshold selected for a 30% calroute budget. Canonical bars average three seeds; wording bars average the three prespecified held-out descriptions within each seed and then average seeds. Schema recalibration uses labeled calfit data. The strong model’s descriptions and both canonical decision thresholds remain fixed. OCT canonical workload already differs from the nominal budget; the source and test distributions differ in class composition and potentially other properties.

Single-image median forward times are 3.87 ms for DINOv2-S/text and 13.60 ms for Med-SigLIP/text on fundus images. The component-based mean-cost estimate is 7.97 ms for the canonical entropy cascade, 14.99 ms after uncorrected wording transfer, and 7.95 ms after schema temperature scaling. Thus estimated savings of 41.4% against the strong-only path become an estimated 10.2% overrun after a wording change. These estimates exclude image I/O and do not establish an API-cost advantage over an LLM. Candidate heads do not outperform linear heads in raw readout speed; their additional cost is small relative to image encoding.

On 512 actual fundus test images, live entropy routing takes a mean 11.33 ms, versus 18.61 ms for always executing MedSigLIP in the same randomly interleaved experiment: a 1.64-fold speed ratio and 39.1% lower mean forward latency. The 95th percentile does not improve (31.17 versus 30.33 ms). Live routing upgrades 168/512 cases, whereas replay of the saved batch-feature mask upgrades 167/512; seven branch decisions differ. The live path recomputes single-image BF16 representations, so these timings do not constitute an exact probability replay of the cached-feature clinical analysis. A separate mask-replay timing has mean 10.96 ms. Both encoders remain resident, peak reserved memory is 2.07 GiB, and no OOM occurs. All values exclude decoding, I/O and host-to-device transfer. This is a local forward benchmark under recorded background load, not a matched LLM or end-to-end service comparison.

### 9.9 External discrimination improves selectively and exposes negative transfer

Table 46 and Figure 14 summarize all six cohorts under the fixed main comparison. On Cháks.u, the text head reaches AUROC 0.873 versus 0.798 for the linear probe (difference 0.075, 95% CI 0.051–0.101), while the matched fixed MLP reaches 0.883. The text head’s entropy has descriptive AUROC 0.705 for nonunanimous five-reader votes; its reader Brier loss is 0.174 versus 0.181 for the linear probe, using the saved internal reader-target temperature. These five-reader results concern image-based suspect judgments and do not validate clinical diagnostic uncertainty.

Transfer is not uniformly favorable. The text-minus-linear AUROC interval includes zero on PAPILA (−0.014, 95% CI −0.038 to 0.015) and Drishti-GS (0.001, −0.020 to 0.029). RIM-ONE DL shows clear negative transfer: text AUROC is 0.693 versus 0.822 for the linear probe, a difference of −0.129 (−0.176 to −0.090). The fixed MLP is similarly affected (0.692). This shared failure argues against attributing the entire loss to candidate semantics, and prevents a general superiority claim for either nonlinear readout.

**Table 46:** Frozen external discrimination on six native cohorts. Values are mean AUROC across three fitted seeds using the same MedSigLIP visual backbone. Text uses MedSigLIP candidates; the fixed MLP matches its trainable capacity. Linear probes use internal G1/G2 soft targets for fundus and hard labels for OCT. PAPILA and OCTDL groups incorporate native patient identities and exact-pixel links; other rows have image groups without verified patient identifiers. Fundus endpoints are native glaucoma or suspect labels, OCTDL is conditional DME/normal discrimination, and CAVRI-H5 uses macro one-versus-rest AUROC over four classes. Values across rows are different estimands and must not be pooled or ranked as a common screening test.

| Cohort and endpoint | Images | Groups | Text | Fixed MLP | Linear |
| --- | --- | --- | --- | --- | --- |
| PAPILA | 420 | 210 | 0.814 | 0.815 | 0.827 |
| Chákṣu | 1338 | 1338 | 0.873 | 0.883 | 0.798 |
| RIM-ONE DL | 485 | 485 | 0.693 | 0.692 | 0.822 |
| Drishti-GS | 101 | 101 | 0.887 | 0.892 | 0.886 |
| OCTDL: DME/NO | 474 | 214 | 0.998 | 0.997 | 0.995 |
| CAVRI-H5: four classes | 474 | 474 | 0.973 | 0.973 | 0.972 |

On OCTDL’s restricted DME/normal subset, text AUROC is 0.9980 versus 0.9954 for the linear probe (difference 0.00252, 95% CI 0.00040–0.00595). Conditional accuracy improves from 95.78% to 97.26% (1.48 percentage points, 0.59–2.62), and summed binary Brier loss decreases by 0.02298 (0.01148–0.03754). The fixed MLP still has higher conditional accuracy (97.89%). Moreover, the text head’s unchanged four-class argmax is correct on only 85.44% of these same images and assigns mean probability 13.56% to CNV/drusen. Thus, the near-ceiling conditional AUROC does not establish four-class external accuracy or a stand-alone calibrated DME risk.

### 9.10 Four-class OCT transfer does not imply detection of an unsupported disease

CAVRI-H5 directly tests the complete CNV/DME/drusen/normal schema. With MedSigLIP visual features, the candidate text head achieves macro AUROC 0.9725, accuracy 80.17%, and Brier loss 0.3104. The linear probe achieves 0.9722, 78.76%, and 0.3295, respectively; all three text-minus-linear intervals include zero. The text accuracy difference is 1.41 percentage points (95% CI −0.60 to 3.45). The fixed MLP reaches macro AUROC 0.9732, accuracy 82.07%, and Brier loss 0.2787. Its accuracy improvement over the linear probe is 3.31 percentage points (1.48–5.20), while the text head is worse than the fixed MLP by 1.90 points (−3.52 to −0.10) and has higher Brier loss by 0.0317 (0.0150–0.0488). These findings support useful frozen four-class transfer, but not a text-specific advantage.

Class-level analysis exposes a gap hidden by high macro AUROC: text-head argmax sensitivity for drusen is 47.45%, compared with 56.45% for the fixed MLP (Figure 15). Nor does the candidate interface reliably flag the unsupported VMA–VMT category. Its entropy-based VMA–VMT-versus-known AUROC is 0.473; the internal entropy threshold rejects only 8.64% of the 135 unsupported images while rejecting 14.21% of known-class images. This is a failed uncertainty stress test, not evidence of new-disease recognition. The 473-component basename sensitivity analysis preserves the principal paired conclusions above.

### 9.11 External threshold and workload transfer remain separate problems

The frozen referral threshold transports poorly even where discrimination remains useful. The text head’s sensitivity is 40.23% on PAPILA, 19.43% on Cháks.u, and 3.29% on RIM-ONE DL, with specificities of 96.20%, 98.67%, and 99.57%. These are different native endpoints and case mixes, not estimates of one screening population. False-positive and false-negative counts per 1,000 in the release use each cohort’s observed prevalence.

**Figure 14:**
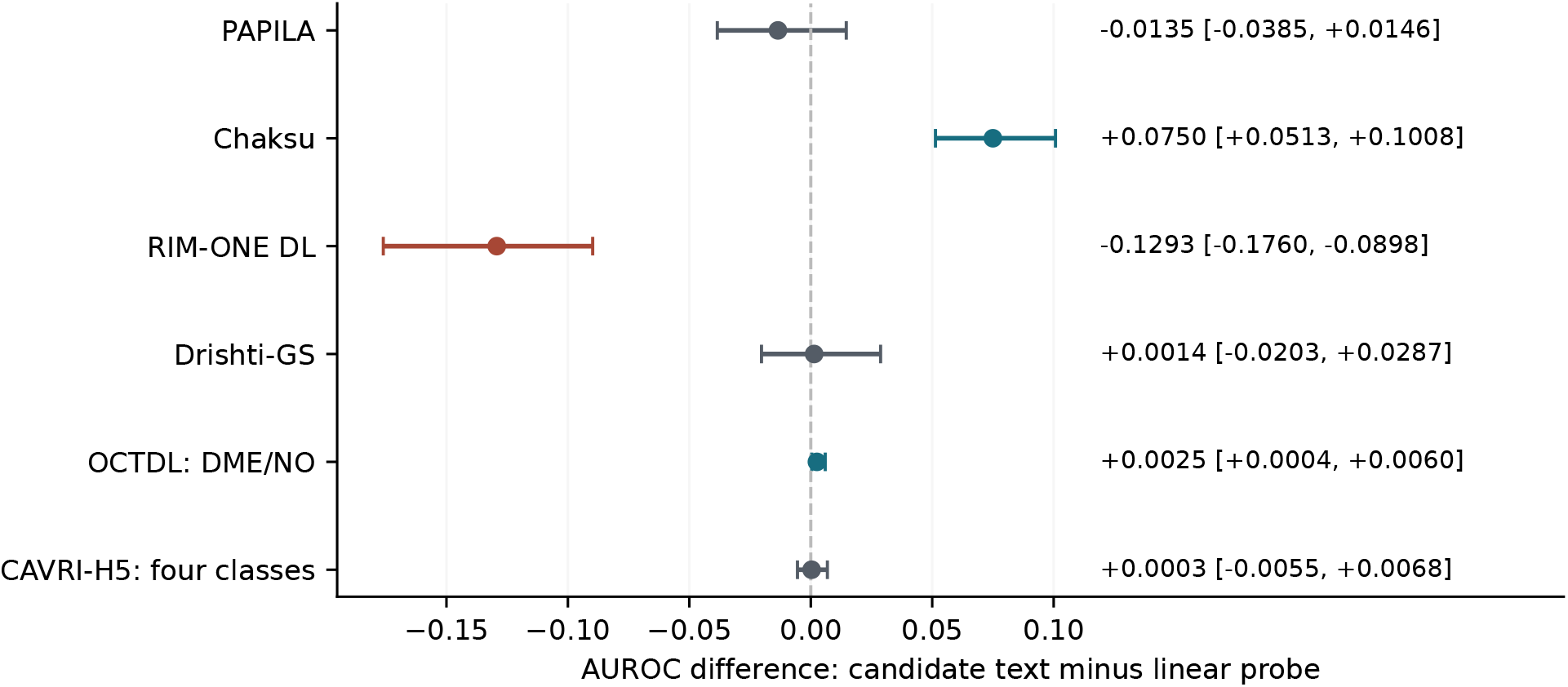
External gains depend on the cohort and endpoint. Points compare the Med-SigLIP candidate-text head with the conventional linear probe on the same visual representation. Bars are 500-draw paired 95% percentile intervals for the mean metric across three fixed seed fits. Patient-plus-pixel components are sampled for PAPILA and OCTDL; other intervals use image components. CAVRI-H5 uses four-class macro AUROC, OCTDL conditional DME/normal AUROC, and fundus datasets their native binary labels. There is no pooled effect. Intervals are exploratory and not multiplicity-adjusted.

The nominal 30% internal text-cascade budget yields canonical external upgrade rates of 31.19% on PAPILA, 57.00% on Cháks.u, 16.01% on RIM-ONE DL, 61.39% on Drishti-GS, 50.63% on OCTDL, and 47.12% on CAVRI-H5. On PAPILA, two held-out descriptions raise upgrades to 100%, whereas their saved internal schema temperatures restore approximately 31.3%. On CAVRI-H5, held-out wording raises mean upgrade use to 57.69%, versus 48.03% with those internal schema temperatures. Recalibration therefore reduces the additional wording effect without restoring the nominal internal budget under cohort shift.

The canonical CAVRI text cascade achieves 80.52% accuracy while upgrading 47.12% of images, compared with 80.17% when always using the stronger model. This descriptive difference has no noninferiority margin or external latency measurement. The RIM-ONE stronger model is itself less discriminative than the cheap model (AUROC 0.693 versus 0.792); upgrading cannot be assumed beneficial after transfer. Together, these results distinguish three requirements: transport of disease discrimination, transport of decision thresholds, and transport of computational workload.

### 9.12 Fusion improves selected rankings but does not dominate all cohorts

Source-only selection favors the anchored mixture among the five prespecified families. Relative to its newly calibrated single-expert anchor, this mixture improves mean AUROC on seven of eight internal-plus-external endpoints, while reducing Cháks.u AUROC from 0.885 to 0.849. This is a descriptive direction count, not a claim of eight independent confirmatory tests. Accuracy and proper scores do not improve uniformly. The six original single-expert results and all mixture families remain available.

The subsequent clinical adapter treats the previously inspected cohorts as development data and reports LOCO results separately. Relative to the source-only mixture, joint adaptation improves mean AUROC and accuracy on five of the six old cohorts. Cháks.u instead decreases from 0.849 to 0.832 AUROC and from 86.37% to 84.01% accuracy. OCTDL conditional Brier also worsens slightly, from 0.0271 to 0.0282. These outcomes remain exploratory because prior cohort results motivated the extension. Bias-only adaptation explains much of the fundus accuracy change while preserving binary ranking, motivating the new-cohort comparisons below.

**Figure 15:**
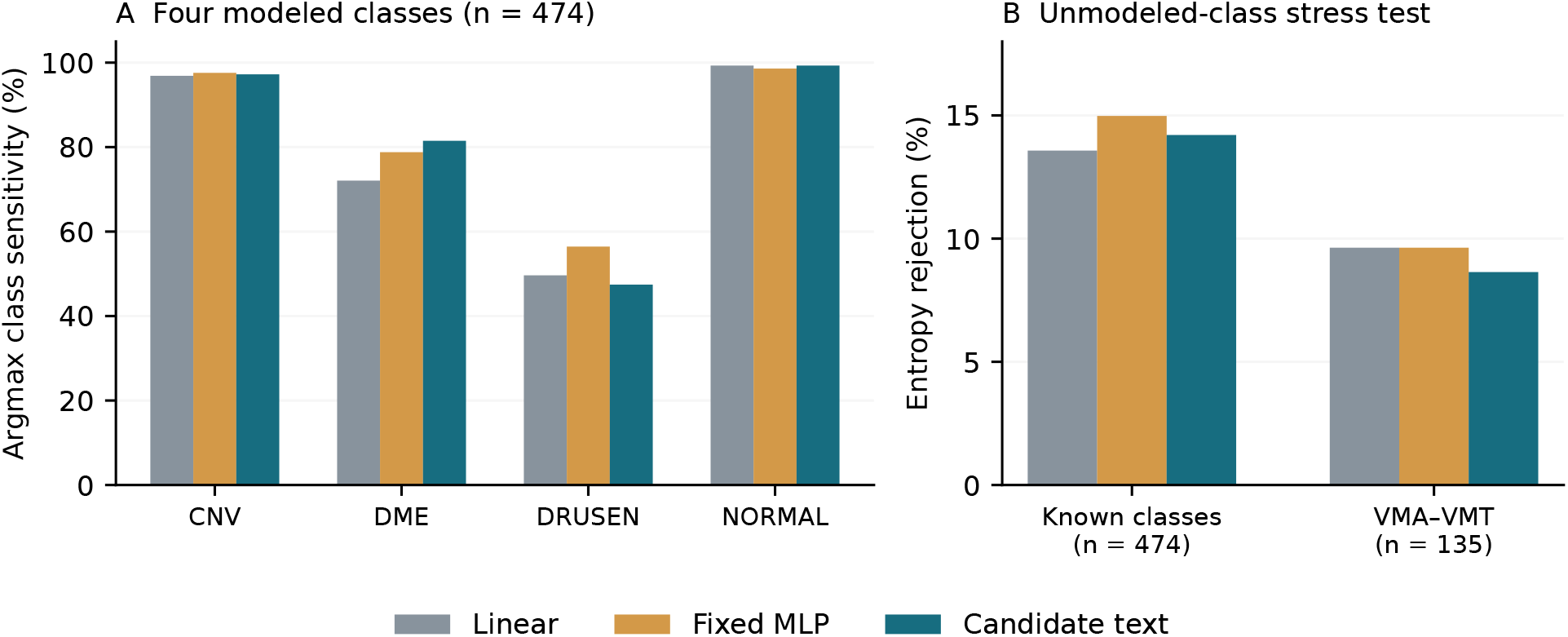
CAVRI-H5 separates four-class discrimination from two clinically relevant failure modes. (A) Argmax sensitivity by native class, averaged over three fitted seeds, reveals low drusen sensitivity despite macro AUROC near 0.97. (B) The original internal 95th-percentile entropy threshold rejects a smaller fraction of unsupported VMA–VMT inputs than known-class inputs. All panels use the same MedSigLIP visual representation and canonical internal calibrators. Bars show descriptive seed means, not independent patient-level estimates or confidence intervals. No external labels set the thresholds, and no fifth-class model is fitted.

### 9.13 New clinical glaucoma data reveal partial correction and persistent failure

Table 47 reports all original single experts alongside the frozen fusion and calibration ablations. On HYGD, the primary adapter reaches AUROC 0.7128, accuracy 50.25%, balanced accuracy 63.90%, and summed binary Brier 0.7377. Against original MedSigLIP text, the accuracy difference is 18.05 percentage points (95% paired CI 14.08–21.99) and Brier decreases by 0.4938 (0.4387–0.5503). However, the AUROC difference of 0.0177 has interval −0.0096 to 0.0440. Against the matched MedSigLIP reader-target linear probe, the AUROC difference is 0.0286 (0.0033–0.0534). These comparisons use additional supervised development cohorts for the adapter, whereas the original controls retain source-only training and calibration.

The improvement remains inadequate in absolute terms. DINOv2-B’s original hard-label linear probe has higher AUROC, 0.7893, than the adapter. Predicting the cohort’s majority class would obtain 73.27% accuracy, and an uninformative probability of one-half has Brier 0.5; the adapter falls short of both references, although its balanced accuracy exceeds chance. Its frozen operating threshold yields only 25.31% sensitivity at 96.62% specificity. Thus the reduction of a large calibration error is not evidence of an adequate glaucoma screening system.

Bias-only adaptation moves source-mixture accuracy from 30.85% to 49.21% without changing AUROC (0.7061); joint fusion then reaches 50.25% accuracy and 0.7128 AUROC. The joint-minus-source AUROC interval includes zero (−0.0020 to 0.0156). Most of the observed accuracy improvement is therefore reproduced by the intercept-only arm. This controlled contrast separates changes in probability scale and default decisions from stronger disease ranking, without identifying prevalence as their sole cause.

**Table 47:** Locked evaluation preserves all single-expert controls and calibration ablations. Values are means of three seed metrics. HYGD uses unique-image weights with 283 patient/pixel resampling components; Srinivasan averages four-class probabilities within each subject before conditional DME/normal evaluation. Reader-target linear training is available only for fundus. All original heads retain their original source calibration. The adapter additionally uses previously inspected development cohorts, so comparisons with original controls do not match supervised adaptation resources. The uniform reference assigns probability one-half to both endpoint classes; its argmax tie is assigned to the negative class, consistently with model evaluation. The HYGD majority-class accuracy is 73.27%, exceeding every model in this table; fusion balanced accuracy is 63.90%.

| Method | HYGD: 737 images |  |  | Srinivasan: 30 subjects |  |  |
| --- | --- | --- | --- | --- | --- | --- |
|  | AUC | Acc. (%) | Brier | AUC | Acc. (%) | Brier |
| MedSigLIP linear (hard) | 0.678 | 31.43 | 1.231 | 1.000 | 87.78 | 0.135 |
| MedSigLIP linear (reader) | 0.684 | 32.43 | 1.214 | – | – | – |
| MedSigLIP fixed | 0.689 | 31.39 | 1.235 | 1.000 | 93.33 | 0.105 |
| MedSigLIP text | 0.695 | 32.20 | 1.232 | 1.000 | 94.44 | 0.095 |
| DINOv2-B linear (hard) | 0.789 | 27.45 | 1.323 | 1.000 | 95.56 | 0.100 |
| DINOv2-B fixed | 0.702 | 28.40 | 1.287 | 1.000 | 97.78 | 0.087 |
| DINOv2-B text | 0.659 | 29.40 | 1.301 | 1.000 | 98.89 | 0.078 |
| Source mixture | 0.706 | 30.85 | 1.255 | 1.000 | 94.44 | 0.093 |
| Bias-only adapter | 0.706 | 49.21 | 0.775 | 1.000 | 97.78 | 0.076 |
| Fusion + intercepts | 0.713 | 50.25 | 0.738 | 1.000 | 98.89 | 0.073 |
| Uniform binary reference | 0.500 | 26.73 | 0.500 | 0.500 | 50.00 | 0.500 |

### 9.14 Small-cohort OCT ranking saturates while calibration remains distinguishable

All compared methods attain AUROC 1.0 on the 30-subject Srinivasan DME/normal endpoint. This saturation cannot distinguish a superior ranker. The primary adapter’s accuracy is 98.89%, matching the DINOv2-B text head, versus 94.44% for MedSigLIP text and 93.33% for its fixed MLP. The adapter-minus-text accuracy interval includes zero (0.00–10.00 percentage points), whereas summed Brier improves from 0.0950 to 0.0733 (difference −0.0217, 95% CI −0.0441 to −0.0032). Relative to the MedSigLIP hard-label linear probe, accuracy improves by 11.11 points (3.33–20.00). Figure 16 shows the corresponding proper-score contrasts.

These are averages of metrics over three models on the same 30 subjects, not 90 independent participants. The zero observed AUROC differences and degenerate bootstrap intervals arise because every resample contains only perfectly ranked observed cases; they do not establish certain population-level equivalence. Full four-class decisions, probability outside the DME/normal pair, and both slice-weighting analyses accompany the primary table in the aggregate release. On the exploratory 45-subject any-pathology endpoint, adapter accuracy is 99.26%, while DINOv2-B text reaches 100%. Neither conditional discrimination nor this broader abnormality endpoint validates three-class dry-AMD diagnosis.

### 9.15 Matching adaptation resources limits the case for fusion superiority

Table 48 adds post-hoc single-expert controls receiving the same development supervision and intercept-fitting budget. On HYGD, the DINOv2-B linear control retains AUROC 0.7893 and reaches Brier 0.6659. The primary fusion is worse on both (AUROC difference −0.0766, 95% CI −0.1282 to −0.0362; Brier difference 0.0719, 0.0233–0.1258). Accuracy differs by −1.13 points with an interval spanning zero. Even the adapted MedSigLIP hard-label linear probe has lower Brier (0.6973): fusion is worse by 0.0405 (0.0187–0.0648). The large improvement over the original unadapted heads therefore does not establish that combining six experts is necessary.

On Srinivasan, adapted DINOv2-B text matches fusion accuracy at 98.89% and has Brier 0.0660 versus 0.0733; the Brier difference interval spans zero (−0.0057 to 0.0236). All adapted single experts retain AUROC 1.0. These controls leave no consistent new-cohort superiority claim for the primary fusion.

**Figure 16:**
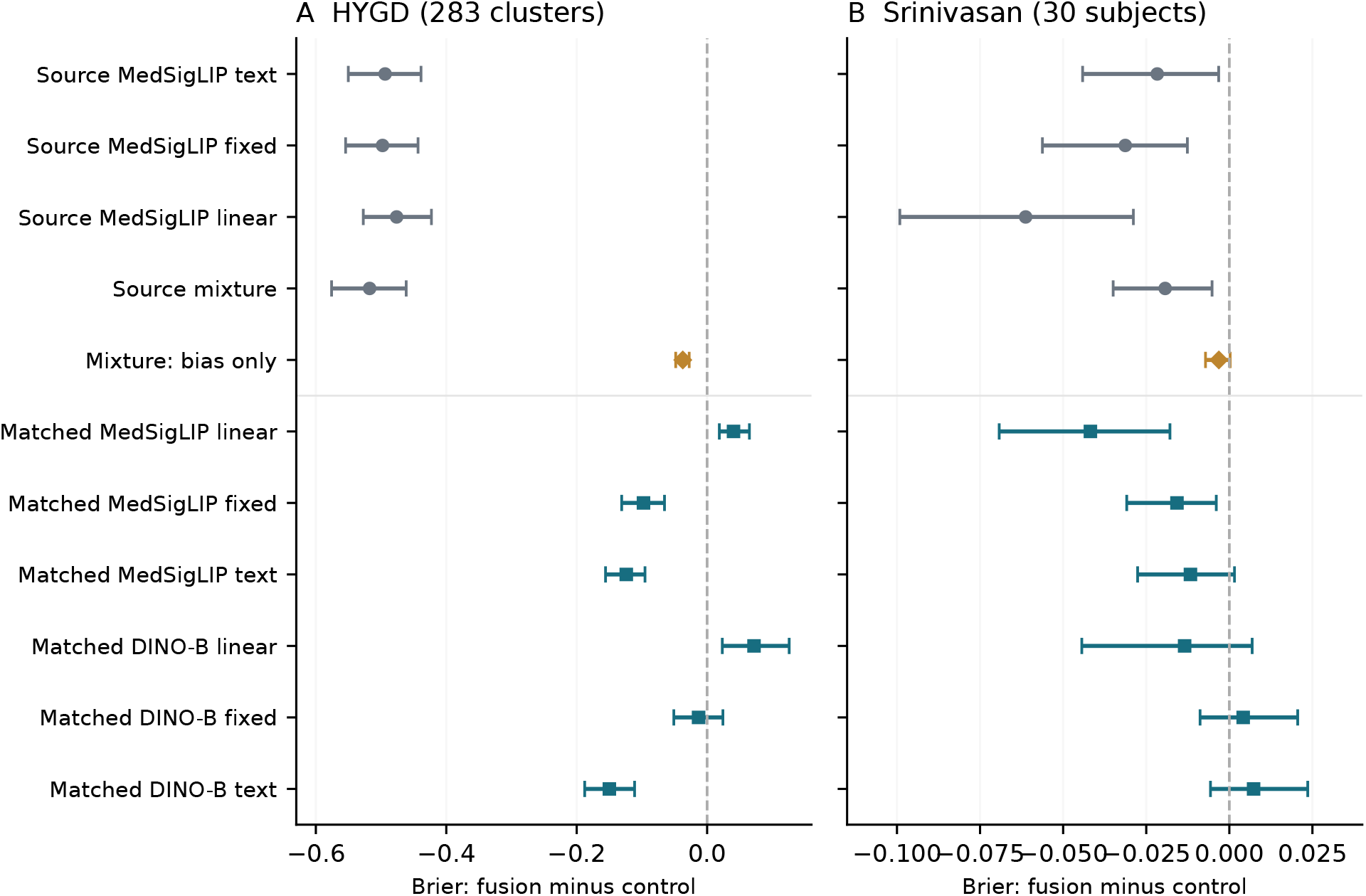
Favorable and unfavorable proper-score contrasts remain visible under matched supervision. Points are primary fusion minus control Brier differences, averaged over three seed metrics. Gray circles use original source-only controls; orange diamonds use mixture bias-only adaptation; teal squares use post-hoc single experts with matched development supervision. Source MedSigLIP linear means reader-target training for HYGD and hard-label training for OCT; the matched linear controls use hard-label experts. Bars are 500-draw paired diagnosis-stratified cluster percentile intervals. Panel scales differ. Negative values favor fusion, but matched DINOv2-B and MedSigLIP linear controls have lower HYGD Brier. Fusion’s absolute HYGD Brier is 0.738, worse than the uninformative reference of 0.5. Intervals condition on fitted models and have no multiplicity correction.

The complete old-cohort LOCO matrix (Supplementary Table 53) contains both favorable and unfavorable contrasts under matched supervision. On CAVRI-H5, fusion exceeds the adapted Med-SigLIP fixed head in macro AUROC by 0.0061 (0.0016–0.0110) and lowers Brier by 0.0187 (0.0005–0.0365); the accuracy interval includes zero. Yet fusion versus the source-only ensemble has intervals spanning zero for all three metrics, so these gains cannot be assigned specifically to external-cohort adaptation. On Cháks.u, fusion is worse than the adapted MedSigLIP fixed head in AUROC, accuracy, and Brier. Drishti Brier is also worse than bias-only adaptation by 0.0189 (0.0050–0.0327). The CAVRI shared-basename sensitivity does not change its interval-direction conclusions. All comparisons are exploratory and unadjusted for multiplicity.

### 9.16 Adaptation changes the original source decision contract

The fit-all clinical adapter is also evaluated on the original internal tests after freezing, without using the result for selection. In fundus screening, argmax accuracy decreases from 97.53% for the source mixture to 90.31%, a 7.23-point regression, and Brier increases from 0.0379 to 0.1490. AUROC remains approximately 0.9623. The bias-only arm shows the same trade-off. At the separately re-established source operating threshold, sensitivity and specificity change little: 79.99%/94.99% for the source mixture and 79.74%/95.05% after joint adaptation. Thus the regression concerns the default decision and probability contract, rather than a corresponding seven-point loss of sensitivity at the source-specific operating point.

**Table 48:** Matching development supervision narrows the interpretation of fusion gains. Every single expert receives bounded class-intercept fitting on the same allowed development cohorts with the same group weights, endpoint losses, regularization, and frozen source-temperature convention. These controls were added after inspecting primary new-cohort outcomes; they are post-hoc component ablations, not a replacement for the locked primary analysis. No HYGD or Srinivasan labels fit the controls. All Srinivasan AUROCs are 1.0. Values are three-seed means with the same units as Table 47. The shared temperature convention does not exhaust all possible single-expert calibration choices.

| Adapter evidence | HYGD: unique images |  |  | Srinivasan: subjects |  |
| --- | --- | --- | --- | --- | --- |
|  | AUC | Acc. (%) | Brier | Acc. (%) | Brier |
| MedSigLIP linear (hard) | 0.678 | 51.20 | 0.697 | 92.22 | 0.115 |
| MedSigLIP fixed | 0.689 | 49.30 | 0.835 | 96.67 | 0.089 |
| MedSigLIP text | 0.695 | 47.99 | 0.862 | 94.44 | 0.085 |
| DINOv2-B linear (hard) | 0.789 | 51.38 | 0.666 | 96.67 | 0.087 |
| DINOv2-B fixed | 0.702 | 48.21 | 0.751 | 98.89 | 0.069 |
| DINOv2-B text | 0.659 | 42.70 | 0.888 | 98.89 | 0.066 |
| Six-expert fusion + intercepts | 0.713 | 50.25 | 0.738 | 98.89 | 0.073 |

For four-class internal OCT, the same adaptation procedure improves accuracy from 85.20% to 91.03% and Brier from 0.2239 to 0.1330 (Figure 17). The two modalities therefore provide different outcomes of the same development rule. An adapter fitted for diagnosis-oriented cohorts cannot be substituted silently for the original screening calibrator. The experiment has no automatic clinical-endpoint selector and no evidence that one state is uniformly appropriate across settings.

## 10 Supplementary methods and complete descriptions

### 10.1 Training and numerical protocol

All candidate heads use 128-dimensional, two-layer GELU image/text projectors with LayerNorm, followed by a shared 384 → 128 → 1 scorer. Fixed heads use the same image projector and choose their hidden width to approximate the candidate-head parameter budget. AdamW uses learning rate 10*^−^*^3^, weight decay 10*^−^*^4^, batch size 1,024, and gradient-norm clipping at 1.0. The epoch cap is 30; patience is five epochs with a minimum development-NLL improvement of 10*^−^*^5^. The consistency coefficient is fixed at 0.1. Both text arms average supervision across two cyclic training templates per batch. Development selection averages hard-label NLL across all three training descriptions. Image feature standardization uses only eligible fitting rows; text vectors are L2-normalized and multiplied by the square root of their dimension.

A numerical preflight using last-layer mean pooling for the FLAIR-adapted BERT produced almost constant candidate vectors and failed checkpoint replay. This was an incompatible feature-extraction choice: the pretrained FLAIR readout uses multiple hidden layers and its learned projection. All outcomes from that attempt are excluded. The native FLAIR readout was restored and verified before the complete revised matrix. Projector LayerNorm and gradient clipping were applied uniformly to every final arm. The final analysis is consequently development-informed and is not described as a blind preregistered experiment. These changes address input compatibility and numerical reproducibility; they were not selections of the highest test score.

MedSigLIP text inputs use fixed padding to 64 tokens and its pretrained tokenizer. The FLAIR-adapted BERT is restored from the complete local checkpoint with strict key matching; its native forward averages hidden layers 1, 2, and the last layer over tokens, normalizes, applies the learned text projection, and normalizes again. It supplies a 512-dimensional representation from a 768-dimensional BERT backbone, compared with 1,152 output dimensions for MedSigLIP text. Per-example versus batched extraction is checked to distinguish padding artifacts from description effects.

**Figure 17:**
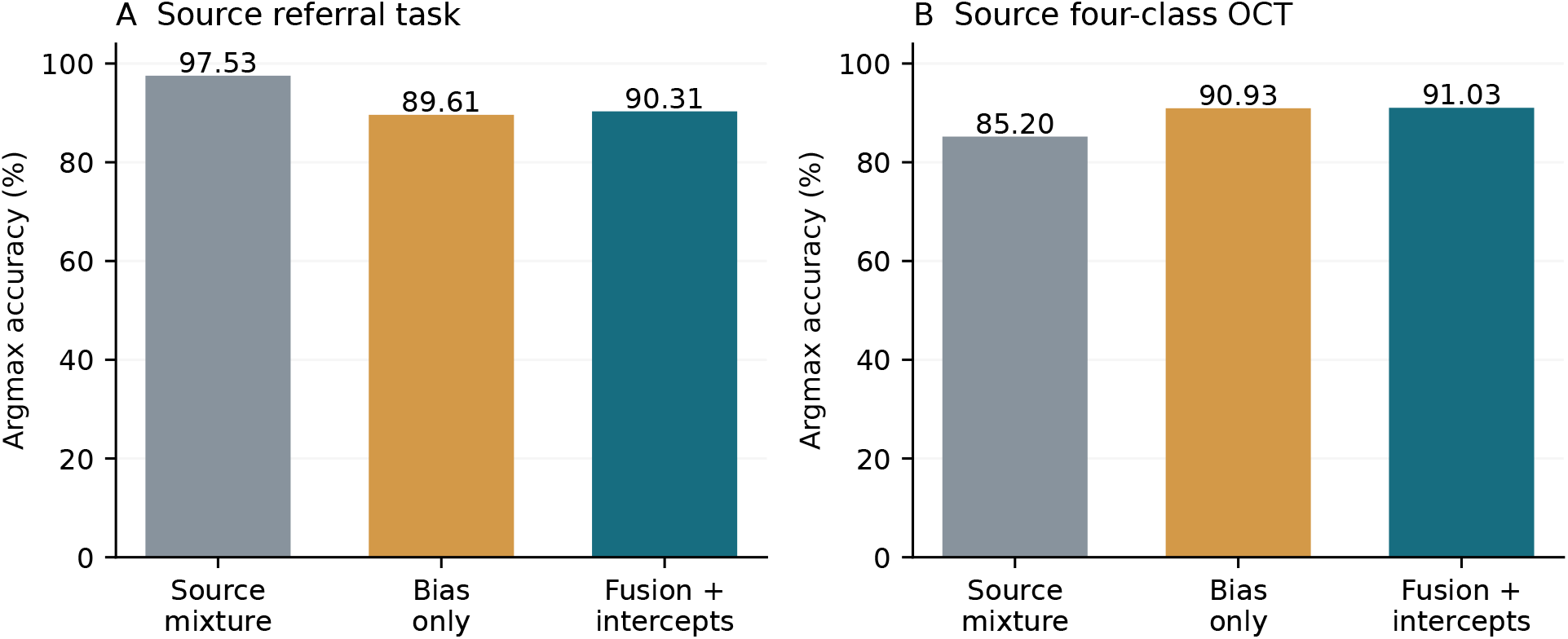
Retaining the source-task audit exposes a clinically consequential trade-off. Bars show mean argmax accuracy over three seed fits on each original internal test. The fundus clinical adapter worsens screening accuracy and Brier while leaving AUROC almost unchanged; the OCT adapter improves four-class accuracy. Bias-only adaptation reproduces much of both changes. These are descriptive fixed-model seed means, with neither a pooled cross-task effect nor a claim of independent confidence intervals. Separate source operating-threshold results are reported in the text.

Schema-specific affine calibration uses a positive scalar logit slope and class biases constrained to sum to zero. It is fitted only on the calibration partition and is reported separately from scalar temperature. Biases can change the operating point and, in multiclass tasks, the predicted class; affine calibration is therefore a stronger intervention than temperature.

### 10.2 Partition sizes

| Task | Seed | Fit | Dev | Calibration | Threshold | Test |
| --- | --- | --- | --- | --- | --- | --- |
| fundus | 2027 | 63,822 | 7,091 | 5,068 | 5,067 | 20,375 |
| fundus | 2028 | 63,895 | 7,100 | 5,076 | 5,078 | 20,274 |
| fundus | 2029 | 63,824 | 7,091 | 5,070 | 5,070 | 20,368 |
| oct | 2027 | 86,347 | 9,595 | 5,415 | 5,417 | 1,000 |
| oct | 2028 | 86,347 | 9,595 | 5,416 | 5,416 | 1,000 |
| oct | 2029 | 86,348 | 9,594 | 5,415 | 5,417 | 1,000 |

Fundus fitting is restricted to cases with two binary reader responses. Development and clinical calibration retain released final labels for all cases in their partitions; reader metrics and reader calibration use the eligible mask. OCT uses every remaining row in each partition. Exact sample indices, eligibility masks, and checksums are retained locally with each run.

### 10.3 Fundus descriptions

Question: Is this fundus photograph referable for glaucomatous optic neuropathy?

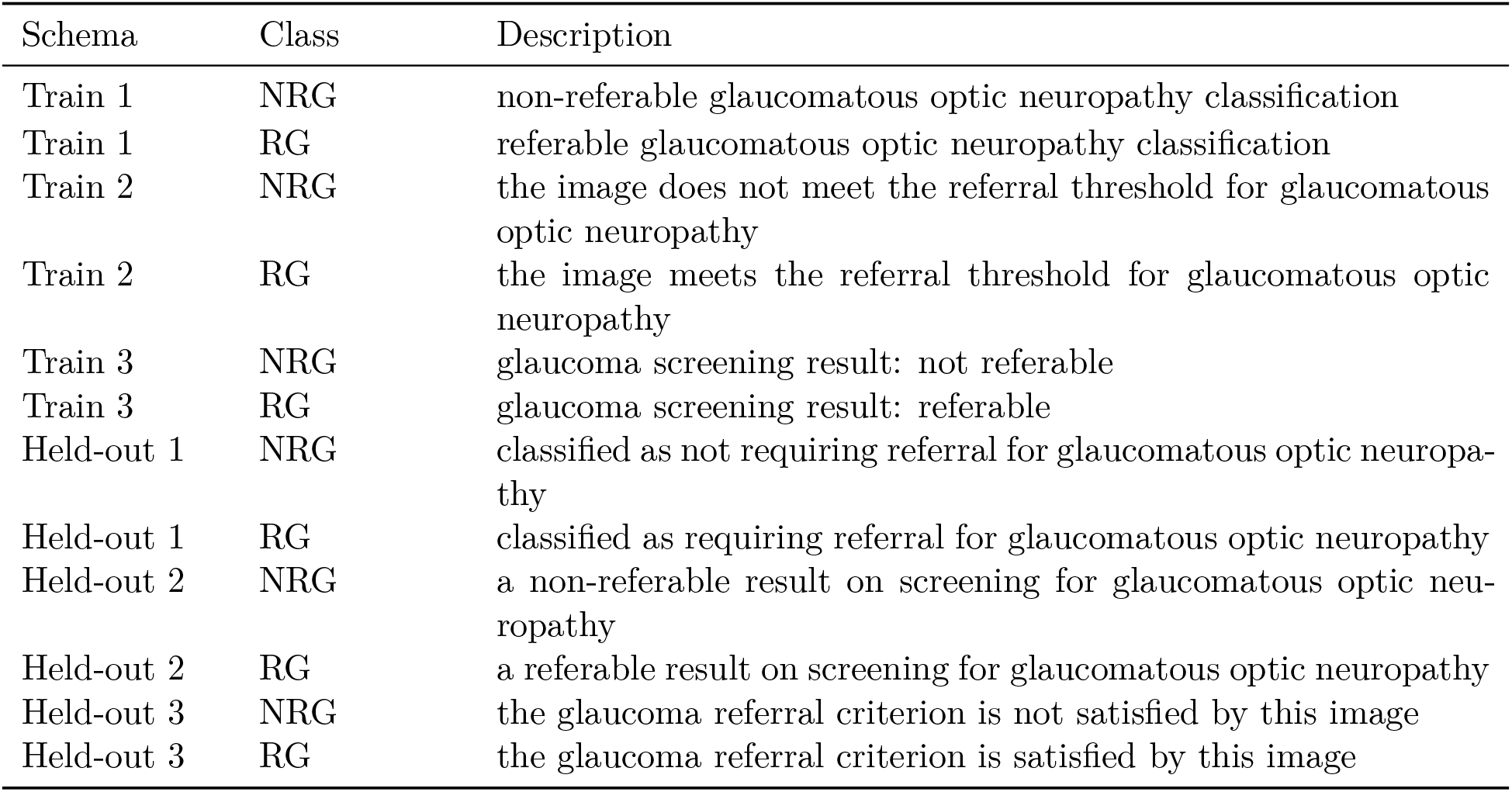

| Schema | Class | Description |
| --- | --- | --- |
| Train 1 | NRG | non-referable glaucomatous optic neuropathy classification |
| Train 1 | RG | referable glaucomatous optic neuropathy classification |
| Train 2 | NRG | the image does not meet the referral threshold for glaucomatous optic neuropathy |
| Train 2 | RG | the image meets the referral threshold for glaucomatous optic neuropathy |
| Train 3 | NRG | glaucoma screening result: not referable |
| Train 3 | RG | glaucoma screening result: referable |
| Held-out 1 | NRG | classified as not requiring referral for glaucomatous optic neuropathy |
| Held-out 1 | RG | classified as requiring referral for glaucomatous optic neuropathy |
| Held-out 2 | NRG | a non-referable result on screening for glaucomatous optic neuropathy |
| Held-out 2 | RG | a referable result on screening for glaucomatous optic neuropathy |
| Held-out 3 | NRG | the glaucoma referral criterion is not satisfied by this image |
| Held-out 3 | RG | the glaucoma referral criterion is satisfied by this image |

### 10.4 OCT descriptions

Question: Which of the four dataset categories describes this retinal OCT image?

10.4 OCT descriptions Question: Which of the four dataset categories describes this retinal OCT image?
| Schema | Class | Description |
| --- | --- | --- |
| Train 1 | CNV | choroidal neovascularization |
| Train 1 | DME | diabetic macular edema |
| Train 1 | DRUSEN | drusen |
| Train 1 | NORMAL | the dataset normal OCT category |
| Train 2 | CNV | CNV: choroidal neovascularization |
| Train 2 | DME | DME: diabetic macular edema |
| Train 2 | DRUSEN | drusen deposits |
| Train 2 | NORMAL | normal OCT findings within the dataset classification |
| Train 3 | CNV | the choroidal neovascularization category |
| Train 3 | DME | the diabetic macular edema category |
| Train 3 | DRUSEN | the drusen category |
| Train 3 | NORMAL | the normal OCT category |
| Held-out 1 | CNV | OCT classified as choroidal neovascularization |
| Held-out 1 | DME | OCT classified as diabetic macular edema |
| Held-out 1 | DRUSEN | OCT classified as drusen |
| Held-out 1 | NORMAL | OCT classified as normal in this dataset |
| Held-out 2 | CNV | a choroidal neovascularization finding |
| Held-out 2 | DME | a diabetic macular edema finding |
| Held-out 2 | DRUSEN | a drusen finding |
| Held-out 2 | NORMAL | a normal OCT finding under the dataset labels |
| Held-out 3 | CNV | the image belongs to the CNV class (choroidal neovascularization) |
| Held-out 3 | DME | the image belongs to the DME class (diabetic macular edema) |
| Held-out 3 | DRUSEN | the image belongs to the drusen class |
| Held-out 3 | NORMAL | the image belongs to the dataset-defined normal class |

## 11 Supplementary numerical results

**Table 51:** All settings, mean over three seeds. M denotes Med-SigLIP text and B denotes FLAIR-BERT. Held-out values average the three descriptions within seed. *B_C_*: canonical Brier; *B_H_*: transferred-calibrator held-out Brier; *B_T_*: new-schema temperature; *B_A_*: new-schema affine. Full per-seed NLL, ECE, AP, and intervention values are in the accompanying CSV/JSON.

| Visual / tower | Head | Params | AUC<sub>C</sub> | AUC<sub>H</sub> | $B_C$ | $B_H$ | $B_T$ | $B_A$ |
| --- | --- | --- | --- | --- | --- | --- | --- | --- |
| **Fundus** |  |  |  |  |  |  |  |  |
| DINOv2-B/B | Fixed | 246,988 | 0.921 | 0.921 | 0.047 | 0.047 | 0.047 | 0.046 |
| DINOv2-B/B | Random ID | 247,041 | 0.922 | 0.922 | 0.047 | 0.047 | 0.047 | 0.046 |
| DINOv2-B/B | Text | 247,041 | 0.918 | 0.640 | 0.047 | 0.299 | 0.198 | 0.052 |
| DINOv2-B/B | Text + JS | 247,041 | 0.923 | 0.641 | 0.047 | 0.284 | 0.198 | 0.052 |
| DINOv2-B/M | Fixed | 328,994 | 0.918 | 0.918 | 0.047 | 0.047 | 0.047 | 0.046 |
| DINOv2-B/M | Random ID | 328,961 | 0.921 | 0.921 | 0.047 | 0.047 | 0.047 | 0.046 |
| DINOv2-B/M | Text | 328,961 | 0.921 | 0.921 | 0.048 | 0.092 | 0.048 | 0.046 |
| DINOv2-B/M | Text + JS | 328,961 | 0.922 | 0.922 | 0.047 | 0.085 | 0.047 | 0.046 |
| DINOv2-S/M | Fixed | 279,842 | 0.906 | 0.906 | 0.051 | 0.051 | 0.051 | 0.050 |
| DINOv2-S/M | Random ID | 279,809 | 0.906 | 0.906 | 0.051 | 0.051 | 0.051 | 0.050 |
| DINOv2-S/M | Text | 279,809 | 0.907 | 0.907 | 0.051 | 0.102 | 0.050 | 0.050 |
| DINOv2-S/M | Text + JS | 279,809 | 0.908 | 0.908 | 0.050 | 0.101 | 0.050 | 0.050 |
| MedSigLIP/B | Fixed | 296,140 | 0.949 | 0.949 | 0.041 | 0.041 | 0.041 | 0.040 |
| MedSigLIP/B | Random ID | 296,193 | 0.951 | 0.951 | 0.040 | 0.040 | 0.040 | 0.040 |
| MedSigLIP/B | Text | 296,193 | 0.952 | 0.652 | 0.040 | 0.274 | 0.194 | 0.048 |
| MedSigLIP/B | Text + JS | 296,193 | 0.952 | 0.651 | 0.040 | 0.288 | 0.194 | 0.048 |
| MedSigLIP/M | Fixed | 378,146 | 0.950 | 0.950 | 0.040 | 0.040 | 0.040 | 0.040 |
| MedSigLIP/M | Random ID | 378,113 | 0.952 | 0.952 | 0.040 | 0.040 | 0.040 | 0.040 |
| MedSigLIP/M | Text | 378,113 | 0.953 | 0.953 | 0.041 | 0.075 | 0.040 | 0.040 |
| MedSigLIP/M | Text + JS | 378,113 | 0.953 | 0.954 | 0.040 | 0.088 | 0.040 | 0.040 |
| RETFound-Green/M | Fixed | 279,842 | 0.917 | 0.917 | 0.049 | 0.049 | 0.049 | 0.048 |
| RETFound-Green/M | Random ID | 279,809 | 0.916 | 0.916 | 0.049 | 0.049 | 0.049 | 0.048 |
| RETFound-Green/M | Text | 279,809 | 0.923 | 0.922 | 0.048 | 0.097 | 0.047 | 0.047 |
| RETFound-Green/M | Text + JS | 279,809 | 0.919 | 0.919 | 0.048 | 0.092 | 0.048 | 0.047 |
| RETFound-CFP/M | Fixed | 361,762 | 0.878 | 0.878 | 0.051 | 0.051 | 0.051 | 0.051 |
| RETFound-CFP/M | Random ID | 361,729 | 0.881 | 0.881 | 0.050 | 0.050 | 0.050 | 0.050 |
| RETFound-CFP/M | Text | 361,729 | 0.880 | 0.880 | 0.051 | 0.107 | 0.051 | 0.050 |
| RETFound-CFP/M | Text + JS | 361,729 | 0.879 | 0.878 | 0.050 | 0.106 | 0.050 | 0.050 |
| **OCT** |  |  |  |  |  |  |  |  |
| DINOv2-B/B | Fixed | 247,007 | 0.989 | 0.989 | 0.265 | 0.265 | 0.265 | 0.270 |
| DINOv2-B/B | Random ID | 247,041 | 0.989 | 0.989 | 0.272 | 0.272 | 0.272 | 0.267 |
| DINOv2-B/B | Text | 247,041 | 0.991 | 0.987 | 0.253 | 0.256 | 0.260 | 0.249 |
| DINOv2-B/B | Text + JS | 247,041 | 0.991 | 0.987 | 0.253 | 0.256 | 0.260 | 0.249 |
| DINOv2-B/M | Fixed | 328,935 | 0.991 | 0.991 | 0.246 | 0.246 | 0.246 | 0.254 |
| DINOv2-B/M | Random ID | 328,961 | 0.988 | 0.988 | 0.269 | 0.269 | 0.269 | 0.271 |
| DINOv2-B/M | Text | 328,961 | 0.987 | 0.984 | 0.273 | 0.287 | 0.294 | 0.273 |
| DINOv2-B/M | Text + JS | 328,961 | 0.987 | 0.984 | 0.273 | 0.288 | 0.295 | 0.273 |
| DINOv2-S/M | Fixed | 279,783 | 0.990 | 0.990 | 0.290 | 0.290 | 0.290 | 0.268 |
| DINOv2-S/M | Random ID | 279,809 | 0.990 | 0.990 | 0.283 | 0.283 | 0.283 | 0.275 |
| DINOv2-S/M | Text | 279,809 | 0.988 | 0.983 | 0.308 | 0.338 | 0.347 | 0.297 |
| DINOv2-S/M | Text + JS | 279,809 | 0.988 | 0.984 | 0.308 | 0.337 | 0.346 | 0.297 |
| MedSigLIP/B | Fixed | 296,159 | 0.993 | 0.993 | 0.241 | 0.241 | 0.241 | 0.241 |
| MedSigLIP/B | Random ID | 296,193 | 0.992 | 0.992 | 0.251 | 0.251 | 0.251 | 0.258 |
| MedSigLIP/B | Text | 296,193 | 0.991 | 0.982 | 0.287 | 0.305 | 0.312 | 0.292 |
| MedSigLIP/B | Text + JS | 296,193 | 0.991 | 0.982 | 0.288 | 0.306 | 0.313 | 0.293 |
| MedSigLIP/M | Fixed | 378,087 | 0.991 | 0.991 | 0.263 | 0.263 | 0.263 | 0.274 |
| MedSigLIP/M | Random ID | 378,113 | 0.990 | 0.990 | 0.265 | 0.265 | 0.265 | 0.270 |
| MedSigLIP/M | Text | 378,113 | 0.992 | 0.990 | 0.231 | 0.252 | 0.259 | 0.249 |
| MedSigLIP/M | Text + JS | 378,113 | 0.992 | 0.989 | 0.255 | 0.274 | 0.281 | 0.271 |
| RETFound-Green/M | Fixed | 279,783 | 0.984 | 0.984 | 0.340 | 0.340 | 0.340 | 0.330 |

| Visual / tower | Head | Params | AUC<sub>C</sub> | AUC<sub>H</sub> | B<sub>C</sub> | B<sub>H</sub> | B<sub>T</sub> | B<sub>A</sub> |
| --- | --- | --- | --- | --- | --- | --- | --- | --- |
| RETFound-Green/M | Random ID | 279,809 | 0.983 | 0.983 | 0.324 | 0.324 | 0.324 | 0.323 |
| RETFound-Green/M | Text | 279,809 | 0.982 | 0.978 | 0.319 | 0.329 | 0.337 | 0.328 |
| RETFound-Green/M | Text + JS | 279,809 | 0.982 | 0.978 | 0.319 | 0.328 | 0.337 | 0.329 |
| RETFound-OCT/M | Fixed | 361,703 | 0.985 | 0.985 | 0.294 | 0.294 | 0.294 | 0.294 |
| RETFound-OCT/M | Random ID | 361,729 | 0.980 | 0.980 | 0.348 | 0.348 | 0.348 | 0.334 |
| RETFound-OCT/M | Text | 361,729 | 0.984 | 0.981 | 0.333 | 0.348 | 0.357 | 0.323 |
| RETFound-OCT/M | Text + JS | 361,729 | 0.984 | 0.981 | 0.313 | 0.328 | 0.337 | 0.319 |

**Table 52:** Paired conditional 95% intervals for the text arm. Effects are summed-class Brier differences, averaged over the three heldout descriptions within each image before 2,000 cluster resamples. Values are multiplied by 10^3^. Wording: held-out minus canonical. Recalibration: new-schema temperature minus transferred canonical temperature. Each row is one fixed fit, not an independent cohort.

| Task | Visual/text | Seed | Wording | 95% CI | Recalibration [95% CI] |
| --- | --- | --- | --- | --- | --- |
| F | DINOv2-B/B | 2027 | 231.19 | [229.49, 232.91] | -79.81 [-80.40, -79.23] |
| F | DINOv2-B/B | 2028 | 314.96 | [312.71, 317.31] | -164.42 [-165.57, -163.30] |
| F | DINOv2-B/B | 2029 | 210.36 | [208.73, 211.97] | -59.28 [-59.70, -58.88] |
| F | DINOv2-B/M | 2027 | 37.72 | [36.51, 38.98] | -37.88 [-39.13, -36.66] |
| F | DINOv2-B/M | 2028 | 54.60 | [53.24, 55.97] | -54.63 [-56.07, -53.22] |
| F | DINOv2-B/M | 2029 | 40.13 | [38.77, 41.43] | -39.87 [-41.18, -38.52] |
| F | MedSigLIP/B | 2027 | 156.93 | [155.88, 157.90] | -2.63 [-2.93, -2.32] |
| F | MedSigLIP/B | 2028 | 289.29 | [287.48, 291.23] | -136.13 [-137.05, -135.28] |
| F | MedSigLIP/B | 2029 | 255.21 | [253.56, 256.90] | -101.82 [-102.48, -101.17] |
| F | MedSigLIP/M | 2027 | 39.64 | [38.43, 40.78] | -39.68 [-40.81, -38.45] |
| F | MedSigLIP/M | 2028 | 29.48 | [28.31, 30.70] | -29.99 [-31.17, -28.83] |
| F | MedSigLIP/M | 2029 | 33.06 | [31.87, 34.32] | -33.00 [-34.24, -31.82] |
| O | DINOv2-B/B | 2027 | 0.83 | [-5.87, 7.46] | 2.97 [2.13, 3.79] |
| O | DINOv2-B/B | 2028 | -1.35 | [-8.27, 4.79] | 4.64 [3.46, 5.86] |
| O | DINOv2-B/B | 2029 | 10.95 | [5.18, 16.63] | 3.41 [2.30, 4.50] |
| O | DINOv2-B/M | 2027 | 10.33 | [7.69, 13.17] | 7.94 [5.95, 10.09] |
| O | DINOv2-B/M | 2028 | 15.37 | [12.27, 18.53] | 6.74 [4.88, 8.59] |
| O | DINOv2-B/M | 2029 | 18.19 | [15.10, 21.55] | 6.37 [4.52, 8.24] |
| O | MedSigLIP/B | 2027 | 13.93 | [7.47, 20.72] | 5.90 [4.65, 7.18] |
| O | MedSigLIP/B | 2028 | 24.56 | [17.20, 31.85] | 8.10 [6.25, 9.82] |
| O | MedSigLIP/B | 2029 | 15.26 | [6.89, 24.41] | 7.34 [5.61, 9.14] |
| O | MedSigLIP/M | 2027 | 22.52 | [18.69, 26.72] | 8.26 [5.97, 10.64] |
| O | MedSigLIP/M | 2028 | 18.23 | [14.88, 21.95] | 6.39 [4.11, 8.63] |
| O | MedSigLIP/M | 2029 | 22.60 | [18.75, 26.52] | 5.83 [3.92, 7.83] |

### 11.1 Complete development-cohort adapter comparisons

**Table 53:**
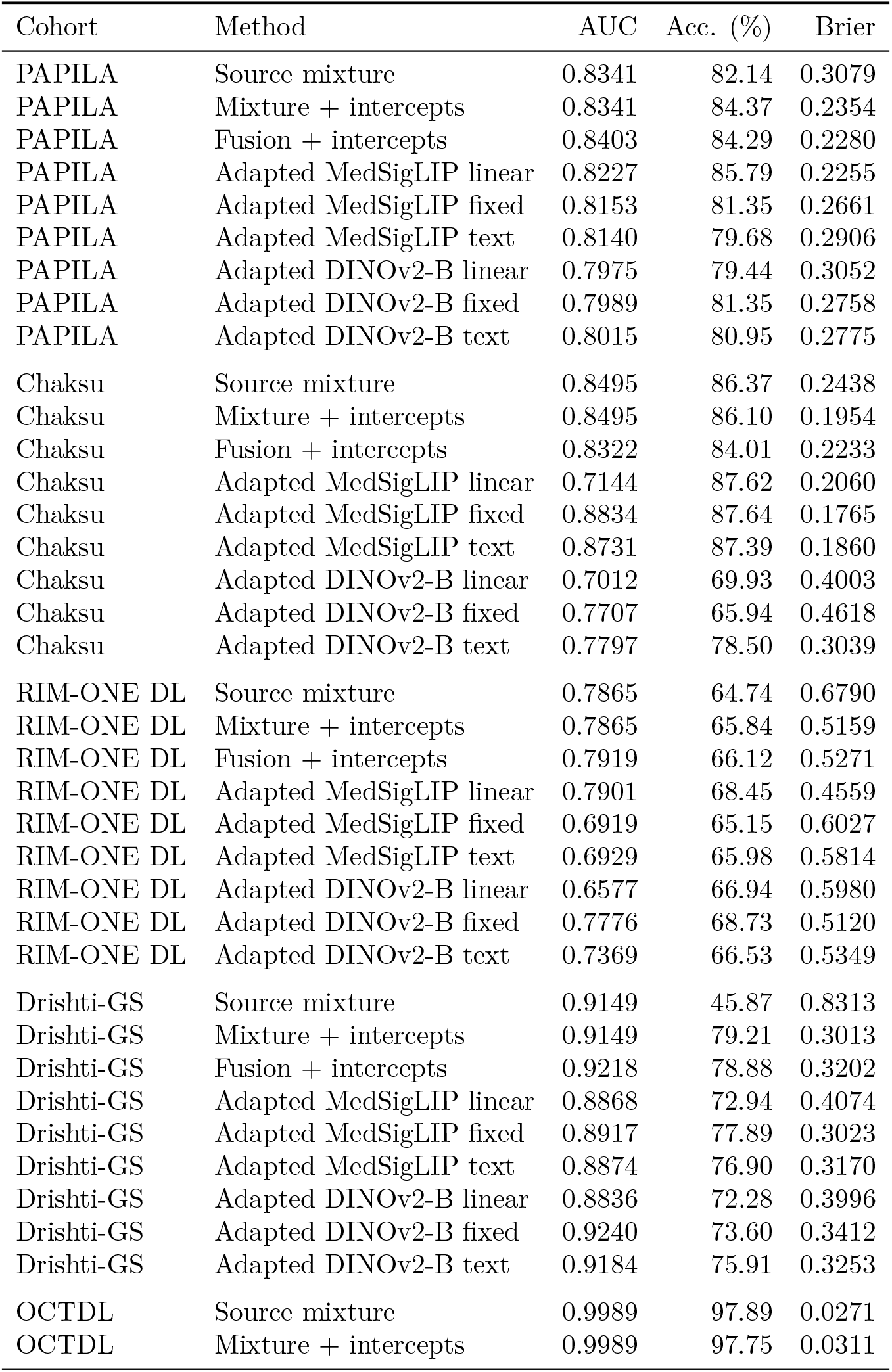
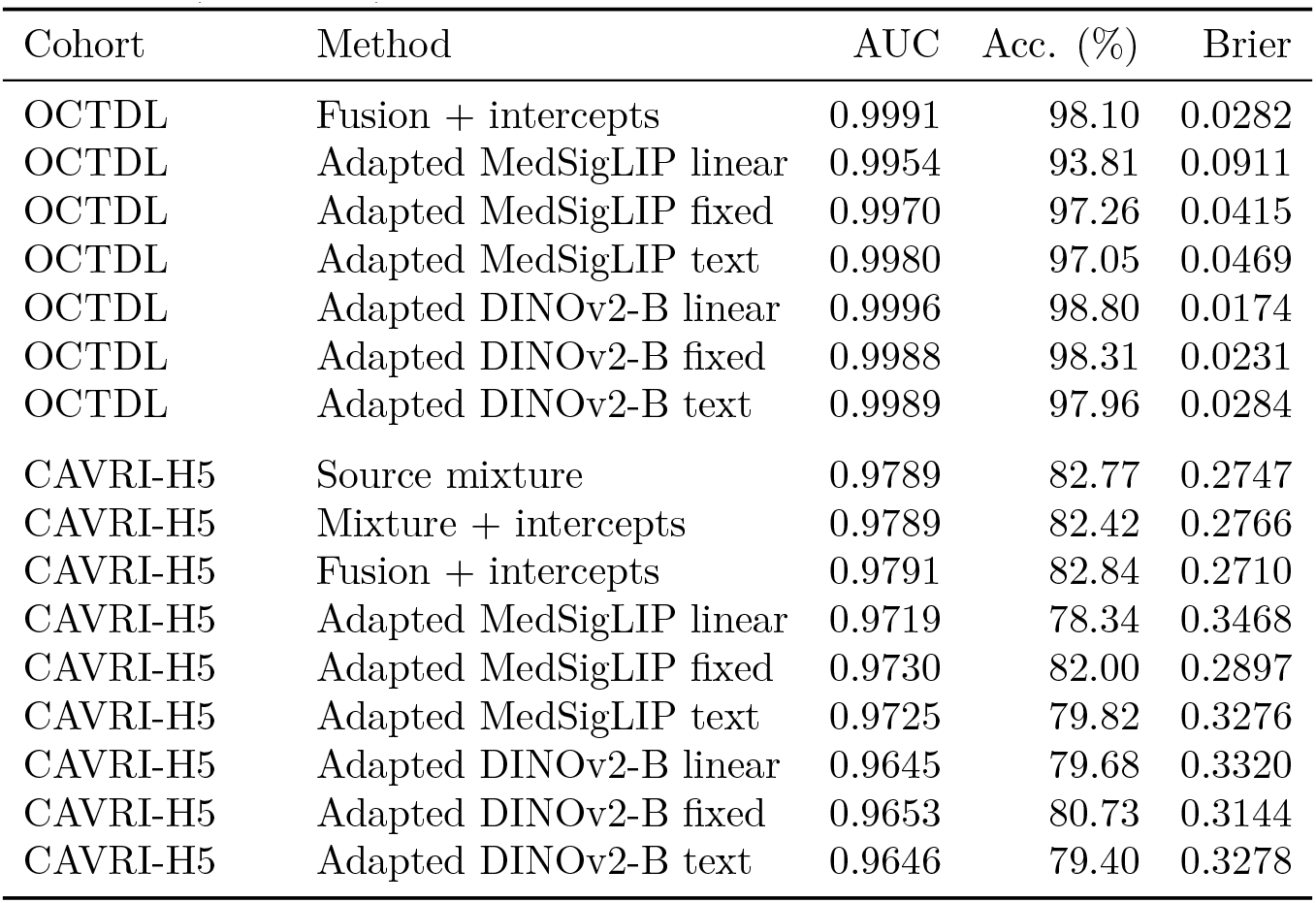
Complete development-cohort transfer matrix. All 54 rows are means of three seed metrics. These six cohorts were inspected before adapter design; leave-one-cohort-out (LOCO) outcomes are exploratory development results. The source mixture uses no external fitting. Adapted methods exclude the evaluated cohort; Chaksu is excluded from all fitting because its suspect end-point is not the clinical adapter target. The six adapted single experts are post-hoc controls with the same permitted supervision and frozen source-temperature convention. Fundus endpoints retain each dataset’s released label definition; OCTDL evaluates conditional DME/normal probabilities, whereas CAVRI-H5 evaluates the four supported classes and excludes VMA–VMT. Brier loss sums over endpoint classes and should not be compared directly across different class sets.

| Cohort | Method | AUC | Acc. (%) | Brier |
| --- | --- | --- | --- | --- |
| PAPILA | Source mixture | 0.8341 | 82.14 | 0.3079 |
| PAPILA | Mixture + intercepts | 0.8341 | 84.37 | 0.2354 |
| PAPILA | Fusion + intercepts | 0.8403 | 84.29 | 0.2280 |
| PAPILA | Adapted MedSigLIP linear | 0.8227 | 85.79 | 0.2255 |
| PAPILA | Adapted MedSigLIP fixed | 0.8153 | 81.35 | 0.2661 |
| PAPILA | Adapted MedSigLIP text | 0.8140 | 79.68 | 0.2906 |
| PAPILA | Adapted DINOv2-B linear | 0.7975 | 79.44 | 0.3052 |
| PAPILA | Adapted DINOv2-B fixed | 0.7989 | 81.35 | 0.2758 |
| PAPILA | Adapted DINOv2-B text | 0.8015 | 80.95 | 0.2775 |
| Chaksu | Source mixture | 0.8495 | 86.37 | 0.2438 |
| Chaksu | Mixture + intercepts | 0.8495 | 86.10 | 0.1954 |
| Chaksu | Fusion + intercepts | 0.8322 | 84.01 | 0.2233 |
| Chaksu | Adapted MedSigLIP linear | 0.7144 | 87.62 | 0.2060 |
| Chaksu | Adapted MedSigLIP fixed | 0.8834 | 87.64 | 0.1765 |
| Chaksu | Adapted MedSigLIP text | 0.8731 | 87.39 | 0.1860 |
| Chaksu | Adapted DINOv2-B linear | 0.7012 | 69.93 | 0.4003 |
| Chaksu | Adapted DINOv2-B fixed | 0.7707 | 65.94 | 0.4618 |
| Chaksu | Adapted DINOv2-B text | 0.7797 | 78.50 | 0.3039 |
| RIM-ONE DL | Source mixture | 0.7865 | 64.74 | 0.6790 |
| RIM-ONE DL | Mixture + intercepts | 0.7865 | 65.84 | 0.5159 |
| RIM-ONE DL | Fusion + intercepts | 0.7919 | 66.12 | 0.5271 |
| RIM-ONE DL | Adapted MedSigLIP linear | 0.7901 | 68.45 | 0.4559 |
| RIM-ONE DL | Adapted MedSigLIP fixed | 0.6919 | 65.15 | 0.6027 |
| RIM-ONE DL | Adapted MedSigLIP text | 0.6929 | 65.98 | 0.5814 |
| RIM-ONE DL | Adapted DINOv2-B linear | 0.6577 | 66.94 | 0.5980 |
| RIM-ONE DL | Adapted DINOv2-B fixed | 0.7776 | 68.73 | 0.5120 |
| RIM-ONE DL | Adapted DINOv2-B text | 0.7369 | 66.53 | 0.5349 |
| Drishti-GS | Source mixture | 0.9149 | 45.87 | 0.8313 |
| Drishti-GS | Mixture + intercepts | 0.9149 | 79.21 | 0.3013 |
| Drishti-GS | Fusion + intercepts | 0.9218 | 78.88 | 0.3202 |
| Drishti-GS | Adapted MedSigLIP linear | 0.8868 | 72.94 | 0.4074 |
| Drishti-GS | Adapted MedSigLIP fixed | 0.8917 | 77.89 | 0.3023 |
| Drishti-GS | Adapted MedSigLIP text | 0.8874 | 76.90 | 0.3170 |
| Drishti-GS | Adapted DINOv2-B linear | 0.8836 | 72.28 | 0.3996 |
| Drishti-GS | Adapted DINOv2-B fixed | 0.9240 | 73.60 | 0.3412 |
| Drishti-GS | Adapted DINOv2-B text | 0.9184 | 75.91 | 0.3253 |
| OCTDL | Source mixture | 0.9989 | 97.89 | 0.0271 |
| OCTDL | Mixture + intercepts | 0.9989 | 97.75 | 0.0311 |
| OCTDL | Fusion + intercepts | 0.9991 | 98.10 | 0.0282 |
| OCTDL | Adapted MedSigLIP linear | 0.9954 | 93.81 | 0.0911 |
| OCTDL | Adapted MedSigLIP fixed | 0.9970 | 97.26 | 0.0415 |
| OCTDL | Adapted MedSigLIP text | 0.9980 | 97.05 | 0.0469 |
| OCTDL | Adapted DINOv2-B linear | 0.9996 | 98.80 | 0.0174 |
| OCTDL | Adapted DINOv2-B fixed | 0.9988 | 98.31 | 0.0231 |
| OCTDL | Adapted DINOv2-B text | 0.9989 | 97.96 | 0.0284 |
| CAVRI-H5 | Source mixture | 0.9789 | 82.77 | 0.2747 |
| CAVRI-H5 | Mixture + intercepts | 0.9789 | 82.42 | 0.2766 |
| CAVRI-H5 | Fusion + intercepts | 0.9791 | 82.84 | 0.2710 |
| CAVRI-H5 | Adapted MedSigLIP linear | 0.9719 | 78.34 | 0.3468 |
| CAVRI-H5 | Adapted MedSigLIP fixed | 0.9730 | 82.00 | 0.2897 |
| CAVRI-H5 | Adapted MedSigLIP text | 0.9725 | 79.82 | 0.3276 |
| CAVRI-H5 | Adapted DINOv2-B linear | 0.9645 | 79.68 | 0.3320 |
| CAVRI-H5 | Adapted DINOv2-B fixed | 0.9653 | 80.73 | 0.3144 |
| CAVRI-H5 | Adapted DINOv2-B text | 0.9646 | 79.40 | 0.3278 |

#### 12 Interpretation of the empirical reader target

For two binary observations, *q_i_* has positive-class values 0, 1*/*2, or 1. Disagreement therefore produces a target at 1*/*2 without asserting that either grader is correct or that referral probability equals 1*/*2. The released final label can disagree with either individual reader and remains a separate endpoint. Equation 5 follows by expanding ||*p*-*e_r_*||^2^_2_ and substituting E[*e_r_*] = *q*; it is verified numerically for all retained reader-analysis predictions. The ambiguity term is zero for observed agreement and ½ for observed binary disagreement. Its empirical magnitude depends on the eligible-case definition and should not be generalized to missing or ungradable cases.

Positive temperature preserves binary score ordering and symmetric-uncertainty ranking. Changing its target therefore does not create a new disagreement-routing model; a distinct human-review policy requires separate evaluation.

## 13 Retrospective clinical text and ophthalmic question extension

This extension was added after the imaging study. Its original text protocol was fixed before text-model outcomes; the adaptive-mixture amendment was introduced after the equal-mixture test results were inspected. No neural checkpoint was fine-tuned. Clinical labels are the source visual-field-defined endpoint; documentation and treatment can reveal existing disease. Reported clinical-note probabilities are not validated future-risk estimates.

All common model inputs have distinct normalized hashes across retained partitions. A train-fitted word unigram/bigram TF–IDF audit finds 30 test notes with maximum train cosine similarity above 0.95 and 4 above 0.99. This diagnostic does not justify post-outcome exclusions. Shared templates and unknown pretraining overlap remain limitations. Yale-BIDS-Chen LMOD contained no data files at review; the released M3LLM pure-text benchmark contained 49 items and no matches to the fixed ophthalmic vocabulary, so it was not added. The publicly available MedMCQA repack was superseded by the original subject-labelled resource. Public benchmark contamination cannot be excluded for any pretrained model.

---

**Table 54:** Complete text comparisons. Calibrated and mixture rows report mean metrics across three development-partition seeds. Uniform probabilities use deterministic first-option argmax; their accuracy is not expected random-choice accuracy. Keyword probabilities are hard 0/1, with NLL clipped at 10*^−^*^12^. No poor-performing arm is omitted.

| Resource | Model | N | Accuracy | AUROC | Brier | NLL | ECE | AURC |
| --- | --- | --- | --- | --- | --- | --- | --- | --- |
| fairvlmed | uniform | 1996 | 0.4885 | 0.5000 | 0.5000 | 0.6931 | 0.0115 | 0.5080 |
| fairvlmed | startlux_0.8B | 1996 | 0.5501 | 0.5971 | 0.5639 | 0.7927 | 0.1749 | 0.4146 |
| fairvlmed | startlux_2B | 1996 | 0.6127 | 0.6607 | 0.5359 | 0.8064 | 0.1651 | 0.3538 |
| fairvlmed | tfidf | 1996 | 0.7836 | 0.8688 | 0.3064 | 0.4718 | 0.0765 | 0.0908 |
| fairvlmed | ophthabert | 1996 | 0.7530 | 0.8499 | 0.3260 | 0.5072 | 0.0758 | 0.1138 |
| fairvlmed | startlux_0.8B_calibrated | 1996 | 0.5501 | 0.5971 | 0.4914 | 0.6845 | 0.0172 | 0.4146 |
| fairvlmed | startlux_2B_calibrated | 1996 | 0.6127 | 0.6607 | 0.4716 | 0.6645 | 0.0408 | 0.3538 |
| fairvlmed | tfidf_calibrated | 1996 | 0.7836 | 0.8688 | 0.2923 | 0.4448 | 0.0112 | 0.0908 |
| fairvlmed | ophthabert_calibrated | 1996 | 0.7530 | 0.8499 | 0.3161 | 0.4809 | 0.0315 | 0.1138 |
| fairvlmed | ophjev_text_equal_fusion | 1996 | 0.7540 | 0.8504 | 0.3564 | 0.5406 | 0.1131 | 0.1193 |
| fairvlmed | diagnosis_keyword | 1996 | 0.4770 | 0.4738 | 1.0461 | 14.4523 | 0.5230 | 0.5305 |
| medmcqa_eye | uniform | 43 | 0.3023 | – | 0.7500 | 1.3863 | 0.0523 | 0.6668 |
| medmcqa_eye | startlux_0.8B | 43 | 0.4651 | – | 0.6383 | 1.1656 | 0.2088 | 0.3852 |
| medmcqa_eye | startlux_2B | 43 | 0.4186 | – | 0.7299 | 1.3289 | 0.3419 | 0.4655 |
| diagnosisarena_eye | uniform | 48 | 0.2500 | – | 0.7500 | 1.3863 | 0.0000 | 0.8053 |
| diagnosisarena_eye | startlux_0.8B | 48 | 0.5000 | – | 0.6344 | 1.1881 | 0.1210 | 0.3360 |
| diagnosisarena_eye | startlux_2B | 48 | 0.5417 | – | 0.6176 | 1.1604 | 0.1154 | 0.3931 |
| fairvlmed | Exploratory validation-fit fusion | 1996 | 0.7886 | 0.8759 | 0.2853 | 0.4367 | 0.0218 | 0.0854 |

**Table 55:** Mechanistic controls. Masking covers glaucoma, POAG, PACG, NTG, OAG, COAG and LTG tokens; examination values, treatments, and indirect proxies remain. The small fixed note subset supports descriptive analysis only.

| Resource | Model | Intervention | N | Original Acc. | Changed Acc. | Flip rate | Mean TV |
| --- | --- | --- | --- | --- | --- | --- | --- |
| fairvlmed | startlux_0.8B | reversed | 64 | 0.5312 | 0.5938 | 0.0625 | 0.0172 |
| fairvlmed | startlux_0.8B | no_evidence | 64 | 0.5312 | 0.5000 | 0.3125 | 0.2716 |
| fairvlmed | startlux_0.8B | masked_diagnosis | 64 | 0.5312 | 0.5156 | 0.2969 | 0.1521 |
| fairvlmed | startlux_2B | reversed | 64 | 0.6094 | 0.6250 | 0.0156 | 0.0328 |
| fairvlmed | startlux_2B | no_evidence | 64 | 0.6094 | 0.5000 | 0.2344 | 0.2896 |
| fairvlmed | startlux_2B | masked_diagnosis | 64 | 0.6094 | 0.5312 | 0.1719 | 0.1230 |
| fairvlmed | tfidf | no_evidence | 64 | 0.8125 | 0.5000 | 0.5625 | 0.2393 |
| fairvlmed | tfidf | masked_diagnosis | 64 | 0.8125 | 0.7969 | 0.0156 | 0.0092 |
| fairvlmed | ophthabert | no_evidence | 64 | 0.7656 | 0.5000 | 0.5469 | 0.4984 |
| fairvlmed | ophthabert | masked_diagnosis | 64 | 0.7656 | 0.7500 | 0.0156 | 0.0244 |
| medmcqa_eye | startlux_0.8B | reversed | 43 | 0.4651 | 0.4884 | 0.1395 | 0.0601 |
| medmcqa_eye | startlux_0.8B | no_evidence | 43 | 0.4651 | 0.3488 | 0.6512 | 0.1829 |
| medmcqa_eye | startlux_2B | reversed | 43 | 0.4186 | 0.4651 | 0.3256 | 0.1544 |
| medmcqa_eye | startlux_2B | no_evidence | 43 | 0.4186 | 0.1860 | 0.6512 | 0.3368 |
| diagnosisarena_eye | startlux_0.8B | reversed | 48 | 0.5000 | 0.4792 | 0.2917 | 0.1017 |
| diagnosisarena_eye | startlux_0.8B | no_evidence | 48 | 0.5000 | 0.3125 | 0.5625 | 0.2065 |
| diagnosisarena_eye | startlux_2B | reversed | 48 | 0.5417 | 0.5833 | 0.3125 | 0.1785 |
| diagnosisarena_eye | startlux_2B | no_evidence | 48 | 0.5417 | 0.1458 | 0.6875 | 0.3904 |

**Table 56:** Exploratory development-selected mixture. Weights are nonnegative and sum to one; fitting uses one development half, with penalty selection on the other. Test outcomes do not select weights. The procedure was designed after original test inspection and needs independent replication.

| Seed | Penalty | TF-IDF | OphthaBERT | StartLux 2B | Selection Brier | Test Brier |
| --- | --- | --- | --- | --- | --- | --- |
| 2027 | 0 | 0.8478 | 0.1522 | 0.0000 | 0.2808 | 0.2862 |
| 2028 | 0.01 | 0.8108 | 0.1892 | 0.0000 | 0.2801 | 0.2853 |
| 2029 | 0 | 0.7778 | 0.2222 | 0.0000 | 0.2807 | 0.2844 |

**Table 57:**
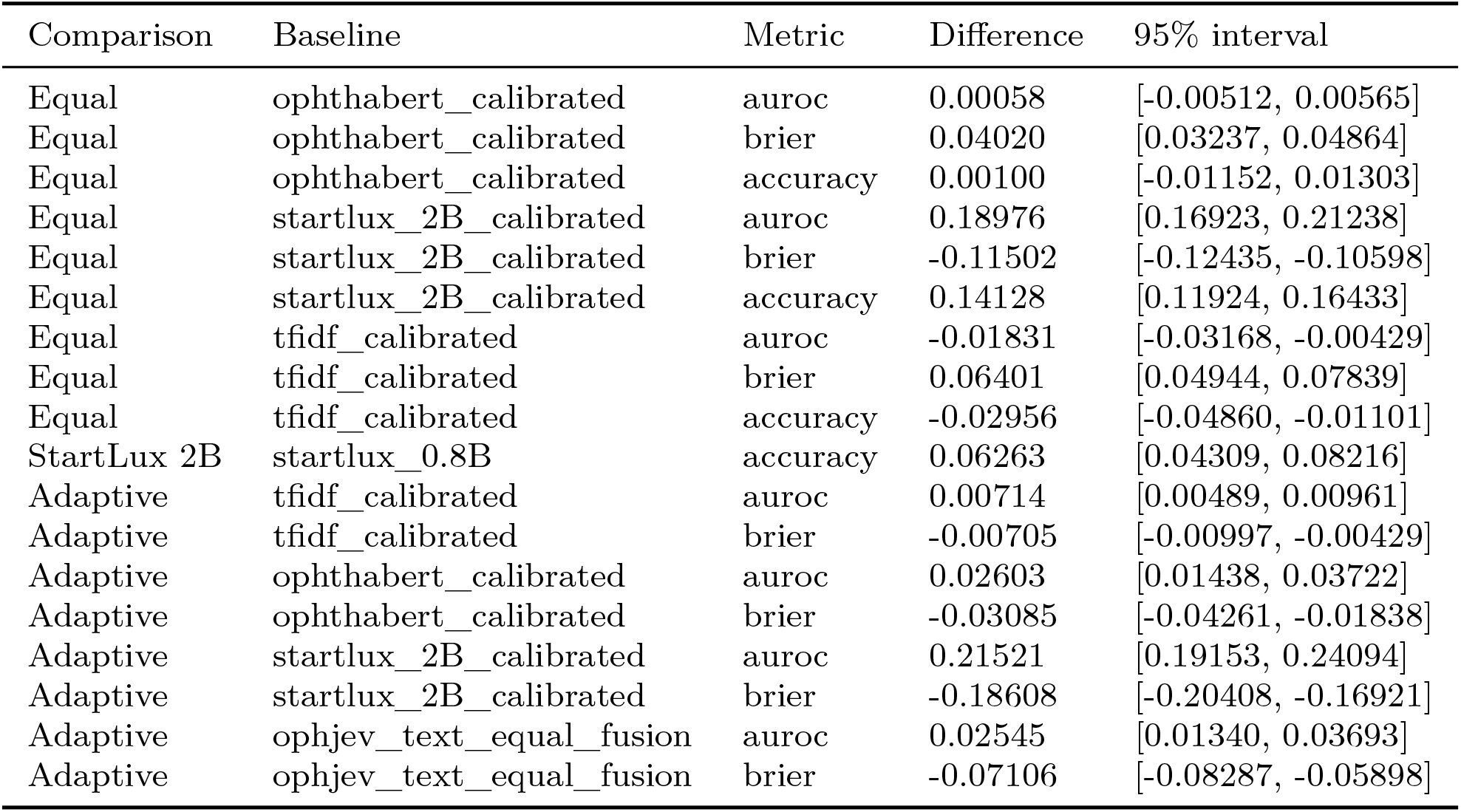
Paired subject-bootstrap differences. Adaptive refers to the exploratory validation-fit fusion. Equal rows include a separately labeled StartLux-size comparison in the underlying machine-readable results; three development partitions are not treated as independent subjects.

**Table 58:** Descriptive subgroup audit for race. These results do not establish fairness equivalence; no subgroup-specific fitting was performed.

| Attribute | Group | Model | N | AUROC | Brier |
| --- | --- | --- | --- | --- | --- |
| race | asian | tfidf_calibrated | 157 | 0.9228 | 0.2215 |
| race | asian | ophthabert_calibrated | 157 | 0.9130 | 0.2454 |
| race | asian | startlux_2B_calibrated | 157 | 0.7092 | 0.4582 |
| race | asian | ophjev_text_equal_fusion | 157 | 0.9091 | 0.3093 |
| race | black | tfidf_calibrated | 305 | 0.8805 | 0.2814 |
| race | black | ophthabert_calibrated | 305 | 0.8444 | 0.3165 |
| race | black | startlux_2B_calibrated | 305 | 0.6578 | 0.4848 |
| race | black | ophjev_text_equal_fusion | 305 | 0.8370 | 0.3645 |
| race | white | tfidf_calibrated | 1534 | 0.8619 | 0.3016 |
| race | white | ophthabert_calibrated | 1534 | 0.8436 | 0.3232 |
| race | white | startlux_2B_calibrated | 1534 | 0.6497 | 0.4699 |
| race | white | ophjev_text_equal_fusion | 1534 | 0.8445 | 0.3594 |

**Table 59:** Descriptive subgroup audit for gender. These results do not establish fairness equivalence; no subgroup-specific fitting was performed.

| Attribute | Group | Model | N | AUROC | Brier |
| --- | --- | --- | --- | --- | --- |
| gender | female | tfidf_calibrated | 1093 | 0.8692 | 0.2916 |
| gender | female | ophthabert_calibrated | 1093 | 0.8510 | 0.3151 |
| gender | female | startlux_2B_calibrated | 1093 | 0.6508 | 0.4757 |
| gender | female | ophjev_text_equal_fusion | 1093 | 0.8519 | 0.3576 |
| gender | male | tfidf_calibrated | 903 | 0.8686 | 0.2930 |
| gender | male | ophthabert_calibrated | 903 | 0.8488 | 0.3172 |
| gender | male | startlux_2B_calibrated | 903 | 0.6739 | 0.4659 |
| gender | male | ophjev_text_equal_fusion | 903 | 0.8493 | 0.3546 |

**Table 60:** Descriptive subgroup audit for truncated_words. These results do not establish fairness equivalence; no subgroup-specific fitting was performed.

| Attribute | Group | Model | N | AUROC | Brier |
| --- | --- | --- | --- | --- | --- |
| truncated_words | False | tfidf_calibrated | 1374 | 0.8749 | 0.2857 |
| truncated_words | False | ophthabert_calibrated | 1374 | 0.8554 | 0.3106 |
| truncated_words | False | startlux_2B_calibrated | 1374 | 0.6659 | 0.4648 |
| truncated_words | False | ophjev_text_equal_fusion | 1374 | 0.8554 | 0.3507 |
| truncated_words | True | tfidf_calibrated | 622 | 0.8568 | 0.3067 |
| truncated_words | True | ophthabert_calibrated | 622 | 0.8338 | 0.3282 |
| truncated_words | True | startlux_2B_calibrated | 622 | 0.6370 | 0.4855 |
| truncated_words | True | ophjev_text_equal_fusion | 622 | 0.8349 | 0.3686 |

